# A genome-wide association study of young onset Parkinson’s disease in European ancestry

**DOI:** 10.64898/2026.06.03.26354799

**Authors:** Ingeborg Haugesag Lie, Janna van Wetering, Miko Valori, Kajsa Atterling Brolin, Kathryn Step, Claudia Schulte, Hirotaka Iwaki, Sara Bandres-Ciga, Hampton Leonard, Manu Sharma, the International Parkinson’s Disease Genomics Consortium the Global Parkinson’s Genetics Program, Andrew Singleton, Lasse Pihlstrøm

## Abstract

Young onset Parkinson’s disease may be caused by biallelic mutations in *PRKN* or other autosomal recessive Parkinson’s disease genes, but the majority of patients do not carry known monogenic variants. Previous studies have found an increased cumulative burden of common genetic risk variants for Parkinson’s disease in young onset patients, but the specific genetic architecture of non-monogenic young onset Parkinson’s disease is not well characterized.

We conducted a genome-wide association study of 1,528 Parkinson’s disease patients with symptom onset between 18 and 40 years and 20,408 controls of European ancestry using data from The Global Parkinson’s Genetic Program, the International Parkinson’s Disease Genomics Consortium, and the NeuroGenetics Research Consortium. We performed meta-analyses of additive and recessive regression models and investigated associations between age at onset groups and different polygenic risk scores.

An additive model meta-analysis identified six independent loci passing a genome-wide significance threshold, including three loci identified in previous genome-wide association studies (near *SNCA*, *GBA1,* and *HIP1R*) and two loci not previously associated with Parkinson’s disease (rs74950462, *P* = 1.24×10^-8^ and rs72848817, *P* = 4.89×10^-8^). Furthermore, we identified a significant signal at the *PRKN* locus, prompting a follow-up analysis employing a recessive model. The recessive genome-wide association meta-analysis identified nine loci passing a genome-wide significance threshold, including *SNCA*, *PRKN,* and seven novel variants. Patients with onset between 18 and 40 years had significantly higher polygenic risk scores than later onset patients when the score was modelled specifically on genome-wide association statistics from independent young onset Parkinson’s disease participants versus healthy controls. This increased polygenic burden was driven in part by loci harbouring mitochondrial pathway genes.

Our results indicate that previously unidentified common and low-frequency variants contribute specifically to the young onset subgroup of Parkinson’s disease. Association signals detected uniquely with a recessive model suggest that genetic susceptibility to young onset Parkinson’s disease may be partially driven by homozygous variation, in line with previous reports of increased runs of homozygosity in this particular group of patients and may be consistent with a loss of function mechanism. The findings support the notion of young onset Parkinson’s disease as a partly distinct subphenotype and highlight the mitochondrial pathway. These results may have implications for future precision medicine but should be interpreted with caution pending independent replication.

## Introduction

Parkinson’s disease (PD) is a neurodegenerative disorder showing increasing prevalence with older age.^1^ In population-based cohorts, mean age at onset is commonly 60-65 years or more^2^, whereas only 5-7% have onset before the age of 40.^3^ In 1987, Quinn, Critchley and Marsden proposed that apparent idiopathic PD beginning between the age of 21 and 39 years should be called young onset PD (YOPD), hypothesizing that this group of patients represent the lower end of a skewed age at onset distribution for idiopathic PD.^4^ Subsequently however, a number of genes that harbour mutations causing autosomal recessive forms of PD have been identified, including *PRKN*^5^, *DJ-1*^6^, *PINK1*^7^ and *VPS13C.*^8^ Biallelic loss of function variants in these genes lead to a phenotype of levodopa-responsive PD, typically with early onset.^8,9^

Multiple genetic screening studies have been performed in younger PD patients, where the fraction of monogenic cases identified depends on the age cutoff.^10–14^ Biallelic pathogenic variants in *PRKN* are the most common cause of autosomal recessive PD^9,10^ and were reported in 17% of PD patients with onset at 21-30 years, 3% with onset at 31-40 years, but only 0.5% with onset 41-50 years in a large screening study.^13^ YOPD patients with negative screening for mutations in genes linked to monogenic forms of the disease may still have an unidentified genetic cause, a hypothesis supported by observations of increased runs of homozygosity in YOPD patients^15,16^ Furthermore, the clinical phenotype of mutation negative YOPD resembles that of *PRKN* carriers, with good levodopa-response, prominent dystonia and early fluctuations, yet limited cognitive involvement.^17,18^

Nevertheless, despite intensive efforts to identify novel causes of monogenic PD, the majority of YOPD patients remain genetically unexplained. Genome-wide association studies (GWASs) have identified more than 150 susceptibility loci for sporadic, idiopathic PD of all ages.^19,20^ Multiple studies of PD polygenic risk scores (PRSs) capturing the cumulative effect of these variants have shown that a higher genetic burden of common low-risk variants is associated with earlier age at onset.^21,22^ A study comparing PRSs across age at onset strata found the highest PRS in the age at onset 31-40 group^21^, an observation that fits well with Quinn, Critchley, and Marsden’s original hypothesis about the lower end of the age at onset distribution for idiopathic PD.^4^ A higher PRS among PD subjects without a known monogenic cause compared to subjects with a known monogenic cause has been reported in an Indian study population with AAO **≤**50.^23^

In this study, we aimed to further elucidate the genetic architecture of YOPD. We hypothesized that the common genetic risk variants contributing to YOPD, and the relative importance of these variants, may not be identical to those of late onset PD, which has made up the majority of PD participants in previous PD GWASs. Taking advantage of large-scale genetic datasets from international consortia, we performed GWASs and meta-analyses of YOPD in participants of European ancestry, comparing the findings with known association signals from general PD GWASs. Our results support the hypothesis that non-monogenic YOPD is associated with genetic risk factors that not only mirror later-onset PD but are also partly distinct, an insight that may have implications for future translational research and precision medicine in PD.

## Methods

### Subjects

We analysed patient and control data of general European ancestry from three different sources. From the International Parkinson’s Disease Genomics Consortium (IPDGC)^24^, we took advantage of data previously compiled for a GWAS of age at onset in PD, where inclusion criteria have been described in detail elsewhere.^25^ We included only subcohorts composed of both patients and healthy control subjects. Next, we analysed data from PD and control participants without neurological disease of European ancestry from the complex network in Global Parkinson’s Genetics Program (GP2) data release 10.^26,27^ Selection of GP2 subcohorts and measures to avoid sample overlap with IPDGC data are described in Supplementary Methods. Finally, we included a third independent PD case-control GWAS dataset from the NeuroGenetics Research Consortium (NGRC)^28^, publicly available in dbGAP.

In the GP2 and NGRC datasets, “age at onset” was defined as the age of subjective onset of PD symptoms, and participants lacking this data point were excluded.^26,28^ In the IPDGC data, age at diagnosis was used as a proxy if subjective age at motor symptom onset was not available.^25^ For the main analyses we included patients with age at onset ≥ 18 to ≤ 39 years. Although the classical definition of YOPD is age at onset between 21–39 years, some studies use a pragmatic lower cutoff at 18 years to include all adults.^21^ In the GP2 data, PD participants with age at onset ≥ 40 years from the same cohorts were included for comparison in follow-up analyses. In order to minimize survival bias in the genotype frequencies of controls, we excluded controls with age at sampling ≥ 80 years. Genetic screening has previously been reported for one of the IPDGC cohorts^29^, excluding LRRK2 p.G2019S carriers, but has otherwise not been systematically performed in all subjects.^27,28^ All participants gave written informed consent to participate in the respective studies responsible for the primary data collection.^25–28^

### Genotyping and quality control

Genotyping of IPDGC data was performed using different arrays for different substudies (Supplementary Table 1) and initial quality checks and filtering were performed as described in a previous publication.^25^ Raw data were imputed using Eagle v2.3 imputation^30^ based on the HRC r1.1 2016 reference panel^31^ at the Michigan imputation server.^32^ All GP2 data were genotyped using the Illumina NeuroBooster array^33^. Quality control and ancestry estimation using GenoTools^34^ and imputation on the TOPMed server were performed centrally by the GP2 consortium.^26^ We performed quality filtering and imputation of the NGRC dataset from dbGAP^28^ as described in a previous publication.^35^

Due to differences in genotyping arrays and the availability of raw data across different, isolated databases or cloud environments, GWASes were performed separately for each of the IPDGC subcohorts, GP2, and NGRC. Analyses of relatedness were performed within each of the three main datasets, excluding one individual from each pair of individuals with a relatedness corresponding to first degree cousins or closer.^36^

### Genome-wide association analysis and meta-analysis

In our primary analysis, we used PLINK 2.0^37^ to perform logistic regression with a standard additive model (a versus A), including sex and the first five principal components as covariates. Given that all patients by definition had age < 40, it was not possible to adjust for age in our model. Although age (at onset of patients and at sampling of controls) is commonly included as a covariate in genetic association studies, we expect the exclusion of age from the model to be of minimal consequence, as genotypes do not vary with age, and any potential age-related population stratification will be adjusted for by including principal components. To potentially capture recessive effects, which are particularly relevant in YOPD, we also repeated logistic regression analyses using a recessive model (aa versus AA or Aa). In the recessive model GWAS, we removed SNPs with extreme effect sizes that were likely to represent errors (beta > 4 or < −4) from individual cohort summary statistics prior to meta-analysis.

Meta-analyses were performed with METAL^38^, version released on 2020-05-05, using the inverse variance method, filtered for minor allele frequency (MAF) ≥ 1% and applying genomic control. The genomic inflation parameter λ_1000_ was calculated for the meta-analysis results. Post-meta-analysis filtering of SNPs based on heterogeneity and study-level missingness increased λ_1000_ values and was therefore not included in the workflow (see Supplementary Methods). We defined a standard genome-wide significance threshold of *P* < 5×10^-8^. We used Functional Mapping and Annotation (FUMA) v1.5.2^39^ with default parameters to identify top SNPs and screen for additional independent signals within the significant loci. The results were compared to previously published association signals from the two largest European general PD GWASs^19,20^ using tables and beta-beta-plots.

Due to limitations on integrated data access, additional analyses that depended on individual-level data were conducted using the GP2 data, the largest subcohort. To further explore independent association signals at the *SNCA* locus, we performed a stepwise conditional logistic regression analysis using the same covariates as in the main analysis. Similar to a previously published approach, we included SNPs within a 1Mb region centred on rs356182, and used a significance threshold of *P* < 0.00023, based on an estimated 220 independent tests.^35^ Total common variant heritability was estimated in GP2 data using Genome-wide Complex Trait Analysis (GCTA-GREML), with an assumed prevalence of YOPD at 14.7/100,000, including sex and PC1 to PC5 as covariates.^40–42^

### Polygenic risk score analyses

Polygenic risk scores (PRSs) were generated in the GP2 dataset using PRSice-2 version 2.3.5.^43^ Allelic weights from summary statistics from Nalls et al. 2019^19^ (excluding 23andMe data) were used as general PD-GWAS weights. The more recent summary statistics from Leonard and GP2^20^ were not used due to sample overlap with the test data. YOPD-specific summary stats were generated by meta-analysis of the IPDGC and NGRC datasets in this study, leaving GP2 out as an independent test dataset. Given the limited statistical power of this YOPD-specific reference GWAS, we included signals at a liberal significance threshold of *P* < 0.05. We used the default PRSice-2 linkage disequilibrium clumping thresholds (250 kb window, r^2^ > 0.1) in PRSice-2 and excluded variants with minor allele frequency < 0.01. PRSs were calculated for individuals with age at onset between 18 and 80 years from the included GP2 subcohorts, and participants were stratified into three groups with onset range 18–39, 40–59, and 60–80 years of age, respectively. Group-wise comparisons of PRSs across age at onset groups were evaluated using independent t-tests.

For each GP2 individual, we also calculated pathway-specific PRSs using the same YOPD weights for six selected pathways relevant to PD pathogenesis, as previously described by Tunold et al^44^. T-tests were used to assess differences across age at onset groups, applying Bonferroni correction for six independent tests (significance threshold *P* < 0.0083).

### Runs of homozygosity and whole genome sequencing analyses

To investigate whether recessive GWAS signals were enriched for extended homozygosity, genomic stretches where an individual has inherited identical alleles from both parents, we evaluated runs of homozygosity (ROHs) overlapping genome-wide significant recessive GWAS loci. For each lead GWAS variant, the 5Mb flanking region on both sides of the variant was extracted from imputed genotype data. Analyses were restricted to individuals in the GP2 dataset with available whole-genome sequencing (WGS) data.16 To facilitate robust ROH detection, variants were filtered to include only autosomal SNPs, minor allele frequency ≥ 5%, genotype missingness ≤ 1%, and a minimum inter-marker spacing of 2 kb. ROHs were identified using PLINK v1.937 with a minimum length threshold of 250 kb and allowing up to two heterozygous sites per segment. ROH intervals were intersected with GWAS lead variant positions to identify individuals harbouring ROHs overlapping these loci. For each GWAS variant, genotype data were extracted and individuals homozygous for the risk allele were identified. The ROHs were filtered to include only those individuals homozygous for the GWAS effect allele.

To identify potential recessive coding or regulatory variants not captured by the GWAS, WGS data within 500 kb of recessive GWAS loci were annotated using Ensembl Variant Effect Predictor (VEP, GRCh38).^45^ Annotation included population allele frequencies, functional consequence predictions, and deleteriousness scores. As a complement to the ROH-based analysis, variants were filtered to retain those observed in a homozygous state in at least one case and absent as homozygotes in controls. Additional filters required evidence of potential functional impact, either Combined Annotation-Dependent Depletion (CADD) ≥ 20^46^, SpliceAI ≥ 0.2^47^, or rare exome variant ensemble learner (REVEL)^48^ ≥ 0.5 or a significant impact on the transcript as annotated by VEP. Filtered variants were evaluated for overlap with individuals homozygous for nearby recessive GWAS lead variants, with and without taking ROH data into account.

## Results

### Allelic model genome-wide association analyses

In total, data from 9,548,709 SNPs and 21,936 participants, including 1,528 YOPD subjects and 20,408 control participants, were analysed in our main GWAS meta-analysis. A demographic summary of subjects and datasets is provided in Table 1. In the allelic model GWAS, λ_1000_ was 1.01, indicating minimal inflation of *P*-values (Supplementary Fig. 1). We identified six genomic loci passing the genome-wide significance threshold (Table 2, Fig. 1, Supplementary Fig. 2). Three of these signals correspond to established PD GWAS loci, including *SNCA* (rs356182, *P* = 8.95×10^-18^, *GBA1* (rs2230288, *P* = 2.58×10^-8^) and *HIP1R* (rs10847864, *P* = 2.43×10^-8^).^19,20^ We also observed a significant signal near *PRKN*, a gene known to cause autosomal recessive PD, typically with young onset (rs73035785, *P* = 4.28×10^-8^). This signal showed a high level of heterogeneity across datasets (I^2^ = 81.1%) and was primarily driven by the Spanish IPDGC cohort (Supplementary Fig. 3), corresponding to a finding from a previous Spanish PD GWAS, where the lead SNP in age at onset association analysis was shown to be tagging the pathogenic *PRKN* variant c.155delA.^49^ Finally, we identified 2 novel genome-wide significant association signals, near *DLGAP2* (rs74950462, *P* = 1.24×10^-8^) and *WDR33* (rs72848817, *P* = 4.89×10^-8^), respectively. Both of these novel top-hit variants had a low allele frequency (0.011 and 0.056, respectively) and showed moderate heterogeneity across datasets in the meta-analyses (Table 2, Fig. 2). Estimated liability-scale common variant heritability based on GCTA-GREML in the GP2 dataset was 0.16 (95% CI 0.08-0.24), but we note that our dataset was underpowered for this type of analysis, which also does not capture the effects of rare alleles.

**Figure 1.**
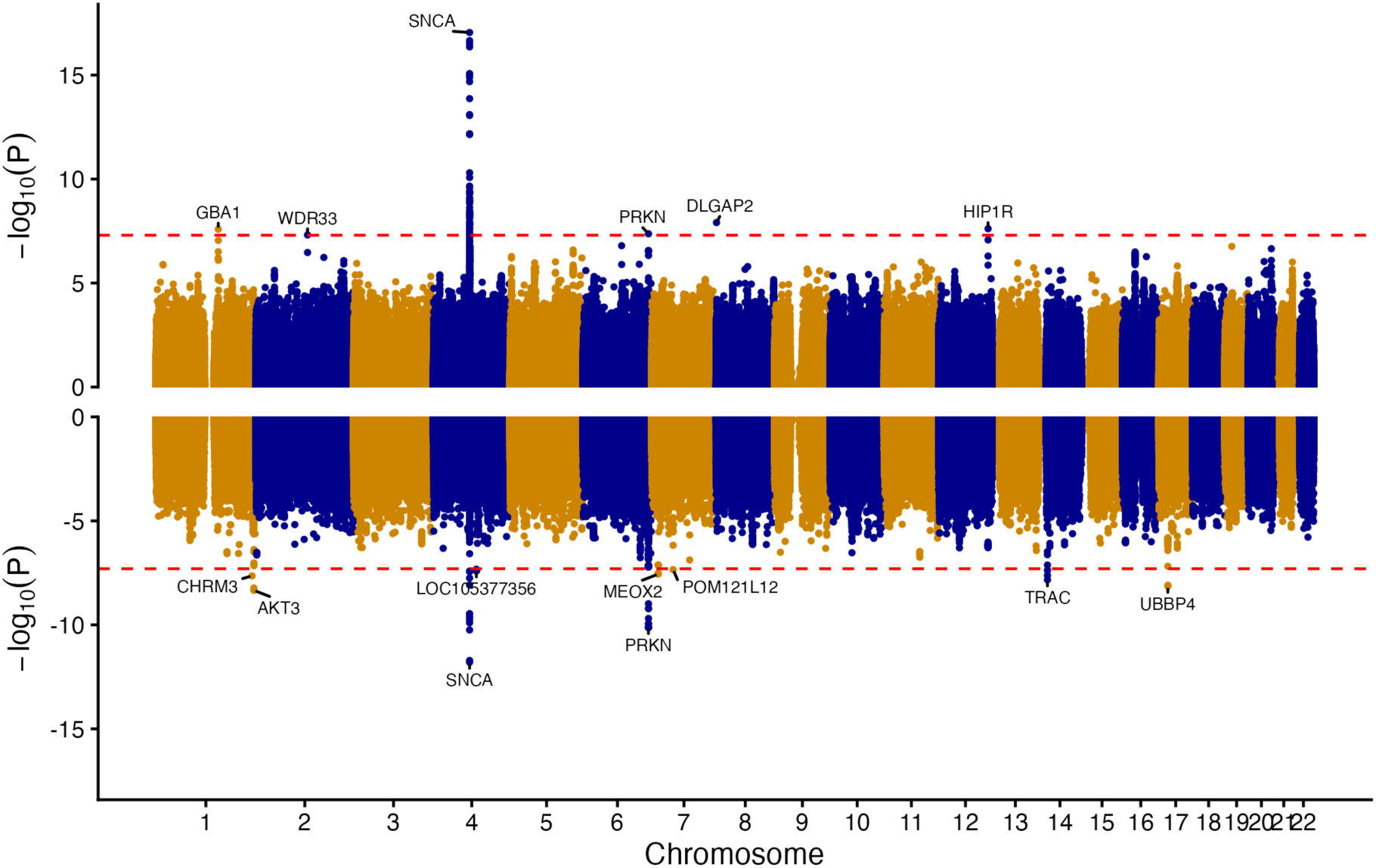
Miami plot of additive and recessive model GWAS results. Colours correspond to autosomes from 1 to 22. Additive model meta-analyses results are shown above, recessive model below, genome wide significant loci are annotated by their closest gene. The dashed red lines indicate the genome-wide significance threshold p< 5×10^-8^.

**Figure 2.**
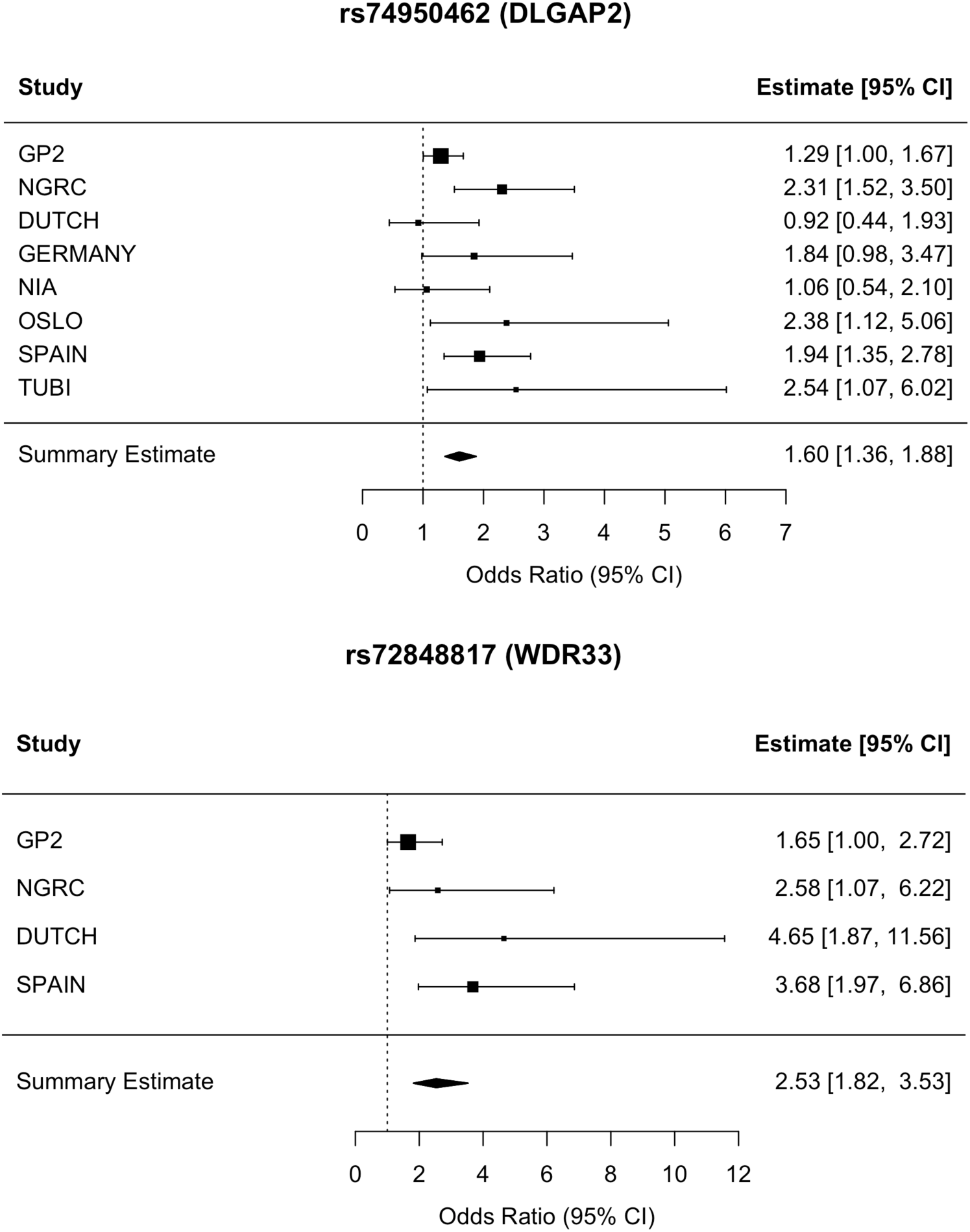
Forest plots. Forest plots of significant SNPs from novel loci identified in the additive meta-analysis.

**Table 1.** Demographic variables for all subcohorts in the meta-analyses.

| Dataset | Ancestry | Participants |  |  | Age controls (Y)<sup>a</sup> |  |  | AAO YOPD (Y) |  |  | Female participants (N, F) |  |  |  |
| --- | --- | --- | --- | --- | --- | --- | --- | --- | --- | --- | --- | --- | --- | --- |
|  |  | Cases (N) | Controls (N) | Total (N) | Mean | SD | Range | Mean | SD | Range | Total (N) | Total (F) | Cases (F) | Controls (F) |
| IPDGC DUTCH | EUR | 61 | 1969 | 2030 | 53.3 | 5.5 | 45–75 | 34.1 | 4.7 | 18–39 | 1114 | 0.55 | 0.31 | 0.56 |
| IPDGC GERMANY | EUR | 63 | 935 | 998 | 47.4 | 12.4 | 19–69 | 35.6 | 3.2 | 28–39 | 465 | 0.47 | 0.32 | 0.48 |
| IPDGC NEUROX | EUR | 293 | 5075 | 5368 | 61.5 | 13.8 | 19–79 | 33.7 | 4.7 | 19–39 | 2393 | 0.45 | 0.34 | 0.45 |
| IPDGC NIA | EUR | 78 | 2937 | 3015 | 62.8 | 9.7 | 19–79 | 33.2 | 5.2 | 18–39 | 1576 | 0.52 | 0.42 | 0.53 |
| IPDGC OSLO | EUR | 44 | 433 | 477 | 60.4 | 9.9 | 38–79 | 35.1 | 3.5 | 24–39 | 191 | 0.40 | 0.18 | 0.42 |
| IPDGC SPAIN | EUR | 215 | 2579 | 2794 | 58.5 | 15.4 | 18–79 | 33.4 | 5.0 | 18–39 | 1476 | 0.53 | 0.37 | 0.54 |
| IPDGC TUBI | EUR | 32 | 498 | 530 | 66.4 | 7.7 | 21–79 | 35.1 | 4.1 | 23–39 | 305 | 0.58 | 0.25 | 0.60 |
| GP2 | EUR | 608 | 4635 | 5243 | 57.3 | 14.0 | 18–79 | 33.7 | 4.9 | 18–39 | 2745 | 0.52 | 0.40 | 0.54 |
| NGRC HAMZA | EUR | 134 | 1347 | 1481 | 62.9 | 10.8 | 21–79 | 34.4 | 5.0 | 18–39 | 868 | 0.59 | 0.29 | 0.62 |
| SUM | EUR | 1528 | 20408 | 21936 |  |  |  |  |  |  | 11133 | 0.51 |  |  |

<sup>a</sup> Age at sampling for controls if available. The variable was missing for 8.3% of the controls, all within the GP2-cohort (N=1687).
Abbreviations: Y = Years, AAO = Age at onset, YOPD = Young onset Parkinson's disease. N = Number, F = Frequency

**Table 2.**
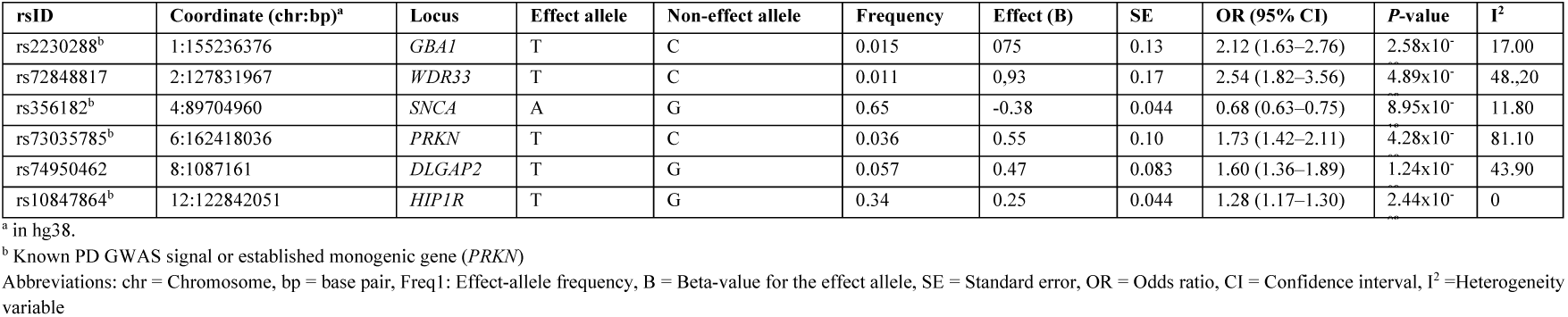
Genome-wide significant loci from the additive meta-analysis.

### Comparison with general Parkinson’s disease GWAS

Of the previously described 134 risk loci and 157 independent significant signals in the 2025 European GWAS^20^, lead SNPs corresponding to 119 risk loci and 138 of the independent signals were present in our meta-analysis summary statistics. We replicated 52 variants in 46 various loci, defined as similar direction of effect and *P* < 0.05, with 130 out of 138 SNPs having the same direction for the effect estimates (Supplementary Table 2). We note, however, that there is some sample overlap between the Leonard and GP2 2025 EUR summary statistics and our meta-analysis. Next, we compared the 90 risk signals reported by Nalls et al 2019^50^ with our analysis and found that 28 out of 88 SNPs present in our results were replicated at *P* < 0.05 (Supplementary Table 2). To further compare the results of our YOPD-specific GWAS with those of previous general European PD GWAS, we generated beta-beta plots of all independently significant SNPs from these studies, showing a strong correlation (r^2^ = 0.89, 95% CI 0.85-0.92 for GP2 vs YOPD; r^2^ = 0.91, 95% CI 0.86-0.94 for Nalls et al. vs YOPD, with only a small number of SNPs displaying a discordant direction of effect (Fig. 3). The two novel loci from our YOPD meta-GWAS showed only weak trends with the same direction of effect in GP2 and Nalls et al. summary statistics, without passing a nominal significance threshold of *P* < 0.05 (Supplementary Table 2), yet we note that this is also the case for the *PRKN* signal. A recent GWAS of PD with age at onset ≤ 50 from India^23^ nominated a novel SNP near *CCDC85A* (rs59330234), which showed no evidence of association with YOPD in our analysis (*P* = 0.58).

**Figure 3.**
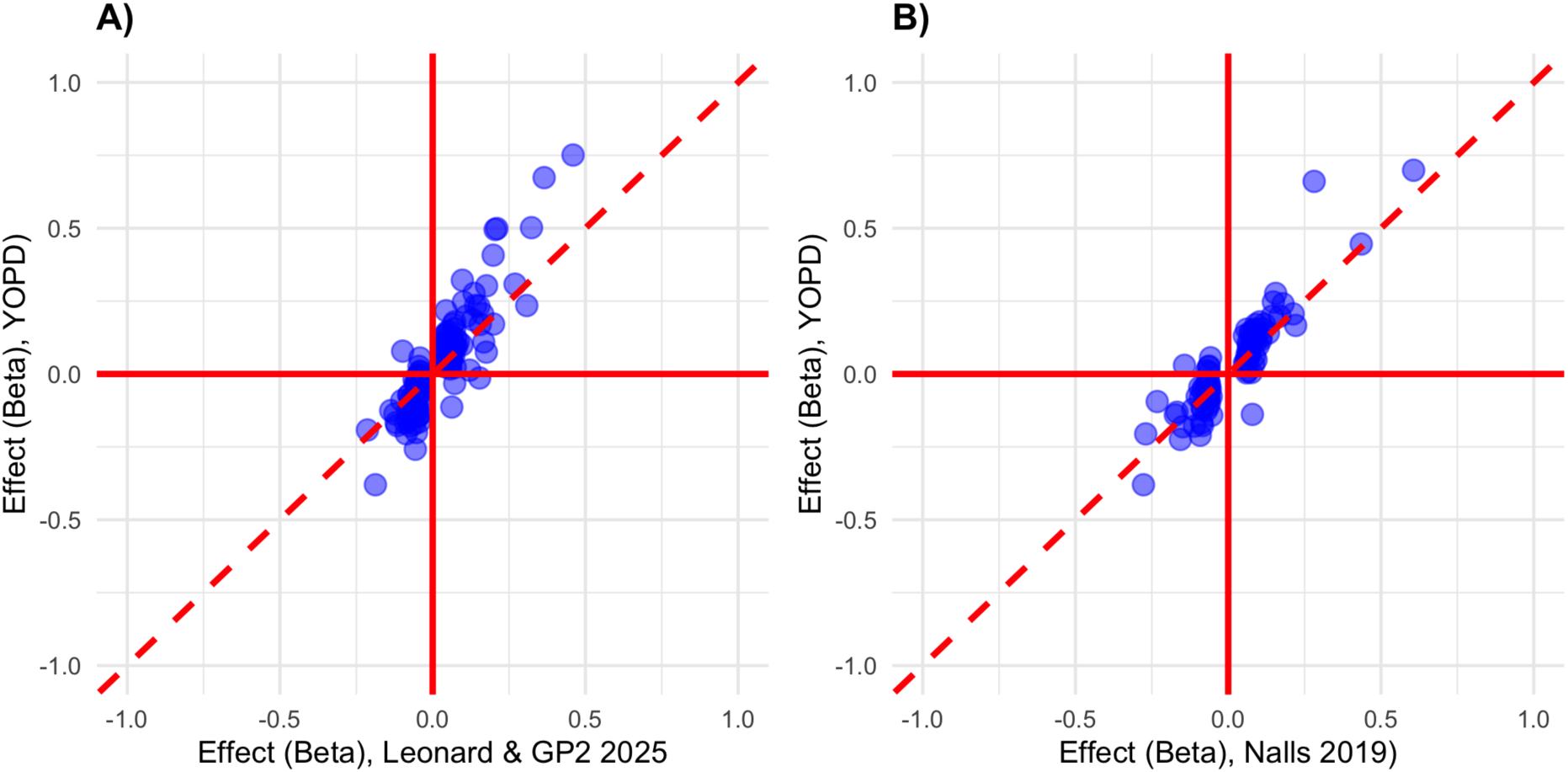
Beta-beta plots comparing effect sizes for the YOPD GWAS with previous PD GWASs. The plots include all genome-wide significant SNPs from the A) Leonard and GP2 2025 and B) Nalls et al. 2019 PD GWAS. The beta coefficients from the original general PD GWASs are plotted against the corresponding coefficients for the same SNPs derived from the present YOPD GWAS study.

### Stepwise conditional analysis of the *SNCA* locus

The *SNCA* region was the only locus where FUMA indicated the potential presence of multiple independent association signals. To further dissect *SNCA*-related risk in YOPD, we performed stepwise conditional analysis on individual level data from our GP2 cohort using PLINK 2.0^37^ focusing on the region ± 500kb from the lead SNP rs356182. We filtered out SNPs with minor allele frequency < 0.01 and included genetically determined sex and PC1–5 as covariates. The top SNP after conditioning on rs356182 was rs2869998 (*P* = 2.5×10^-5^, OR 1.38 (95% CI 1.19-1.60)). This signal is located further 3’, downstream of rs356182, and is in high linkage disequilibrium with a previously published independent *SNCA* signal, emerging as a tertiary signal in general PD.^35^ In further stepwise conditional analyses, two more SNPs passed the significance threshold, which were both low frequency variants located on the 3’ side of *SNCA* (Supplementary Table 3). The secondary GWAS signal from the 5’ end of the gene which has previously been highlighted both as a secondary signal in general PD^35,51,52^, and a top signal in dementia with Lewy bodies^53,54^ and REM sleep behaviour disorder^55^, reached nominal significance when conditioned on rs356182 and rs2869998 (Candidate analysis of rs7681154^51^: *P* = 0.042, OR = 0.87 (0.76-1.00), Supplementary Table 3).

### Recessive model genome-wide association analyses

The significant signal from the *PRKN* locus in the additive model GWAS prompted us to extend our analyses to a recessive model GWAS. A total of 6,875,354 SNPs were meta-analysed in the recessive model GWAS. We observed a lambda_1000_ of 1.17, indicating potential genomic inflation. However, lambda_1000_ was not increased above 1 in any individual dataset and did not decrease readily by applying SNP filtering based on missingness, minor allele frequency or heterogeneity across cohorts (Supplementary Table 4). The recessive meta-analysis identified nine loci passing the genome-wide significance threshold (Table 3, Fig. 1). Significant signals from the *SNCA* (rs356219, rs10008290) and *PRKN* (rs17529104) loci mirrored the additive model findings. Another signal was identified near *AKT3* (rs12139868, *P* = 2.27×10^8^), which also emerged as a significant locus in the recent European GP2 PD GWAS.^20^ In the case of *PRKN*, a known recessive PD locus, we observed a striking difference in effect size comparing the additive (OR = 1.73, CI 1.42-2.11) and recessive (OR = 23.5, CI 9.07-60.7) models. We compared odds ratios across models and assessed enrichment of homozygous genotypes in YOPD patients for the novel recessive signals. We found a similar pattern for a subset of these, albeit less extreme than for the *PRKN* locus (Supplementary Table 5).

**Table 3.**
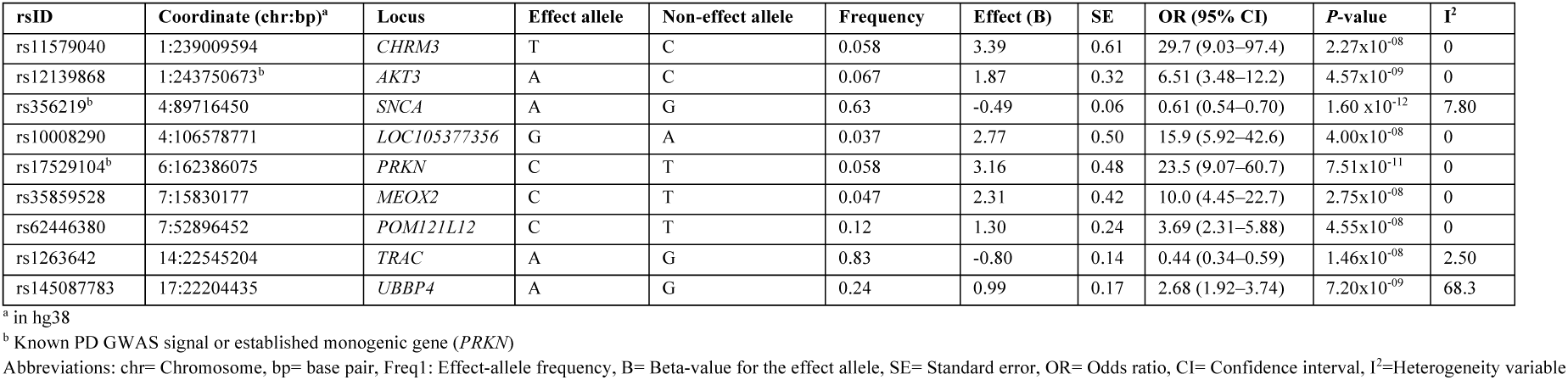
Genome-wide significant loci from the recessive meta-analysis.

### Polygenic risk score analysis

Separate PRS analyses were conducted using weights derived either from independent YOPD-GWAS summary statistics or from the general PD GWAS by Nalls et al.^50^ Comparisons across age at onset groups showed that the YOPD-PRS was significantly higher in YOPD compared to both other groups whereas the general PD-PRS showed a significant difference only when comparing age at onset 40–59 to 60–80 (Table 4). Based on this indication that the YOPD-PRS captures aspects of genetic risk that are specific to YOPD, we went on to investigate whether this risk is driven by one or more specific molecular pathways. Out of the six selected pathways we investigated, only the mitochondrial PRS was significantly higher in YOPD than in the other groups (*P* = 0.0060 vs 40-59 and *P* = 0.0040 vs 60-80) (Fig. 4).

**Figure 4.**
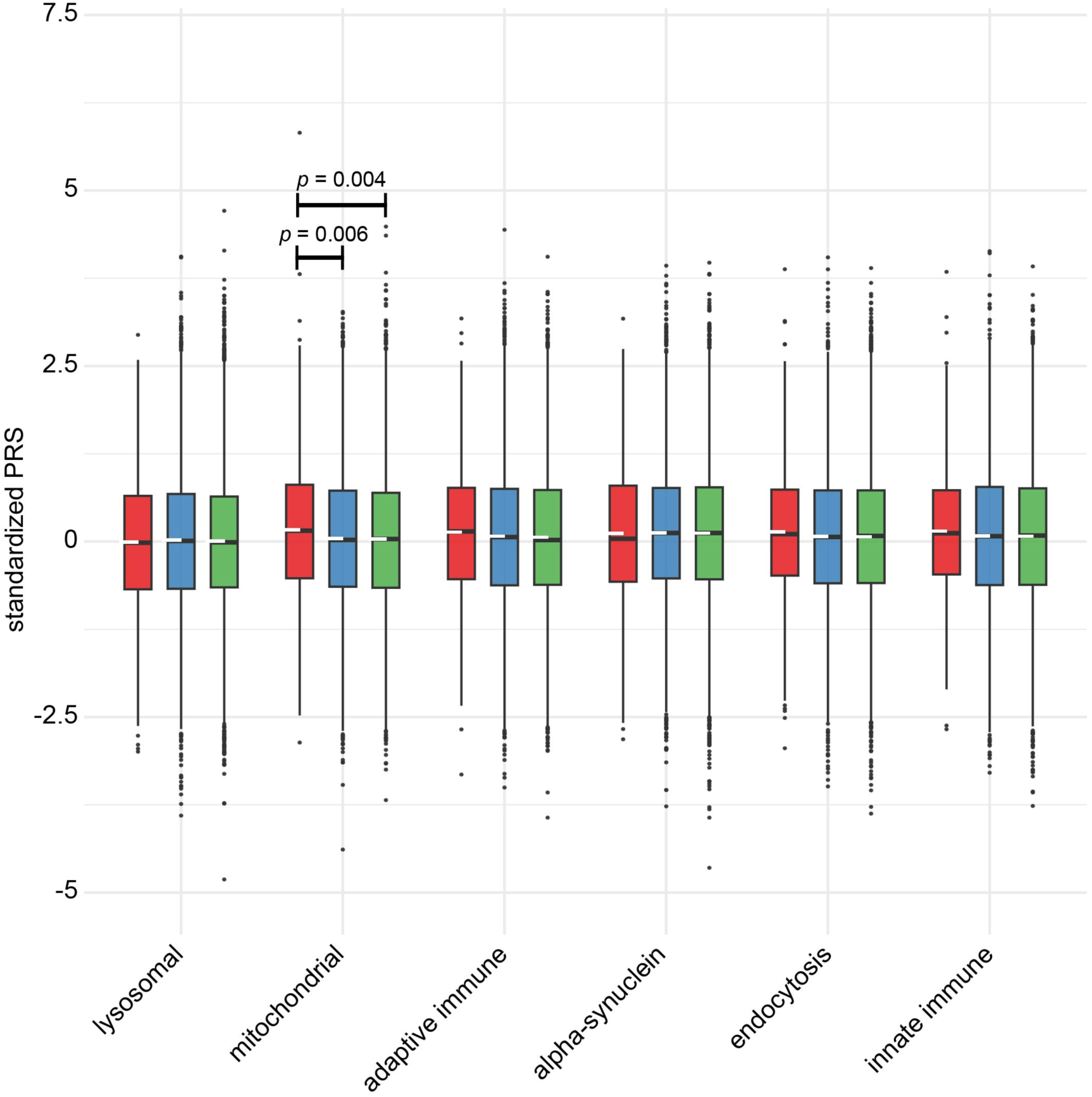
Pathway-specific YOPD-PRS across age at onset groups. Boxplots of pathway-specific polygenic risk cores in age at onset groups 18-39 (red, left), 40-59 (blue, middle) and 60-80 (green, right). Note that the mitochondrial PRS is significantly higher in the young onset group, compared to both other age at onset groups.

**Table 4.** Comparison of Parkinson’s disease polygenic risk scores across age at onset groups.

| Weights | Age at onset groups (years) |  |  | Groupwise comparison T-tests |  |  |
| --- | --- | --- | --- | --- | --- | --- |
|  | A: 18 – 39 | B: 40 – 59 | C: 60 – 80 | P-value A vs B | P-value A vs C | P-value B vs C |
| YOPD-GWAS | 0.31 (0.23-0.40) | 0.16 (0.14-0.19) | 0.16 (0.13-0.18) | 0.0012 | 0.00059 | 0.72 |
| General PD-GWAS | 0.26 (0.18-0.34) | 0.29 (0.26-0.32) | 0.20 (0.17-0.22) | 0.53 | 0.13 | 4.7x10<sup>-7</sup> |

Abbreviations: YOPD = Young onset Parkinson's disease, GWAS = Genome wide association study, PD = Parkinson's disease, Y = Years, OR = Odds ratio, CI = Confidence interval
Parkinson's disease polygenic risk scores among different age at onset groups, modeled on either YOPD-GWAS specific or General PD-GWAS weights (Nalls et al 2019<sup>19</sup> without 23andMe data). Values correspond to mean (95% confidence interval).

### Runs of homozygosity

A total of 146 cases and 223 controls in the GP2 dataset had available WGS data. Across 5 out of 8 loci from the recessive GWAS (excluding *SNCA*), a varying number of homozygous carriers of lead variants were observed in both cases and controls with available WGS. We did not observe any individual homozygous for a recessive GWAS effect allele also harbouring an ROH overlapping the GWAS variant position. Overall, no statistically significant differences in ROH presence at recessive GWAS loci were detected between cases and controls in the WGS subset, which may reflect the limited number of samples with WGS data and, consequently, reduced statistical power.

### Analysis of homozygous deleterious variants from WGS data

Annotation and filtering of WGS variants within flanking regions of recessive model GWAS signals identified six rare variants meeting criteria for potential recessive deleteriousness and present as homozygotes in cases but not controls. However, only one variant was observed in an individual who was also homozygous for the corresponding nearby recessive GWAS lead variant effect allele, namely rs12699752 within 9.8kb of the GWAS lead SNP rs35859528. This SNP was flagged because of its CADD score of 20.1 and it sits inside an ENCODE4 annotated distal enhancer element. The carrying individual was not found to have a ≥ 250kb ROH at this site, but the distance from the lead SNPs was considerably smaller than this minimum threshold (9.8kb).

## Discussion

We have conducted a GWAS meta-analysis of YOPD in cohorts of European ancestry. Unsurprisingly, the results confirm that the genetic risk factors of sporadic YOPD are largely overlapping with those of PD in general, which is in line with previous observations of increasing PD PRS in young onset patients compared to later onset PD.^21,22^ However, we also present a number of findings indicating that the genetic architecture of YOPD includes distinctive elements, which are specific to the young onset phenotype. The additive model GWAS analysis identified two novel signals from loci not previously associated with PD. The first of these, rs74950462, is located within an intron of *DLGAP2*, encoding DLG associated protein 2, a membrane-bound protein of the postsynaptic density, involved in synapse organization and neuronal signaling.^56^ Previous work has demonstrated downregulation of *DLGAP2* in an induced pluripotent stem cell model of familial PD^57^ and rare mutations in this gene have been implicated in autism spectrum disorder.^58^ The lead SNP of the second novel signal, rs72848817, is located in the intergenic region between *WDR33* and *POL2RD*. Both novel signals had lead SNPs of relatively low frequency, which were not present in all the meta-analysed datasets, and the associations showed moderate heterogeneity (Table 2, Fig. 2). These results should therefore be interpreted with caution and corroboration of these nominated loci will require further replication in independent datasets.

The results from PRS analyses provide further evidence that the genetic architecture of YOPD is partly distinct, even in sporadic disease. Firstly, PRS estimates in GP2 participants differentiated significantly between YOPD and groups with later-onset PD when independent YOPD-specific summary statistics were used as weights, but not when standard PD summary statistics from a large previous GWAS study were used. Next, pathway-specific YOPD-PRS analyses showed that this effect was primarily driven by genetic risk annotated to the mitochondrial pathway. We find this result both intriguing and biologically plausible, given the fact that mitochondrial dysfunction is the primary molecular mechanism most strongly implicated in autosomal recessive PD, typically causing a young onset phenotype.^59^ Our findings suggest that mitochondrial genes are disproportionately important in young onset patients, not only in monogenic PD, but also in sporadic disease.

Effect estimates in YOPD showed a strong correlation with results from general PD GWAS and as in these studies, the strongest of all association signals was rs356182, near the 3’ end of *SNCA*. Compared to published PD studies, the two independent 3’ *SNCA* association signals showed stronger effect sizes in YOPD, whereas the 5’ *SNCA* signal previously reported in PD, DLB, and RBD^35,51–55^ was only marginally significant, with a smaller effect size than reported in previous conditional analyses of PD with any age at onset.^35,51,52^ This observation was based on a limited sample size, but could potentially fit well with established clinical evidence indicating lower rates of both dementia and RBD in YOPD patients compared to later onset PD.^60^

The additive model GWAS analysis identified a significant signal near *PRKN*, the most common gene causing autosomal recessive PD.^6,10^ A similar observation has been reported previously in a PD GWAS from Spain, including samples that overlap with the present study, where the lead SNP was shown to be in high LD with a coding *PRKN* mutation specific to the Spanish population (c.155delA).^49^ The same signal is driving the association observed in our analyses, where the *PRKN* signal is almost entirely driven by the Spanish dataset (Supplementary Figure 3). While not novel, this *PRKN* finding provides proof of principle that low frequency variants in LD with highly penetrant recessive mutations may indeed be detectable by GWAS methodology, and it prompted us to extend our GWAS analysis by also applying a recessive model. Interestingly, the *PRKN* signal showed a striking increase in both the level of significance and the effect estimate in the recessive model (Table 2 and Table 3), further justifying this approach.

The recessive model GWAS meta-analysis identified seven significant loci besides *SNCA* and *PRKN*, a larger number of signals compared to the additive model. In the absence of true recessive effects, a recessive model should be expected to have lower statistical power. Higher λ_1000_ value in the recessive model GWAS indicates inflated *P*-values, which could potentially be the result of more vulnerable test statistics as groups of homozygous low frequency SNP carriers become very small. Filtering SNPs based on allele frequency, heterogeneity, or missingness did not readily remove all inflation, however. An alternative explanation could be a true enrichment of low *P*-values driven by recessive effects linked to excess homozygosity in YOPD, which has been reported in several previous publications.^15,16^ Follow-up analysis in WGS data from a minor subset of GP2 YOPD subjects did not identify deleterious variants in homozygous state shared by multiple patients, so we were not able to nominate potential strong causal drivers of the recessive signals. We note that it is not necessarily biologically implausible *a priori* that genetic risk factors with lower penetrance may act in a recessive manner, although this type of effect has been less studied in GWAS to date. Further studies exploring potential recessive effects may also be warranted in larger PD datasets of any age at onset.

We acknowledge that this study has several limitations. Importantly, 1,528 patients is a small sample size for a GWAS, and the study lacked power for a range of relevant post-GWAS analyses.^39–41,61,62^ Nominated novel loci should be regarded as preliminary pending independent replication. Results from the recessive model analysis should be interpreted with particular caution, given the low number of homozygous carriers on which these findings are based. Several of the novel significant signals we report are low frequency variants that were not present in all included datasets. The *PRKN* signal illustrates that even within subjects of European ancestry, rare or low frequency alleles may be population-specific, increasing the risk of both type I and type II errors in a relatively small GWAS meta-analysis. Moreover, genotyping arrays used for different cohorts in this study had variable density, which may impact imputation accuracy and contribute to incomplete data on low-frequency variants. Due to limited availability of data from YOPD subjects, our analysis included only subjects of European ancestry, and further studies are needed to elucidate the genetic architecture of YOPD across a broader range of diverse populations.

An International Parkinson and Movement Disorder Society Task Force on Early Onset Parkinson’s Disease recently recommended the term early-onset PD (EOPD), rather than YOPD, with an age at onset cutoff of 50 years.^63^ The task force highlighted that the literature has seen a shift towards using EOPD as a potentially less age-stigmatizing term and pointed to changing oestrogen levels in women around menopause as an argument for using 50 years as a cutoff. While we acknowledge these important arguments and agree that there are contexts where a cutoff at 50 years would be more appropriate, we considered onset below 40 years to be most relevant for our particular research question, hypothesizing a partly distinct genetic architecture. We therefore deliberately used the term YOPD for this cutoff, as coined in the original article by Quinn, Crithcley and Marsden^4^, reserving the term EOPD for the group with onset before the age of 50 years.

In conclusion, we conducted a GWAS meta-analysis of YOPD in European ancestry subjects, showing substantial overlap between YOPD and sporadic PD in general, but also providing evidence for a partly distinct genetic architecture of YOPD. We nominate novel YOPD-specific risk loci and highlight the mitochondrial pathway as a significant driver of polygenic YOPD risk. Further research is needed to corroborate these findings and expand the exploration of YOPD genetics to other ancestries. Recognizing that PD patients with disease onset in their 20ies or 30ies may differ from typical late onset patients genetically, even in the absence of a monogenic cause, resonates with clinical experience of a particular symptom profile, and may have important implications for future therapeutic research and clinical trial design.

## Supporting information

GP2 members

## Data availability

Data used in the preparation of this article were obtained from the Global Parkinson’s Genetics Program (GP2; https://gp2.org). Specifically, we used Tier 2 data from GP2 release 10 (https://doi.org/10.5281/zenodo.7904831). Data from GP2 are hosted in collaboration with the Accelerating Medicines Partnership in Parkinson’s disease and are available via an application on the website (https://amp-pd.org/register-for-amp-pd). Summary statistics from Nalls et al 2019 (without 23andMe data)^19^ and Leonard and GP2 2025^20^ were accessed through the GP2 platform. Data from NGRC are available from the database of Genotypes and Phenotypes (dbGaP, https://dbgap.ncbi.nlm.nih.gov/), study accession phs000196.v2.p1. Data from two IPDGC subcohorts are available in dbGaP (study accessions phs000918.v1.p1, phs000089.v3.p2). For other IPDGC subcohorts, raw data have not been made public due to restrictions relating to privacy regulations, informed consent or ethical approvals.

## Code availability

Summary statistics and code for all analyses, including specifications of software versions, are publicly available on GitHub at https://github.com/lpihlstrom/projects. This repository was assigned a persistent identifier via Zenodo at https://doi.org/10.5281/zenodo.20476724.

## Acknowledgements

The authors would like to thank all the participants who contributed to the studies that this work was based on. Data and/or code used in the preparation of this article were obtained from Global Parkinson’s Genetics Program (GP2; https://gp2.org). GP2 is funded by the Aligning Science Across Parkinson’s (ASAP) (https://ror.org/03zj4c476) initiative and implemented by The Michael J. Fox Foundation for Parkinson’s Research (https://ror.org/03arq3225). For a complete list of GP2 members and organizations seehttps://gp2.org. We would like to thank all members of GP2, the International Parkinson’s Disease Genomics Consortium, the NeuroGenetics Research Consortium^64–68^ and the Funding source for the NGRC study, (5R01NS36960-10), The National Institutes of Health, Bethesda, MD, USA. GPT UiO – The University of Oslo’s privacy friendly GPT chat (https://www.uio.no/english/services/it/ai/gpt-uio/index.html) and OpenAi’s Chat GPT were used in code editing.

## Funding

IHL, MV and LP were funded by grants from the South-Eastern Regional Health Authority, Norway. KS is supported by The Michael J. Fox Foundation and Aligning Sciences Across Parkinson’s Disease Global Parkinson Genetic Program. JVW was supported by travel grants from Alzheimer Nederland and FENS-IBRO.

## Competing interests

The authors report no competing interests.

## Notes

### Competing Interest Statement

The authors have declared no competing interest.

### Author Declarations

Regional committees for medical and health research ethics South-East Norway gave ethical approval for this work.

