## Supplementary for "A genome-wide association study of young onset Parkinson’s disease in European ancestry"

**Suppmentary Materials to:**

### Supplementary Table 1: Overview of genotyping arrays used

| Dataset | Ancestry | Array type |
|---|---|---|
| IPDGC DUTCH | EUR | Illumina Human660W-Quad beadchips, Illumina Human610K beadchips |
| IPDGC GERMANY | EUR | Illumina HumanHap550, Illumina human Omni express |
| IPDGC NEUROX DBGAP | EUR | Illumina NeuroX |
| IPDGC NIA | EUR | Illumina HumanHap550 |
| IPDGC OSLO | EUR | Illumina Human OmniExpress |
| IPDGC SPAIN | EUR | NeuroChip |
| IPDGC TUBI | EUR | NeuroChip |
| GP2 | EUR | Illumina NeuroBooster Array |
| NGRC HAMZA | EUR | Illumina HumanOmni1-Quad_v1-0_B |

Information on genotyping was retreived from the original publications[1-6] and the supplementary table S1 from Leonard and GP2 2025 GWAS.[7]

### Supplementary Table 2 Comparison of lead SNP association statistics across the present study, Nalls et al. 2019, and Leonard and GP2 2025

**Significant loci from the additive YOPD GWAS compared to Leonard and GP2 2025 GWAS sumstats and Nalls et al 2019 GWAS sumstats**

| rsID | CHR | BP | Nearest gene | Effect allele | Other allele | Freq YOPD | Effect YOPD | StdErr YOPD | P YOPD | Freq GP2 | Beta GP2 | SE GP2 | P GP2 | Freq Nalls | Effect Nalls | StdErr Nalls | P Nalls |
|---|---|---|---|---|---|---|---|---|---|---|---|---|---|---|---|---|---|
| rs2230288 | 1 | 155236376 | *GBA* | T | C | 0.01 | 0.75 | 0.13 | 2.6E-08 | 0.02 | 0.46 | 0.02 | 1.0E-78 | 0.02 | 0.73 | 0.07 | 5.5E-27 |
| rs72848817 | 2 | 127831967 | *WDR33* | T | C | 0.01 | 0.93 | 0.17 | 4.9E-08 | 0.01 | -0.02 | 0.04 | 0.69 | | | | |
| rs356182 | 4 | 89704960 | *SNCA* | A | G | 0.65 | -0.38 | 0.04 | 9.0E-18 | 0.64 | -0.19 | 0.01 | 7.5E-158 | 0.62 | -0.26 | 0.02 | 4.9E-34 |
| rs73035785 | 6 | 162418036 | *PRKN* | T | C | 0.04 | 0.55 | 0.10 | 4.3E-08 | 0.04 | 0.03 | 0.02 | 0.061 | 0.04 | 0.00 | 0.06 | 0.95 |
| rs74950462 | 8 | 1087161 | *DLGAP2* | T | G | 0.06 | 0.47 | 0.08 | 1.2E-08 | 0.06 | -0.01 | 0.01 | 0.63 | 0.05 | 0.06 | 0.05 | 0.29 |
| rs10847864 | 12 | 122842051 | *HIP1R* | T | G | 0.34 | 0.25 | 0.04 | 2.4E-08 | 0.35 | 0.10 | 0.01 | 4.2E-40 | 0.36 | 0.13 | 0.02 | 1.3E-12 |

**Significant signals from Leonard 2025 GWAS compared to YOPD additive GWAS summary statistics (138 overlapping SNPs)**

| rsID | CHR | BP | Nearest Gene | Effect allele | Ref allele | Freq GP2 | Beta GP2 | SE GP2 | P GP2 | Freq YOPD | Beta YOPD | SE YOPD | P YOPD | Effect direction concordant |
|---|---|---|---|---|---|---|---|---|---|---|---|---|---|---|
| rs377808 | 1 | 52724687 | *ZYG11B* | G | T | 0.50 | -0.05 | 0.008 | 1.02E-10 | 0.50 | -0.15 | 0.064 | 0.02332 | TRUE |
| rs12128848 | 1 | 112106855 | *KCND3* | G | A | 0.29 | -0.04 | 0.007 | 3.15E-08 | 0.29 | -0.02 | 0.048 | 0.6976 | TRUE |
| rs75548401 | 1 | 155236246 | *GBA1* | A | G | 0.01 | 0.36 | 0.034 | 1.36E-27 | 0.01 | 0.67 | 0.184 | 0.0002554 | TRUE |
| rs2230288 | 1 | 155236376 | *GBA1* | T | C | 0.02 | 0.46 | 0.024 | 1.05E-78 | 0.01 | 0.75 | 0.135 | 2.577E-08 | TRUE |
| rs72712862 | 1 | 161000299 | *F11R* | T | C | 0.01 | 0.21 | 0.034 | 4.28E-10 | 0.01 | 0.50 | 0.277 | 0.07121 | TRUE |
| rs72712893 | 1 | 161067049 | *ARHGAP30* | G | C | 0.01 | 0.20 | 0.033 | 4.66E-10 | 0.01 | 0.50 | 0.274 | 0.07044 | TRUE |
| rs72714968 | 1 | 161327996 | *SDHC* | G | A | 0.02 | 0.15 | 0.022 | 7.16E-12 | 0.02 | -0.01 | 0.187 | 0.9412 | FALSE |
| rs61802091 | 1 | 161419856 | *SDHC* | A | G | 0.13 | -0.07 | 0.010 | 1.1E-10 | 0.13 | -0.10 | 0.070 | 0.1608 | TRUE |
| rs708723 | 1 | 205770138 | *RAB29* | T | C | 0.55 | 0.08 | 0.007 | 1.5E-28 | 0.56 | 0.12 | 0.040 | 0.002104 | TRUE |
| rs10495249 | 1 | 226731418 | *ITPKB* | G | A | 0.28 | -0.06 | 0.007 | 1.4E-14 | 0.29 | -0.12 | 0.044 | 0.006092 | TRUE |

| | | | | | | | | | | | | | | |
|---|---|---|---|---|---|---|---|---|---|---|---|---|---|---|
| rs12073680 | 1 | 232508908 | *SIPA1L2* | G | A | 0.13 | 0.07 | 0.010 | 6.76E-11 | 0.13 | 0.17 | 0.055 | 0.001986 | TRUE |
| rs7517340 | 1 | 243546888 | *AKT3* | C | T | 0.81 | -0.05 | 0.009 | 5.09E-09 | 0.81 | -0.03 | 0.057 | 0.5704 | TRUE |
| rs76116224 | 2 | 17966582 | *KCNS3* | T | A | 0.10 | -0.07 | 0.012 | 6.13E-10 | 0.10 | -0.18 | 0.080 | 0.02563 | TRUE |
| rs12478701 | 2 | 23718478 | *KLHL29* | C | T | 0.49 | 0.04 | 0.007 | 8.32E-09 | 0.49 | 0.13 | 0.043 | 0.002359 | TRUE |
| rs6749019 | 2 | 31572741 | *SRD5A2* | A | C | 0.54 | 0.04 | 0.007 | 4.83E-09 | 0.54 | 0.04 | 0.044 | 0.4065 | TRUE |
| rs13010404 | 2 | 32623787 | *TTC27* | G | T | 0.34 | -0.05 | 0.008 | 5.29E-12 | 0.35 | -0.20 | 0.069 | 0.003507 | TRUE |
| rs9309337 | 2 | 61536072 | *XPO1* | C | T | 0.76 | -0.05 | 0.008 | 7.58E-11 | 0.76 | -0.13 | 0.051 | 0.0112 | TRUE |
| rs9309428 | 2 | 69465874 | *AAK1* | T | C | 0.44 | 0.05 | 0.007 | 3.21E-13 | 0.44 | 0.06 | 0.040 | 0.1567 | TRUE |
| rs7563891 | 2 | 95326214 | *KCNIP3* | T | C | 0.64 | 0.05 | 0.007 | 3.56E-10 | 0.67 | 0.06 | 0.047 | 0.2166 | TRUE |
| rs60954733 | 2 | 101170053 | *TBC1D8* | G | A | 0.24 | -0.05 | 0.008 | 8.05E-09 | 0.24 | -0.10 | 0.052 | 0.05782 | TRUE |
| rs6758044 | 2 | 134834675 | *ACMSD* | C | T | 0.41 | -0.10 | 0.007 | 1.25E-44 | 0.42 | -0.09 | 0.061 | 0.1355 | TRUE |
| rs3754541 | 2 | 147901026 | *ACVR2A* | A | G | 0.20 | -0.05 | 0.009 | 9.11E-10 | 0.20 | -0.10 | 0.081 | 0.2056 | TRUE |
| rs10173157 | 2 | 157798690 | *ACVR1* | G | A | 0.80 | -0.05 | 0.009 | 8.14E-09 | 0.81 | -0.12 | 0.055 | 0.02842 | TRUE |
| rs1710669 | 2 | 160967491 | *TANK* | G | C | 0.75 | 0.05 | 0.008 | 4.29E-08 | 0.76 | 0.08 | 0.052 | 0.1132 | TRUE |
| rs12618884 | 2 | 168129365 | *STK39* | G | A | 0.47 | 0.04 | 0.007 | 4.67E-09 | 0.46 | 0.07 | 0.039 | 0.07694 | TRUE |
| rs2102808 | 2 | 168260515 | *STK39* | T | G | 0.13 | 0.15 | 0.010 | 4.76E-52 | 0.12 | 0.23 | 0.056 | 0.00002784 | TRUE |
| rs17046610 | 3 | 7024398 | *GRM7* | A | G | 0.04 | -0.12 | 0.016 | 1.08E-13 | 0.05 | -0.17 | 0.118 | 0.1574 | TRUE |
| rs144678625 | 3 | 17710565 | *TBC1D5* | G | A | 0.01 | 0.17 | 0.030 | 3.2E-08 | 0.01 | 0.11 | 0.197 | 0.5769 | TRUE |
| rs73034368 | 3 | 18247575 | *TBC1D5* | A | C | 0.04 | 0.13 | 0.017 | 5.37E-15 | 0.04 | 0.19 | 0.102 | 0.0673 | TRUE |
| rs1461806 | 3 | 28658687 | *RBMS3* | G | A | 0.61 | -0.05 | 0.007 | 4.17E-13 | 0.63 | -0.12 | 0.040 | 0.003213 | TRUE |
| rs1768208 | 3 | 39481512 | *MOBP* | C | T | 0.72 | -0.04 | 0.007 | 1.48E-08 | 0.72 | 0.01 | 0.044 | 0.8266 | FALSE |
| rs56384862 | 3 | 58410136 | *PXK* | G | A | 0.37 | 0.04 | 0.008 | 4.22E-08 | 0.38 | 0.08 | 0.040 | 0.03413 | TRUE |
| rs58088236 | 3 | 121842918 | *EAF2* | T | C | 0.38 | -0.04 | 0.007 | 7.33E-09 | 0.38 | -0.05 | 0.041 | 0.2538 | TRUE |
| rs11717169 | 3 | 151405709 | *MED12L* | C | T | 0.37 | -0.05 | 0.007 | 3.26E-10 | 0.38 | -0.05 | 0.040 | 0.1925 | TRUE |
| rs359545 | 3 | 155551064 | *PLCH1* | G | A | 0.84 | 0.06 | 0.009 | 6.42E-10 | 0.85 | 0.02 | 0.062 | 0.781 | TRUE |
| rs11929238 | 3 | 161328149 | *SPTSSB* | T | A | 0.32 | 0.06 | 0.008 | 7.37E-13 | 0.31 | 0.05 | 0.042 | 0.2117 | TRUE |
| rs3729674 | 3 | 179199217 | *PIK3CA* | G | A | 0.19 | 0.06 | 0.009 | 3.83E-13 | 0.20 | 0.12 | 0.053 | 0.02375 | TRUE |
| rs34822404 | 3 | 183028420 | *MCCC1* | T | C | 0.19 | -0.12 | 0.009 | 2.13E-41 | 0.20 | -0.18 | 0.052 | 0.0006942 | TRUE |
| rs77186370 | 4 | 940049 | *TMEM175* | A | C | 0.02 | 0.12 | 0.022 | 3.28E-08 | 0.02 | 0.01 | 0.133 | 0.9208 | TRUE |
| rs34884217 | 4 | 950422 | *TMEM175* | C | A | 0.11 | -0.12 | 0.011 | 4.07E-27 | 0.11 | -0.14 | 0.067 | 0.03961 | TRUE |
| rs34311866 | 4 | 958159 | *TMEM175* | C | T | 0.19 | 0.16 | 0.008 | 2.84E-85 | 0.18 | 0.21 | 0.048 | 0.00001795 | TRUE |
| rs2857866 | 4 | 2934914 | *MFSD10* | G | T | 0.81 | 0.07 | 0.009 | 3.78E-16 | 0.82 | -0.03 | 0.051 | 0.5247 | FALSE |
| rs4698412 | 4 | 15735725 | *BST1* | A | G | 0.55 | 0.07 | 0.007 | 1.24E-26 | 0.54 | 0.18 | 0.039 | 4.564E-06 | TRUE |
| rs13117238 | 4 | 76252761 | *FAM47E-STBD1* | A | C | 0.16 | 0.07 | 0.009 | 5.18E-16 | 0.16 | 0.09 | 0.052 | 0.07466 | TRUE |
| rs28628748 | 4 | 76278760 | *FAM47E-STBD1* | A | G | 0.37 | -0.07 | 0.007 | 2.64E-24 | 0.36 | -0.08 | 0.041 | 0.04226 | TRUE |
| rs356182 | 4 | 89704960 | *SNCA* | A | G | 0.64 | -0.19 | 0.007 | 7.47E-158 | 0.65 | -0.38 | 0.044 | 8.951E-18 | TRUE |
| rs2301134 | 4 | 89837794 | *SNCA* | G | A | 0.50 | 0.04 | 0.007 | 1.3E-09 | 0.51 | 0.05 | 0.039 | 0.2485 | TRUE |
| rs13117519 | 4 | 113447909 | *CAMK2D* | T | C | 0.17 | 0.07 | 0.009 | 1.74E-15 | 0.17 | 0.15 | 0.052 | 0.002855 | TRUE |
| rs72696649 | 4 | 169711869 | *CLCN3* | A | T | 0.35 | -0.05 | 0.007 | 3.94E-13 | 0.36 | -0.06 | 0.041 | 0.1348 | TRUE |
| rs75739600 | 4 | 169996341 | *MFAP3L* | C | A | 0.08 | 0.07 | 0.013 | 2.08E-08 | 0.08 | 0.02 | 0.083 | 0.7826 | TRUE |
| rs461753 | 5 | 1431099 | *SLC6A3* | T | C | 0.22 | -0.04 | 0.008 | 3.51E-08 | 0.22 | 0.00 | 0.054 | 0.96 | TRUE |
| rs1867598 | 5 | 60842132 | *ELOVL7* | G | A | 0.09 | 0.14 | 0.011 | 6.74E-33 | 0.10 | 0.28 | 0.061 | 5.428E-06 | TRUE |
| rs246814 | 5 | 76303383 | *SV2C* | T | C | 0.08 | 0.08 | 0.012 | 4.21E-12 | 0.08 | 0.11 | 0.069 | 0.1071 | TRUE |
| rs10054911 | 5 | 80530472 | *FAM151B* | A | T | 0.05 | -0.09 | 0.015 | 8.62E-09 | 0.05 | -0.21 | 0.151 | 0.1755 | TRUE |
| rs11241774 | 5 | 124872055 | *ZNF608* | T | A | 0.52 | 0.04 | 0.007 | 2.26E-08 | 0.53 | 0.03 | 0.044 | 0.5598 | TRUE |

| | | | | | | | | | | | | | | |
|---|---|---|---|---|---|---|---|---|---|---|---|---|---|---|
| rs2350640 | 5 | 138443292 | *REEP2* | C | A | 0.55 | -0.04 | 0.007 | 5.12E-09 | 0.54 | -0.01 | 0.045 | 0.8926 | TRUE |
| rs7741241 | 6 | 13664623 | *RANBP9* | A | G | 0.37 | -0.04 | 0.007 | 8.32E-09 | 0.37 | -0.05 | 0.045 | 0.2481 | TRUE |
| rs4713072 | 6 | 27224312 | *POM121L2* | C | T | 0.26 | 0.05 | 0.008 | 1.17E-09 | 0.27 | 0.14 | 0.043 | 0.0009161 | TRUE |
| rs6913724 | 6 | 27287064 | *POM121L2* | T | A | 0.49 | 0.04 | 0.007 | 1.39E-10 | 0.49 | 0.10 | 0.040 | 0.009564 | TRUE |
| rs41316625 | 6 | 29015989 | *ZNF311* | A | G | 0.04 | -0.10 | 0.017 | 5E-09 | 0.04 | 0.08 | 0.112 | 0.4832 | FALSE |
| rs2647066 | 6 | 32603345 | *HLA-DRB1* | T | C | 0.17 | -0.14 | 0.009 | 1.13E-49 | 0.16 | -0.13 | 0.080 | 0.1164 | TRUE |
| rs12528068 | 6 | 71778059 | *RIMS1* | T | C | 0.28 | 0.06 | 0.007 | 8.51E-15 | 0.28 | 0.06 | 0.043 | 0.1952 | TRUE |
| rs13340407 | 6 | 111836412 | *FYN* | C | T | 0.14 | -0.06 | 0.010 | 2.93E-09 | 0.15 | -0.17 | 0.058 | 0.003229 | TRUE |
| rs146617529 | 6 | 132828914 | *RPS12* | G | A | 0.03 | 0.20 | 0.021 | 1.1E-20 | 0.03 | 0.17 | 0.174 | 0.3241 | TRUE |
| rs6461688 | 7 | 23076996 | *KLHL7* | A | G | 0.41 | -0.08 | 0.007 | 8.19E-29 | 0.42 | -0.07 | 0.045 | 0.1046 | TRUE |
| rs2127183 | 7 | 95711872 | *DYNC1I1* | C | A | 0.42 | -0.04 | 0.007 | 1.9E-09 | 0.42 | 0.01 | 0.044 | 0.8766 | FALSE |
| rs6979335 | 7 | 100492237 | *NYAP1* | C | T | 0.19 | -0.06 | 0.010 | 1.6E-10 | 0.19 | -0.02 | 0.082 | 0.8218 | TRUE |
| rs80148128 | 7 | 158605408 | *PTPRN2* | A | G | 0.50 | 0.04 | 0.007 | 3.21E-08 | 0.50 | 0.11 | 0.064 | 0.09116 | TRUE |
| rs10087654 | 8 | 1761106 | *CLN8* | A | G | 0.70 | 0.05 | 0.008 | 4.03E-09 | 0.71 | 0.11 | 0.072 | 0.1384 | TRUE |
| rs620490 | 8 | 16840070 | *FGF20* | G | T | 0.28 | -0.08 | 0.008 | 1.68E-27 | 0.28 | -0.13 | 0.044 | 0.00419 | TRUE |
| rs4872005 | 8 | 22672162 | *BIN3* | C | G | 0.31 | -0.04 | 0.007 | 7.86E-09 | 0.32 | -0.05 | 0.043 | 0.2041 | TRUE |
| rs6469273 | 8 | 109633256 | *SYBU* | G | C | 0.23 | 0.05 | 0.008 | 8.18E-11 | 0.23 | 0.04 | 0.050 | 0.408 | TRUE |
| rs2663613 | 8 | 129873161 | *CYRIB* | C | T | 0.48 | 0.04 | 0.007 | 6.65E-09 | 0.48 | 0.07 | 0.039 | 0.07382 | TRUE |
| rs8181077 | 9 | 17580618 | *SH3GL2* | T | A | 0.34 | -0.07 | 0.007 | 3.21E-25 | 0.34 | -0.12 | 0.042 | 0.003284 | TRUE |
| rs1536072 | 9 | 17701891 | *SH3GL2* | G | C | 0.25 | 0.08 | 0.008 | 5.76E-25 | 0.26 | 0.09 | 0.045 | 0.03454 | TRUE |
| rs13296299 | 9 | 34154849 | *UBAP1* | A | T | 0.27 | 0.05 | 0.008 | 7.1E-12 | 0.26 | 0.09 | 0.043 | 0.03633 | TRUE |
| rs10796307 | 10 | 15507549 | *ITGA8* | C | A | 0.71 | 0.05 | 0.008 | 9.22E-10 | 0.72 | 0.14 | 0.045 | 0.002556 | TRUE |
| rs2282295 | 10 | 102415138 | *PSD* | A | G | 0.11 | 0.06 | 0.011 | 1.79E-08 | 0.10 | -0.11 | 0.076 | 0.137 | FALSE |
| rs12764899 | 10 | 102875346 | *AS3MT* | A | G | 0.24 | -0.05 | 0.008 | 1.03E-11 | 0.24 | -0.03 | 0.046 | 0.491 | TRUE |
| rs144814361 | 10 | 119651405 | *BAG3* | T | C | 0.02 | 0.32 | 0.028 | 1.68E-31 | 0.02 | 0.50 | 0.154 | 0.001098 | TRUE |
| rs2234962 | 10 | 119670121 | *BAG3* | C | T | 0.22 | 0.05 | 0.008 | 2.87E-11 | 0.21 | 0.11 | 0.047 | 0.01508 | TRUE |
| rs12284617 | 11 | 83789062 | *DLG2* | G | A | 0.43 | -0.04 | 0.007 | 1.07E-09 | 0.44 | 0.05 | 0.039 | 0.1634 | FALSE |
| rs10750027 | 11 | 113721110 | *ZW10* | A | G | 0.53 | -0.04 | 0.007 | 1.97E-09 | 0.54 | -0.01 | 0.039 | 0.8801 | TRUE |
| rs11223628 | 11 | 133930880 | *IGSF9B* | A | G | 0.38 | 0.05 | 0.007 | 2.2E-14 | 0.36 | 0.11 | 0.066 | 0.1045 | TRUE |
| rs2286028 | 12 | 906303 | *WNK1* | C | G | 0.19 | 0.05 | 0.009 | 3.62E-09 | 0.20 | 0.15 | 0.053 | 0.005501 | TRUE |
| rs28370650 | 12 | 40006146 | *SLC2A13* | A | T | 0.02 | 0.31 | 0.024 | 2.78E-37 | 0.02 | 0.23 | 0.131 | 0.07238 | TRUE |
| rs17443099 | 12 | 40179612 | *LRRK2* | A | G | 0.02 | 0.27 | 0.029 | 3.86E-21 | 0.02 | 0.31 | 0.164 | 0.05955 | TRUE |
| rs76904798 | 12 | 40220632 | *LRRK2* | T | C | 0.14 | 0.11 | 0.010 | 3E-30 | 0.14 | 0.20 | 0.053 | 0.0001999 | TRUE |
| rs35303786 | 12 | 40320097 | *LRRK2* | C | T | 0.02 | 0.18 | 0.028 | 3.51E-10 | 0.02 | 0.30 | 0.139 | 0.02988 | TRUE |
| rs6582585 | 12 | 46022785 | *SCAF11* | C | T | 0.47 | -0.05 | 0.007 | 9.28E-12 | 0.47 | -0.14 | 0.040 | 0.0003333 | TRUE |
| rs933738 | 12 | 49549339 | *KCNH3* | G | A | 0.18 | 0.06 | 0.009 | 7.97E-12 | 0.18 | 0.08 | 0.056 | 0.1707 | TRUE |
| rs61754230 | 12 | 71785666 | *RAB21* | T | C | 0.02 | 0.18 | 0.025 | 3.51E-12 | 0.02 | 0.07 | 0.209 | 0.7201 | TRUE |
| rs7964712 | 12 | 100991017 | *ANO4* | G | A | 0.38 | -0.04 | 0.007 | 0.00000005 | 0.39 | -0.05 | 0.045 | 0.3048 | TRUE |
| rs7350560 | 12 | 107466794 | *ABTB3* | G | A | 0.22 | 0.05 | 0.008 | 1.78E-08 | 0.21 | 0.10 | 0.052 | 0.04544 | TRUE |
| rs2270375 | 12 | 109586736 | *MVK* | G | A | 0.26 | -0.04 | 0.008 | 2.93E-08 | 0.26 | -0.07 | 0.050 | 0.1819 | TRUE |
| rs10847864 | 12 | 122842051 | *HIP1R* | T | G | 0.35 | 0.10 | 0.008 | 4.25E-40 | 0.34 | 0.25 | 0.044 | 2.437E-08 | TRUE |
| rs1198499 | 13 | 49341314 | *CAB39L* | C | T | 0.50 | 0.05 | 0.007 | 4.48E-12 | 0.52 | 0.05 | 0.039 | 0.2001 | TRUE |
| rs1995052 | 13 | 97227087 | *MBNL2* | C | T | 0.66 | -0.04 | 0.007 | 1.03E-08 | 0.66 | -0.14 | 0.041 | 0.0006315 | TRUE |
| rs1805097 | 13 | 109782884 | *IRS2* | T | C | 0.34 | 0.04 | 0.007 | 3.99E-08 | 0.34 | 0.09 | 0.046 | 0.06243 | TRUE |
| rs7155501 | 14 | 54881109 | *GCH1* | G | A | 0.42 | -0.08 | 0.007 | 1.79E-30 | 0.42 | -0.07 | 0.044 | 0.1028 | TRUE |

| | | | | | | | | | | | | | | |
|---|---|---|---|---|---|---|---|---|---|---|---|---|---|---|
| rs979812 | 14 | 87997920 | *GALC* | T | G | 0.44 | 0.04 | 0.007 | 6.49E-11 | 0.43 | 0.06 | 0.039 | 0.09739 | TRUE |
| rs4774417 | 15 | 61701503 | *VPS13C* | A | G | 0.72 | 0.07 | 0.008 | 5.48E-22 | 0.73 | 0.10 | 0.045 | 0.032 | TRUE |
| rs648397 | 15 | 64604181 | *ZNF609* | C | A | 0.97 | 0.16 | 0.027 | 1.2E-08 | 0.98 | 0.17 | 0.176 | 0.3335 | TRUE |
| rs34631560 | 15 | 88973692 | *MFGE8* | A | G | 0.28 | -0.05 | 0.008 | 3.39E-09 | 0.28 | 0.03 | 0.048 | 0.5894 | FALSE |
| rs8058190 | 16 | 1666009 | *CRAMP1* | C | A | 0.54 | -0.05 | 0.007 | 6.61E-13 | 0.56 | -0.01 | 0.058 | 0.8023 | TRUE |
| rs6497339 | 16 | 19266171 | *SYT17* | T | A | 0.54 | -0.06 | 0.007 | 1.39E-17 | 0.55 | -0.08 | 0.045 | 0.07243 | TRUE |
| rs732172 | 16 | 31038712 | *STX4* | T | C | 0.36 | -0.09 | 0.007 | 3.22E-36 | 0.35 | -0.17 | 0.042 | 0.0000697 | TRUE |
| rs1861761 | 16 | 50811622 | *CYLD* | G | T | 0.50 | -0.05 | 0.007 | 3.01E-14 | 0.51 | -0.04 | 0.044 | 0.3309 | TRUE |
| rs8046994 | 16 | 52610012 | *TOX3* | T | C | 0.41 | 0.06 | 0.007 | 7.21E-15 | 0.41 | 0.09 | 0.040 | 0.02937 | TRUE |
| rs1971534 | 16 | 52939375 | *CHD9* | G | A | 0.77 | 0.05 | 0.009 | 2.31E-09 | 0.77 | 0.13 | 0.054 | 0.01644 | TRUE |
| rs2243093 | 17 | 4932600 | *GP1BA* | C | T | 0.13 | 0.06 | 0.010 | 1.71E-09 | 0.13 | 0.04 | 0.059 | 0.5012 | TRUE |
| rs222852 | 17 | 7237287 | *PHF23* | G | A | 0.42 | -0.04 | 0.007 | 1.93E-09 | 0.41 | -0.04 | 0.040 | 0.2674 | TRUE |
| rs72827590 | 17 | 7594953 | *FXR2* | G | A | 0.12 | 0.06 | 0.011 | 2.84E-08 | 0.12 | 0.02 | 0.060 | 0.7017 | TRUE |
| rs2078050 | 17 | 16091574 | *NCOR1* | T | C | 0.55 | -0.04 | 0.007 | 5.27E-11 | 0.56 | -0.03 | 0.039 | 0.4409 | TRUE |
| rs12941356 | 17 | 17813217 | *SREBF1* | G | A | 0.57 | 0.04 | 0.007 | 1.04E-09 | 0.56 | 0.06 | 0.040 | 0.1362 | TRUE |
| rs11080149 | 17 | 31296270 | *OMG* | T | C | 0.13 | 0.06 | 0.010 | 1.61E-10 | 0.13 | 0.06 | 0.059 | 0.325 | TRUE |
| rs12601457 | 17 | 42628829 | *TUBG1* | T | C | 0.27 | -0.05 | 0.008 | 1.48E-10 | 0.27 | -0.11 | 0.074 | 0.1458 | TRUE |
| rs5848 | 17 | 44352876 | *GRN* | T | C | 0.29 | 0.06 | 0.008 | 2.18E-15 | 0.30 | 0.05 | 0.050 | 0.3459 | TRUE |
| rs34359486 | 17 | 44402945 | *GPATCH8/GRN* | A | G | 0.07 | 0.10 | 0.013 | 6.64E-14 | 0.07 | 0.10 | 0.091 | 0.2737 | TRUE |
| rs2532384 | 17 | 46239844 | *KANSL1* | A | C | 0.21 | -0.21 | 0.009 | 1.61E-132 | 0.20 | -0.19 | 0.081 | 0.01761 | TRUE |
| rs190825568 | 17 | 47535476 | *NPEPPS* | A | G | 0.13 | -0.06 | 0.010 | 3.5E-08 | 0.13 | -0.04 | 0.099 | 0.7168 | TRUE |
| rs847680 | 17 | 50146710 | *PPP1R9B* | C | T | 0.17 | -0.05 | 0.009 | 8.85E-09 | 0.17 | -0.06 | 0.060 | 0.3094 | TRUE |
| rs11871753 | 17 | 61779284 | *BRIP1* | G | A | 0.74 | 0.05 | 0.008 | 1.38E-08 | 0.75 | 0.09 | 0.052 | 0.07398 | TRUE |
| rs72843781 | 17 | 62016181 | *MED13* | C | A | 0.35 | 0.04 | 0.008 | 1.15E-08 | 0.35 | 0.22 | 0.069 | 0.001585 | TRUE |
| rs58426209 | 17 | 68281293 | *SLC16A6* | G | T | 0.20 | -0.06 | 0.009 | 5.85E-11 | 0.21 | -0.26 | 0.084 | 0.002126 | TRUE |
| rs8074498 | 17 | 81996668 | *ASPSCR1* | A | T | 0.57 | 0.04 | 0.007 | 4.62E-09 | 0.59 | 0.04 | 0.045 | 0.3951 | TRUE |
| rs12456492 | 18 | 43093415 | *RIT2* | G | A | 0.32 | 0.07 | 0.007 | 3.11E-22 | 0.32 | 0.10 | 0.041 | 0.01844 | TRUE |
| rs1128402 | 19 | 2353152 | *SPPL2B* | A | C | 0.21 | 0.05 | 0.008 | 6.1E-09 | 0.21 | 0.05 | 0.048 | 0.3223 | TRUE |
| rs8105994 | 19 | 18482743 | *ELL* | C | T | 0.34 | -0.04 | 0.007 | 6.85E-10 | 0.34 | 0.00 | 0.046 | 0.9285 | TRUE |
| rs11669800 | 19 | 32563836 | *PDCD5* | A | G | 0.02 | 0.14 | 0.024 | 6.45E-09 | 0.02 | 0.24 | 0.197 | 0.2317 | TRUE |
| rs4802574 | 19 | 49145273 | *PPFIA3* | G | A | 0.44 | -0.04 | 0.007 | 1.45E-09 | 0.46 | -0.07 | 0.044 | 0.1394 | TRUE |
| rs55785911 | 20 | 3172857 | *LZTS3* | A | G | 0.36 | -0.04 | 0.007 | 2.09E-09 | 0.37 | -0.16 | 0.042 | 0.00007896 | TRUE |
| rs117517602 | 20 | 50535462 | *PTPN1* | C | T | 0.08 | 0.07 | 0.013 | 4.19E-08 | 0.08 | 0.06 | 0.081 | 0.4313 | TRUE |
| rs11701836 | 21 | 37519947 | *DYRK1A* | G | A | 0.26 | 0.06 | 0.008 | 2.25E-17 | 0.27 | 0.13 | 0.049 | 0.009237 | TRUE |
| rs4816690 | 21 | 40577012 | *DSCAM* | C | T | 0.49 | 0.04 | 0.007 | 2.59E-08 | 0.49 | 0.09 | 0.043 | 0.03045 | TRUE |
| rs4148103 | 21 | 42221495 | *ABCG1* | A | G | 0.08 | -0.08 | 0.013 | 7.75E-10 | 0.08 | -0.16 | 0.088 | 0.07069 | TRUE |
| rs118075805 | 21 | 44366954 | *TRPM2* | T | G | 0.02 | 0.20 | 0.034 | 4.03E-09 | 0.02 | 0.41 | 0.218 | 0.06205 | TRUE |
| rs8130097 | 21 | 45161847 | *ADARB1* | A | C | 0.05 | 0.10 | 0.016 | 8.04E-10 | 0.05 | 0.32 | 0.092 | 0.0004519 | TRUE |

**Significant signals from Nalls et al. 2019 GWAS compared to YOPD additive GWAS summary statistics (88 overlapping SNPs)**

| rsID | CHR | BP | Nearest gene | Effect allele | Other allele | Freq Nalls | Beta Nalls | SE Nalls | P_Nalls | Freq YOPD | Beta YOPD | SE YOPD | P YOPD | Effect direction contcordane |
|---|---|---|---|---|---|---|---|---|---|---|---|---|---|---|
| rs114138760 | 1 | 154925709 | *PMVK* | C | G | 0.01 | 0.28 | 0.048 | 4.19E-09 | 0.01 | 0.66 | 0.162 | 0.00004742 | TRUE |
| rs35749011 | 1 | 155162560 | *KRTCAP2* | A | G | 0.02 | 0.61 | 0.034 | 1.72E-70 | 0.01 | 0.70 | 0.137 | 3.108E-07 | TRUE |
| rs6658353 | 1 | 161499264 | *FCGR2A* | C | G | 0.50 | 0.07 | 0.009 | 6.1E-12 | 0.49 | 0.11 | 0.040 | 0.007029 | TRUE |
| rs11578699 | 1 | 171750629 | *VAMP4* | T | C | 0.19 | -0.07 | 0.012 | 4.47E-09 | 0.20 | 0.01 | 0.049 | 0.8229 | FALSE |

| | | | | | | | | | | | | | | |
|---|---|---|---|---|---|---|---|---|---|---|---|---|---|---|
| rs823118 | 1 | 205754444 | *NUCKS1* | T | C | 0.57 | 0.11 | 0.009 | 1.11E-29 | 0.56 | 0.13 | 0.040 | 0.001333 | TRUE |
| rs11557080 | 1 | 205768611 | *RAB29* | A | G | 0.14 | 0.13 | 0.014 | 2.5E-22 | 0.13 | 0.14 | 0.055 | 0.01021 | TRUE |
| rs4653767 | 1 | 226728377 | *ITPKB* | C | T | 0.28 | -0.08 | 0.010 | 1.38E-15 | 0.29 | -0.12 | 0.044 | 0.005947 | TRUE |
| rs10797576 | 1 | 232528865 | *SIPA1L2* | T | C | 0.14 | 0.11 | 0.013 | 6.84E-17 | 0.13 | 0.16 | 0.055 | 0.004758 | TRUE |
| rs76116224 | 2 | 17966582 | *KCNS3* | T | A | 0.10 | -0.11 | 0.019 | 1.27E-08 | 0.10 | -0.18 | 0.080 | 0.02563 | TRUE |
| rs2042477 | 2 | 95335195 | *KCNIP3* | T | A | 0.76 | 0.07 | 0.012 | 1.38E-08 | 0.76 | 0.07 | 0.053 | 0.216 | TRUE |
| rs11683001 | 2 | 101780501 | *MAP4K4* | A | T | 0.34 | 0.07 | 0.010 | 8.04E-13 | 0.32 | 0.07 | 0.041 | 0.1091 | TRUE |
| rs57891859 | 2 | 134707046 | *TMEM163* | G | A | 0.28 | -0.08 | 0.011 | 4.55E-14 | 0.29 | -0.05 | 0.044 | 0.2981 | TRUE |
| rs1474055 | 2 | 168253884 | *STK39* | T | C | 0.13 | 0.18 | 0.014 | 2.54E-39 | 0.12 | 0.24 | 0.056 | 0.00001497 | TRUE |
| rs73038319 | 3 | 18320267 | *SATB1* | C | A | 0.04 | 0.17 | 0.024 | 5.94E-13 | 0.04 | 0.20 | 0.102 | 0.05113 | TRUE |
| rs6808178 | 3 | 28664199 | *LINC00693* | C | T | 0.62 | -0.07 | 0.010 | 8.09E-12 | 0.63 | -0.11 | 0.040 | 0.005005 | TRUE |
| rs12497850 | 3 | 48711556 | *IP6K2* | T | G | 0.65 | 0.06 | 0.010 | 1.36E-10 | 0.64 | 0.01 | 0.041 | 0.7997 | TRUE |
| rs55961674 | 3 | 122478045 | *KPNA1* | T | C | 0.17 | 0.09 | 0.013 | 9.98E-12 | 0.17 | 0.06 | 0.051 | 0.2261 | TRUE |
| rs11707416 | 3 | 151391177 | *MED12L* | A | T | 0.37 | -0.06 | 0.010 | 1.13E-10 | 0.38 | -0.05 | 0.040 | 0.1842 | TRUE |
| rs1450522 | 3 | 161359842 | *SPTSSB* | G | A | 0.33 | 0.06 | 0.010 | 5.01E-10 | 0.32 | 0.05 | 0.042 | 0.2375 | TRUE |
| rs10513789 | 3 | 183042285 | *MCCC1* | G | T | 0.19 | -0.15 | 0.012 | 1.22E-34 | 0.20 | -0.18 | 0.052 | 0.0005137 | TRUE |
| rs873786 | 4 | 931588 | *GAK* | T | C | 0.10 | -0.17 | 0.018 | 1.79E-21 | 0.11 | -0.14 | 0.067 | 0.03499 | TRUE |
| rs34311866 | 4 | 958159 | *TMEM175* | C | T | 0.19 | 0.21 | 0.012 | 9.98E-70 | 0.18 | 0.21 | 0.048 | 0.00001795 | TRUE |
| rs4698412 | 4 | 15735725 | *BST1* | A | G | 0.55 | 0.10 | 0.009 | 2.06E-28 | 0.54 | 0.18 | 0.039 | 4.564E-06 | TRUE |
| rs34025766 | 4 | 17967188 | *LCORL* | A | T | 0.16 | -0.08 | 0.013 | 2.87E-10 | 0.16 | -0.18 | 0.056 | 0.001526 | TRUE |
| rs6825004 | 4 | 76189212 | *SCARB2* | G | C | 0.31 | -0.06 | 0.010 | 1.17E-09 | 0.31 | 0.00 | 0.042 | 0.9071 | FALSE |
| rs4101061 | 4 | 76226816 | *FAM47E* | G | A | 0.29 | 0.09 | 0.010 | 4.97E-19 | 0.28 | 0.05 | 0.043 | 0.2587 | TRUE |
| rs6854006 | 4 | 76276901 | *FAM47E-STBD1* | T | C | 0.36 | -0.09 | 0.010 | 5.82E-21 | 0.36 | -0.08 | 0.041 | 0.04183 | TRUE |
| rs356182 | 4 | 89704960 | *SNCA* | A | G | 0.63 | -0.28 | 0.011 | 3.89E-154 | 0.65 | -0.38 | 0.044 | 8.951E-18 | TRUE |
| rs5019538 | 4 | 89715479 | *SNCA* | A | G | 0.68 | -0.16 | 0.012 | 1.13E-36 | 0.70 | -0.22 | 0.041 | 5.95E-08 | TRUE |
| rs13117519 | 4 | 113447909 | *CAMK2D* | T | C | 0.17 | 0.09 | 0.012 | 9.82E-13 | 0.17 | 0.15 | 0.052 | 0.002855 | TRUE |
| rs62333164 | 4 | 169662006 | *CLCN3* | A | G | 0.33 | -0.06 | 0.010 | 2E-10 | 0.33 | -0.06 | 0.042 | 0.1232 | TRUE |
| rs1867598 | 5 | 60842132 | *ELOVL7* | G | A | 0.10 | 0.16 | 0.016 | 2.52E-23 | 0.10 | 0.28 | 0.061 | 5.428E-06 | TRUE |
| rs26431 | 5 | 103030090 | *PAM* | C | G | 0.70 | 0.06 | 0.010 | 1.57E-09 | 0.69 | 0.05 | 0.043 | 0.2457 | TRUE |
| rs11950533 | 5 | 134863415 | *C5orf24* | A | C | 0.10 | -0.09 | 0.016 | 7.16E-09 | 0.10 | -0.21 | 0.070 | 0.002645 | TRUE |
| rs4140646 | 6 | 27771022 | *LOC100131289* | A | G | 0.21 | 0.08 | 0.012 | 5.62E-12 | 0.23 | 0.03 | 0.049 | 0.5175 | TRUE |
| rs9261484 | 6 | 30140906 | *TRIM40* | T | C | 0.25 | -0.06 | 0.011 | 1.62E-08 | 0.24 | -0.08 | 0.048 | 0.07786 | TRUE |
| rs504594 | 6 | 32610995 | *HLA-DRB5* | A | C | 0.16 | -0.17 | 0.015 | 6.96E-28 | 0.16 | -0.13 | 0.062 | 0.035 | TRUE |
| rs12528068 | 6 | 71778059 | *RIMS1* | T | C | 0.28 | 0.07 | 0.010 | 1.63E-10 | 0.28 | 0.06 | 0.043 | 0.1952 | TRUE |
| rs997368 | 6 | 111922088 | *FYN* | G | A | 0.20 | -0.07 | 0.012 | 1.84E-09 | 0.20 | -0.13 | 0.050 | 0.01252 | TRUE |
| rs75859381 | 6 | 132889222 | *RPS12* | C | T | 0.03 | 0.22 | 0.034 | 1.04E-10 | 0.03 | 0.17 | 0.161 | 0.3008 | TRUE |
| rs199351 | 7 | 23260430 | *GPNMB* | C | A | 0.41 | -0.10 | 0.010 | 5.25E-26 | 0.42 | -0.08 | 0.040 | 0.05257 | TRUE |
| rs76949143 | 7 | 66544864 | *GS1-124K5.11* | A | T | 0.05 | -0.14 | 0.025 | 1.43E-08 | 0.05 | 0.03 | 0.105 | 0.7779 | FALSE |
| rs1293298 | 8 | 11854934 | *CTSB* | C | A | 0.26 | -0.09 | 0.011 | 3.99E-16 | 0.26 | -0.05 | 0.045 | 0.3131 | TRUE |
| rs620513 | 8 | 16840084 | *FGF20* | T | G | 0.27 | -0.09 | 0.011 | 2.72E-15 | 0.27 | -0.11 | 0.045 | 0.01028 | TRUE |
| rs2280104 | 8 | 22668467 | *BIN3* | C | T | 0.64 | -0.06 | 0.010 | 1.16E-08 | 0.64 | -0.08 | 0.040 | 0.05289 | TRUE |
| rs2086641 | 8 | 129889663 | *FAM49B* | C | T | 0.28 | 0.06 | 0.011 | 1.81E-08 | 0.27 | 0.15 | 0.048 | 0.001428 | TRUE |
| rs13294100 | 9 | 17579692 | *SH3GL2* | G | T | 0.66 | 0.09 | 0.010 | 8.72E-18 | 0.65 | 0.12 | 0.041 | 0.004216 | TRUE |
| rs10756907 | 9 | 17727067 | *SH3GL2* | G | A | 0.23 | 0.09 | 0.011 | 5.06E-17 | 0.24 | 0.11 | 0.045 | 0.01267 | TRUE |
| rs6476434 | 9 | 34046393 | *UBAP2* | T | C | 0.73 | -0.06 | 0.011 | 6.58E-09 | 0.73 | -0.10 | 0.043 | 0.02067 | TRUE |

| | | | | | | | | | | | | | | |
|---|---|---|---|---|---|---|---|---|---|---|---|---|---|---|
| rs896435 | 10 | 15515407 | *ITGA8* | T | C | 0.69 | 0.07 | 0.010 | 3.41E-13 | 0.69 | 0.14 | 0.043 | 0.001396 | TRUE |
| rs10748818 | 10 | 102255522 | *GBF1* | G | A | 0.15 | 0.08 | 0.013 | 1.05E-09 | 0.15 | -0.14 | 0.058 | 0.01654 | FALSE |
| rs72840788 | 10 | 119656173 | *BAG3* | A | G | 0.22 | 0.08 | 0.011 | 1.57E-11 | 0.21 | 0.12 | 0.047 | 0.01213 | TRUE |
| rs117896735 | 10 | 119776815 | *INPP5F* | A | G | 0.02 | 0.44 | 0.039 | 2.36E-28 | 0.02 | 0.45 | 0.155 | 0.00399 | TRUE |
| rs7938782 | 11 | 10537230 | *RNF141* | G | A | 0.12 | -0.09 | 0.015 | 2.12E-09 | 0.13 | -0.17 | 0.064 | 0.008545 | TRUE |
| rs12283611 | 11 | 83776234 | *DLG2* | A | C | 0.41 | -0.06 | 0.010 | 2.61E-10 | 0.42 | 0.02 | 0.039 | 0.5359 | FALSE |
| rs3802920 | 11 | 133917106 | *IGSF9B* | T | G | 0.21 | 0.11 | 0.012 | 6.26E-20 | 0.20 | 0.12 | 0.048 | 0.01427 | TRUE |
| rs76904798 | 12 | 40220632 | *LRRK2* | T | C | 0.14 | 0.14 | 0.013 | 1.52E-28 | 0.14 | 0.20 | 0.053 | 0.0001999 | TRUE |
| rs7134559 | 12 | 46025303 | *SCAF11* | T | C | 0.40 | -0.05 | 0.010 | 3.96E-08 | 0.40 | -0.14 | 0.041 | 0.0004876 | TRUE |
| rs10847864 | 12 | 122842051 | *HIP1R* | T | G | 0.36 | 0.15 | 0.012 | 1.47E-37 | 0.34 | 0.25 | 0.044 | 2.437E-08 | TRUE |
| rs11610045 | 12 | 132487182 | *FBRSL1* | A | G | 0.49 | 0.06 | 0.009 | 1.77E-10 | 0.49 | 0.00 | 0.039 | 0.9159 | TRUE |
| rs9568188 | 13 | 49353596 | *CAB39L* | C | T | 0.26 | -0.06 | 0.011 | 1.15E-08 | 0.26 | -0.03 | 0.045 | 0.4614 | TRUE |
| rs4771268 | 13 | 97212767 | *MBNL2* | C | T | 0.77 | -0.07 | 0.011 | 1.45E-09 | 0.77 | -0.08 | 0.046 | 0.09994 | TRUE |
| rs12147950 | 14 | 37520065 | *MIPOL1* | C | T | 0.56 | 0.05 | 0.010 | 3.54E-08 | 0.56 | 0.13 | 0.040 | 0.0009351 | TRUE |
| rs11158026 | 14 | 54882151 | *GCH1* | T | C | 0.32 | -0.08 | 0.010 | 1.66E-16 | 0.32 | -0.06 | 0.043 | 0.179 | TRUE |
| rs3742785 | 14 | 74906331 | *RPS6KL1* | C | A | 0.21 | -0.07 | 0.012 | 1.92E-09 | 0.21 | -0.04 | 0.054 | 0.4901 | TRUE |
| rs979812 | 14 | 87997920 | *GALC* | T | G | 0.44 | 0.06 | 0.009 | 6.19E-11 | 0.43 | 0.06 | 0.039 | 0.09739 | TRUE |
| rs2251086 | 15 | 61705186 | *VPS13C* | C | T | 0.86 | 0.12 | 0.014 | 6.08E-18 | 0.86 | 0.17 | 0.059 | 0.003804 | TRUE |
| rs6497339 | 16 | 19266171 | *SYT17* | T | A | 0.55 | -0.06 | 0.010 | 2.76E-11 | 0.55 | -0.08 | 0.045 | 0.07243 | TRUE |
| rs2904880 | 16 | 28933075 | *CD19* | G | C | 0.69 | 0.07 | 0.011 | 7.87E-10 | 0.69 | 0.08 | 0.043 | 0.0662 | TRUE |
| rs11150601 | 16 | 30966478 | *SETD1A* | A | G | 0.64 | 0.09 | 0.010 | 5.12E-20 | 0.64 | 0.17 | 0.042 | 0.00005336 | TRUE |
| rs6500328 | 16 | 50702745 | *NOD2* | G | A | 0.40 | -0.06 | 0.010 | 1.82E-09 | 0.40 | -0.05 | 0.045 | 0.2541 | TRUE |
| rs3104783 | 16 | 52602330 | *CASC16* | A | C | 0.43 | 0.07 | 0.009 | 1.29E-12 | 0.42 | 0.10 | 0.039 | 0.01218 | TRUE |
| rs10221156 | 16 | 52935514 | *CHD9* | A | G | 0.09 | -0.12 | 0.018 | 1.08E-10 | 0.10 | -0.12 | 0.078 | 0.1221 | TRUE |
| rs12600861 | 17 | 7452302 | *CHRNB1* | C | A | 0.35 | 0.06 | 0.010 | 1.01E-08 | 0.35 | 0.04 | 0.041 | 0.3149 | TRUE |
| rs12951632 | 17 | 42588995 | *RETREG3* | C | T | 0.27 | -0.06 | 0.011 | 1.4E-09 | 0.27 | -0.03 | 0.044 | 0.5701 | TRUE |
| rs2269906 | 17 | 44216969 | *UBTF* | C | A | 0.35 | -0.06 | 0.010 | 6.24E-10 | 0.34 | -0.05 | 0.046 | 0.3278 | TRUE |
| rs850738 | 17 | 44357262 | *FAM171A2* | G | A | 0.39 | 0.07 | 0.011 | 1.29E-11 | 0.40 | 0.08 | 0.047 | 0.1045 | TRUE |
| rs62053943 | 17 | 45666837 | *CRHR1* | T | C | 0.16 | -0.27 | 0.016 | 3.58E-68 | 0.15 | -0.21 | 0.063 | 0.001187 | TRUE |
| rs117615688 | 17 | 45720942 | *CRHR1* | A | G | 0.07 | -0.23 | 0.029 | 6.71E-16 | 0.06 | -0.09 | 0.119 | 0.4252 | TRUE |
| rs11658976 | 17 | 46789439 | *WNT3* | A | G | 0.58 | -0.06 | 0.011 | 3.52E-08 | 0.61 | 0.03 | 0.045 | 0.5481 | FALSE |
| rs61169879 | 17 | 61840005 | *BRIP1* | T | C | 0.16 | 0.08 | 0.013 | 9.28E-10 | 0.16 | 0.15 | 0.057 | 0.008905 | TRUE |
| rs666463 | 17 | 78429399 | *DNAH17* | T | A | 0.17 | 0.08 | 0.013 | 3.2E-09 | 0.18 | 0.01 | 0.052 | 0.9195 | TRUE |
| rs1941685 | 18 | 33724354 | *ASXL3* | T | G | 0.50 | 0.05 | 0.009 | 1.69E-08 | 0.49 | 0.04 | 0.040 | 0.3523 | TRUE |
| rs12456492 | 18 | 43093415 | *RIT2* | G | A | 0.32 | 0.10 | 0.010 | 3.8E-23 | 0.32 | 0.10 | 0.041 | 0.01844 | TRUE |
| rs8087969 | 18 | 51157219 | *MEX3C* | G | T | 0.45 | -0.06 | 0.010 | 1.41E-08 | 0.44 | 0.06 | 0.044 | 0.2026 | FALSE |
| rs55818311 | 19 | 2341049 | *SPPL2B* | T | C | 0.69 | -0.07 | 0.011 | 4.18E-10 | 0.69 | -0.03 | 0.048 | 0.479 | TRUE |
| rs77351827 | 20 | 6025395 | *CRLS1* | T | C | 0.13 | 0.08 | 0.014 | 8.87E-09 | 0.12 | 0.12 | 0.058 | 0.03599 | TRUE |
| rs2248244 | 21 | 37480059 | *DYRK1A* | A | G | 0.28 | 0.07 | 0.011 | 2.74E-11 | 0.28 | 0.14 | 0.048 | 0.002859 | TRUE |

**Supplementary Table 3 Results from stepwise conditional analysis of the *SNCA* locus in GP2 data**

| SNP | Position with alleles | Frequency | Conditioned on | OR (95% CI) | P-value |
|---|---|---|---|---|---|
| rs2869998 | chr4:89557936:C:A | 0.22 | rs356182 | 1.38 (1.19-1.60) | $2.5 \times 10^{-5}$ |
| rs182585138 | chr4:89602489:T:C | 0.048 | rs356182 and rs2869998 | 1.68 (1.31-2.16) | $4.8 \times 10^{-5}$ |
| rs187935141 | chr4:89269880:G:A | 0.012 | rs356182 and rs2869998 and rs182585138 | 2.64 (1.60-4.39) | 0.00016 |
| rs7681154 | chr4:89842552:A:C | 0.49 | rs356182 and rs2869998 | 0.87 (0.76-1.00) | 0.042 |

Results are based on individual level data from the GP2 cohort only. Results from forward stepwise logistic regression including sex and five principal components are reported, starting with conditioning on the top-SNP rs356182 and including SNPs with $P < 0.0023$ (see main manuscript). Note that while rs2869998 is in high LD with a previously published genome-wide significant independent association signal, the other two SNPs have low allele frequency and have not been replicated. We also show the most significant result for rs7681154, a SNP representing the independent 5' *SNCA* association signal highlighted in multiple previous studies (see main text).

**Supplementary Table 4. Exploration of genomic inflation in the recessive model GWAS**

| Subcohort or SNP subset | N SNPs | $Lambda_{1000}$ |
|---|---|---|
| All SNPs in meta-analysis | 6,875,354 | 1.17 |
| MAF > 5% | 5,749,753 | 1.13 |
| Present in ≥ 5 cohorts | 4,186,319 | 1.10 |
| Heterogeneity $I^2 < 50$ | 6,523,174 | 1.16 |
| GP2 | 8,212,110 | 0.82 |
| NIA | 4,281,280 | 1.0 |
| Spain | 6,053,305 | 0.84 |
| Dutch | 4,049,713 | 1.0 |
| Oslo | 3,733,800 | 1.0 |
| IPDGC NeuroX | 589,155 | 0.99 |
| Germany | 4,066,547 | 1.0 |
| Tubi | 3,423,921 | 1.0 |
| Hamza | 6,739,399 | 0.77 |

**Supplementary Table 5. Odds ratios and genotype frequencies of significant recessive model lead SNPs (excluding *SNCA*)**

| rsID | Coordinate (chr:bp) | Locus | Effect allele | Non effect allele | OR recessive model | OR allelic model | Frequency effect allele | Frequency of minor allele homozyous genotype in PD | Frequency of minor allele homozygous genotype in controls | Expected frequency of minor allele homozygous genotype given HWE | Fold enrichment of minor allele homozygous genotype relative to expected under HWE in PD | Fold enrichment of minor allele homozygous genotype relative to expected under HWE in controls |
|---|---|---|---|---|---|---|---|---|---|---|---|---|
| rs11579040 | 1:239009594 | *CHRM3* | T | C | 29.67 | 1.03 | 0.0584 | 0.015 | 0.0024 | 0.0034 | 4.4 | 0.7 |
| rs12139868 | 1:243750673 | *AKT3* | A | C | 6.51 | 1.13 | 0.0669 | 0.018 | 0.0030 | 0.0045 | 4.1 | 0.7 |
| rs10008290 | 4:106578771 | *LOC105377356* | G | A | 15.88 | 1.06 | 0.037 | 0.0089 | 0.0010 | 0.0014 | 6.5 | 0.7 |
| rs17529104 | 6:162386075 | *PRKN* | C | T | 23.46 | 1.59 | 0.0589 | 0.037 | 0.0031 | 0.0035 | 10.7 | 0.9 |
| rs35859528 | 7:15830177 | *MEOX2* | C | T | 10.04 | 1.27 | 0.0465 | 0.011 | 0.0018 | 0.0022 | 5.1 | 0.8 |
| rs62446380 | 7:52896452 | *POM121L12* | C | T | 3.69 | 1.24 | 0.1222 | 0.040 | 0.014 | 0.015 | 2.7 | 0.9 |
| rs1263642 | 14:22545204 | *TRAC* | A | G | 0.45 | 0.82 | 0.8312 | 0.055 | 0.027 | 0.028 | 1.9 | 0.9 |
| rs145087783 | 17:22204435 | *UBBP4* | A | G | 2.68 | 1.37 | 0.2425 | 0.110 | 0.058 | 0.059 | 1.9 | 1.0 |

**Supplementary Figure 1. Q-Q plot for the additive model GWAS**

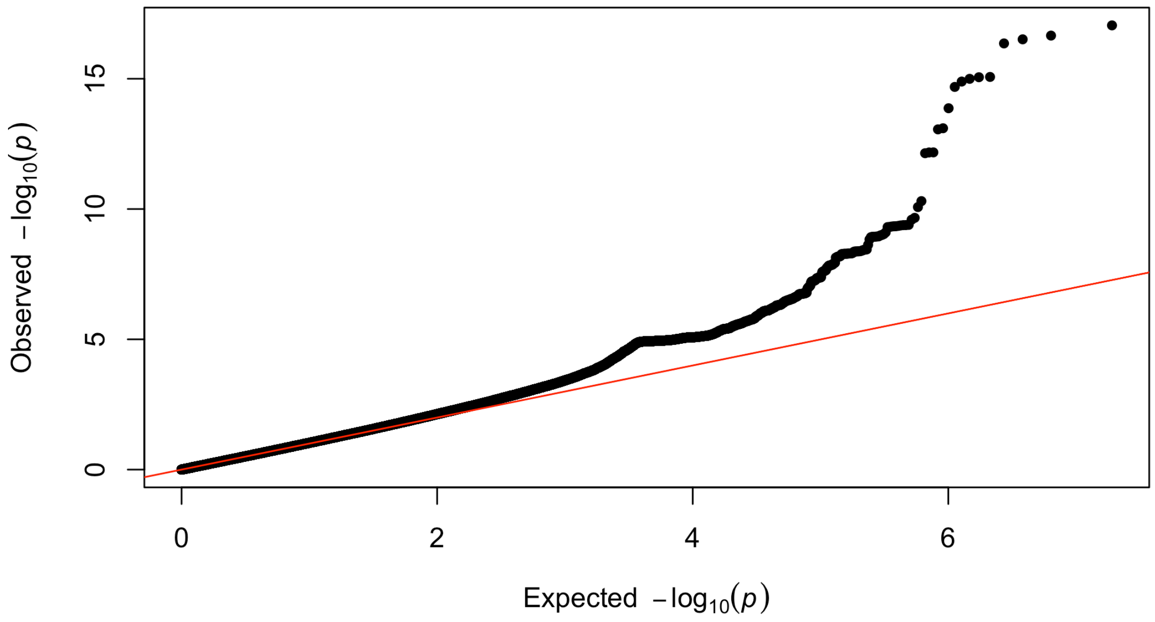

**Supplementary Figure 2. Local Manhattan (LocusZoom) plots for significant additive model signals**

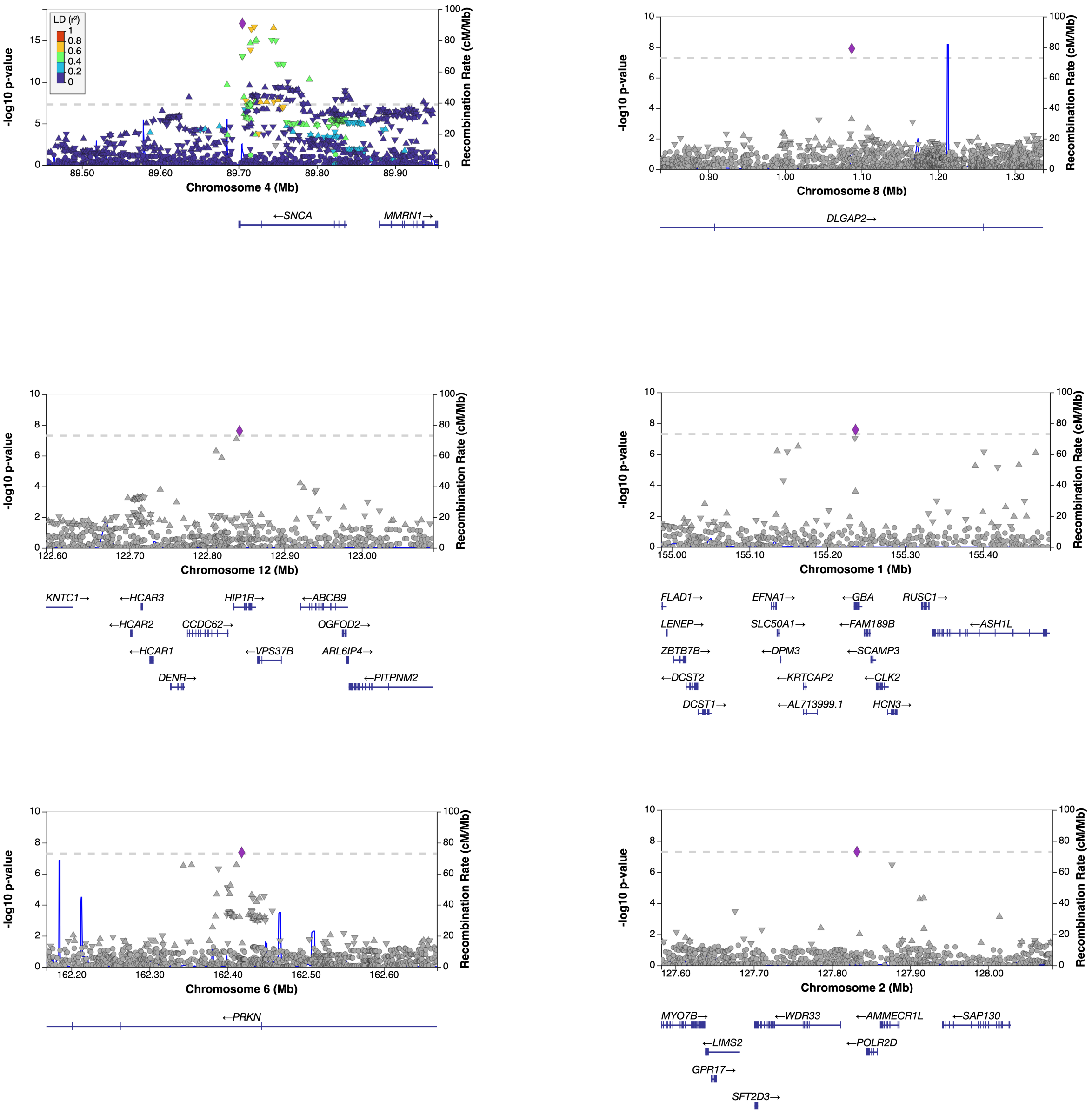


Coordnates correspond to hg38 genome build. Plots were generated using LocusZoom.[8]

**Supplementary Figure 3. Forest plot of the significant signal from the PRKN locus in the additive model GWAS**

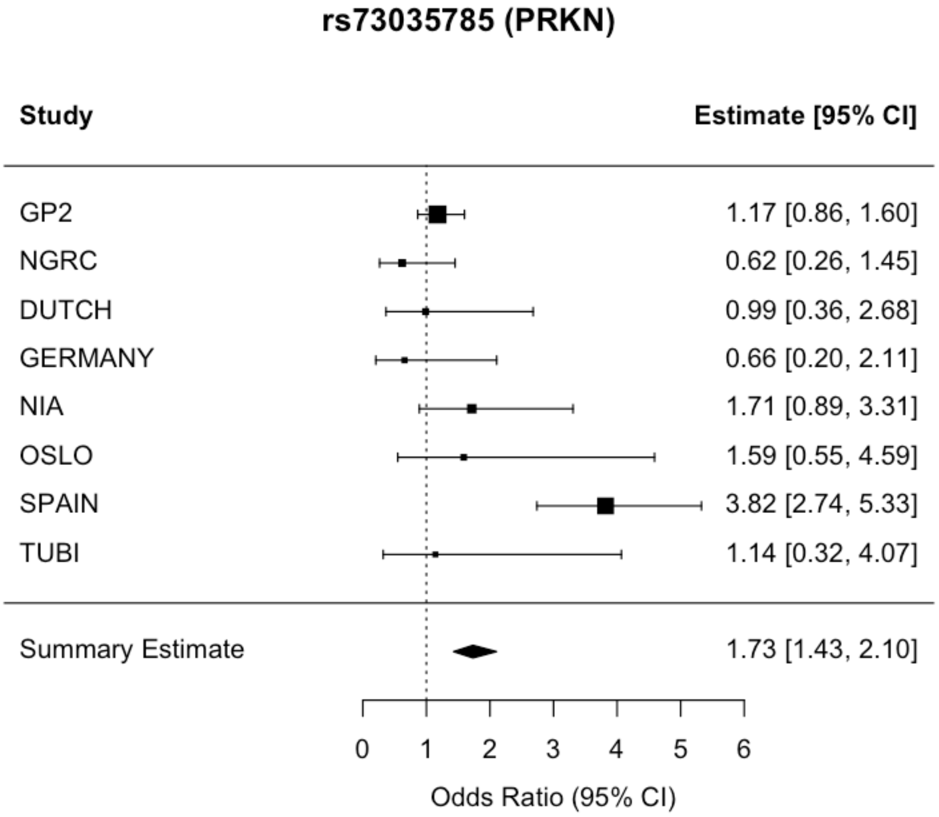


The plot shows how the association is almost completely driven by the Spanish subcohort

**Supplementary Methods**

The work of this study was conducted at different times across different data environments for the IPDGC, GP2 and NGRC subcohorts. As a consequence, integrated analysis of individual-level data for all participants was not possible. There is a known sample overlap between samples and subcohorts included in the datasets of the IPDGC and GP2 consortia. As relatedness analyses based on individual level data across these datasets were not an available option, a conservative approach was taken to only include GP2 subcohorts with a minimal risk of sample overlap: GP2 cohorts were considered to be independent from the IPDGC cohorts if they originated in full from countries not contributing to the IPDGC data used in this study (i.e. the Netherlands, Norway, Spain, UK, France, Greece and the USA). Furthermore, we considered the risk of overlap as minimal when GP2 data originated from sample collecting studies where institutions or principal investigators had no overlap with institutions or co-authors in either Blauwendraat 2019 age at onset GWAS[9], Nalls 2014 GWAS, or any of the IPDGC-cohort specific papers.[3,5,6,10] Finally, for GP2 cohorts that included particularly many YOPD cases or controls where the data originated from the same country or institution, or had a principal investigator mentioned as an author in one of the articles listed above, we communicated directly with the GP2 cohort's principal investigator regarding the origin of the GP2 samples and known overlap with IPDGC. This led to exclusion of some subcohorts, inclusion of others, and in one instance (Tuebingen), exclusion of specific participants known to overlap across IPDGC and GP2. In cases of uncertainty, the subcohort in question was excluded from our analyses. We excluded cohorts if the total number of YOPD cases was less than 10 or the total number of controls was less than 20 from the relevant country of origin.

After selection of GP2 subcohorts, we performed further filtering at the individual level using following principles:
- Only individuals with European ancestry were included (GP2 variable "nba_label" = "EUR", GenoTools genetically predicted ancestry)
- Only individuals from the complex network in GP2 were included (and not cases recruited through the monogenic network).
- We included only individuals labeled as PD or Controls at baseline (GP2 variable "baseline_GP2_phenotype_for_qc")

- We excluded individuals pruned from the genetic data QC as performed previously by the GP2
- We removed one from each pair of related individuals, ensuring that no individuals had a KING kinship coefficient >0.0884 (corresponding to relatedness closer than second cousins).

The final GP2 dataset used in the study included N=608 YOPD cases and N=4635 cases from 11 different countries: Australia, Brasil, Canada, Germany, Great Briatain, Moldova, Norway, New zealand, USA and South Africa. The participants were enrolled in 31 different GP2 cohorts.
