## Supplementary material for "A genome-wide association study of young onset Parkinson’s disease in European ancestry": GP2 members

### Banner authorship list of Global Parkinson's Genetics Program (GP2) members (as of GP2 release 11)

| Country | Name | Email address | Institution | City | (USA, Canada, | Funders and Disclosures |
|---|---|---|---|---|---|---|
| Algeria | Yasser Mecheri | | Centre Hospitalo-Universitaire Dr Benbadis Constantine | Constantine | | Nothing to declare |
| Algeria | Bouchetara Mohamed Sofiane | | Hospital university of Oran-Algeria | Oran | | Nothing to declare |
| Algeria | Benhassine Traki | | Faculty of Biological Sciences, USTHB Bab Ezzouar, Algiers | Algiers | | Nothing to declare |
| Argentina | Emilia M Gatto | | Sanatorio de la Trinidad Mitre- INEBA | Buenos Aires | | Nothing to declare |
| Argentina | Marcelo Kauffman | | Hospital JM Ramos Mejia | Buenos Aires | | Nothing to declare |
| Argentina | Federico Capparelli | | Centro de Educación Médica e Investigaciones Clínicas Norberto Quirno | Buenos Aires | | Nothing to declare |
| Argentina | Maria Valentina Muller | | Hospital General San Martin | La Plata | | Nothing to declare |
| Argentina | Marcela Susana Tela | | Hospital Fernandez | Buenos Aires | | Nothing to declare |
| Argentina | Adamec, Dario Sergio | | HOSPITAL NACIONAL PROFESOR ALEJANDRO POSADAS | Buenos Aires | | Nothing to declare |
| Argentina | Cesar Luis Avila | | CONICET-UNT | San Miquel de Tucumán | | Nothing to declare |
| Argentina | Valentin Selvaggi | | Hospital Ramos Mejia | Buenos Aires | | Nothing to declare |
| Argentina | Nahuel Fonseca | | Consejo Nacional de Investigaciones Científicas y Técnicas | Buenos Aires | | Nothing to declare |
| Argentina | Elena AVALE | | CONICET | Buenos Aires | | Nothing to declare |
| Argentina | Marcelo Merello | | Fleni | Buenos Aires | | Nothing to declare |
| Argentina | Griselda Judith Alvarado | | SIPROSA | Tucumán | | Nothing to declare |
| Argentina | Juan Pablo Diaz Rearte | | SIPROSA | Tucumán | | Nothing to declare |
| Argentina | Luciana Rojas Vazquez | | SIPROSA | Tucumán | | Nothing to declare |
| Argentina | Lucía Wang | | PARKINSON ARGENTINA/ALAPA | Buenos Aires | | Nothing to declare |
| Argentina | Carlos Matias López Razquin | | H.I.G.A. "Gral. San Martín" - ALAPA | La Plata | | Nothing to declare |
| Argentina | Andrea Bril | Av Cerviño 3356 | Hospital Fernández | Buenos Aires | | Nothing to declare |
| Argentina | Florencia Nicole Wainberg | | Fleni | Buenos Aires | | Nothing to declare |
| Argentina | Maria Fernanda López | r | Hospital Nacional Alejandro Posadas. Argentina | El Palomar | | Nothing to declare |
| Armenia | Samson Khachatryan | | Somnus Neurology Clinic | Yerevan | | Nothing to declare |
| Armenia | Zaruhi Tavadyan | | Somnus Neurology Clinic | Yerevan | | Nothing to declare |
| Armenia | Mariam Isayan | | Somnus Neurology Clinic | Yerevan | | Nothing to declare |
| Australia | Claire E Shepherd | | Neuroscience Research Australia | Sydney | Wales | The Sydney Brain Bank is located at and supported by Neuroscience Research Australia |
| Australia | Kishore Kumar | | Garvan Institute of Medical Research and Concord Repatriation General Hospital | Darlinghurst | Wales | Paul Ainsworth Family Foundation |
| Australia | Melina Ellis | | Concord Hospital | Concord | Wales | Nothing to declare |
| Australia | Miguel E. Rentería | | QIMR Berghofer Medical Research Institute | Herston | Queensland | Foundation and The Michael J. Fox Foundation for Parkinson's Research |
| Australia | Sulev Koks | | Murdoch University | Perth | Australia | Nothing to declare |
| Australia | Simon Rowe | | Neuroscience Research Australia | Sydney | New South Wales | Nothing to declare |
| Australia | Dennis Yeow | | Neuroscience Research Australia | Sydney | New South Wales | Nothing to declare |
| Australia | Carolyn Sue | | Neuroscience Research Australia | Sydney | New South Wales | Nothing to declare |
| Australia | Victor Flores Ocampo | | QIMR Berghofer Medical Research Institute | Brisbane | Queensland | Nothing to declare |
| Australia | Christine Wools | | Epworth hospital | Melbourne | Victoria | Nothing to declare |
| Australia | Keren Aliza Weiss | | Garvan Institute of Medical Research | Sydney | New South Wales | Nothing to declare |
| Australia | Ryan L Davis | | University of Sydney | Sydney | New South Wales | Nothing to declare |
| Australia | Amanda Willis | | Garvan Institute of Medical Research | Sydney | New South Wales | Nothing to declare |
| Australia | Steven He | | Garvan Institute of Medical Research | Sydney | New South Wales | Nothing to declare |
| Australia | Robert Arthur Wilcox | | Flinders Medical Centre | Bedford Park | South Australia | Nothing to declare |
| Australia | Denise Howting | | Perron Institute for Neurological and Translational Science | Nedlands | Western Australia | Nothing to declare |
| Australia | Jack David Price | | Perron Institute | Perth | Western Australia | Nothing to declare |
| Australia | Pak Leng Cheong | | Sydney Local Health District | Sydney | New South Wales | Nothing to declare |
| Australia | Michel Tchan | | Westmead Hospital | Westmead | New South Wales | Nothing to declare |
| Australia | Mary-Anne Young | | MonoPD | Sydney | New South Wales | Nothing to declare |
| Australia | Catriona Mclean | | Florey neuroscience | Melbourne | Victoria | Nothing to declare |
| Australia | Nicholas G. Martin | | QIMR Berghofer Medical Research Institute | Brisbane | Queensland | Nothing to declare |
| Australia | Hugo Morales Briceño | | Westmead Hospital | Sydney | New South Wales | Nothing to declare |
| Australia | Thomas Kimber | | Central Adelaide Local Health Network | Adelaide | South Australia | Nothing to declare |
| Australia | Kathy H. C. Wu | | St Vincent's Hospital Sydney | Darlinghurst | New South Wales | Nothing to declare |
| Australia | John O'Sullivan | | University of Queensland | Brisbane | Queensland | Nothing to declare |
| Australia | Lewis M Singleton | | Perron Institute of Neurological and Translational Science | Perth | Western Australia | Nothing to declare |
| Australia | Laura Ivete Rudaks | | Concord Repatriation General Hospital | Sydney | New South Wales | Nothing to declare |
| Australia | Luis M. García-Marín | | Queensland Institute of Medical Research (QIMR Berghofer) | Brisbane | Queensland | Nothing to declare |
| Australia | Amanda Lim | | Queensland Institute of Medical Research (QIMR Berghofer) | Brisbane | Queensland | Nothing to declare |
| Australia | Sue-Faye Siow | | Garvan Institute of Medical Research | Sydney | Wales | Nothing to declare |
| Australia | Natalia Soledad Ogonowski | | QIMR Berghofer | Brisbane | Queensland | Nothing to declare |
| Australia | Santiago Diaz Torres | | Queensland Institute of Medical Research (QIMR Berghofer) / The University of Queensland | Brisbane | Queensland | Nothing to declare |
| Australia | Fangyuan Cao | | QIMR Berghofer | Brisbane | Queensland | Nothing to declare |
| Austria | Alexander Zimprich | | Medical University Vienna Austria | Vienna | | Nothing to declare |
| Azerbaijan | Kanan Jafarov | | Istanbul Klinik | Baku | | Nothing to declare |
| Bangladesh | Imran Sarker | | National Institute of Neurosciences and Hospital | Dhaka | | Nothing to declare |
| Belgium | David Crosiers | | University of Antwerp | Antwerp | | Nothing to declare |
| Bolivia | Erick Gonzalez Delgado | | MedicalCenter Neurocenter S.A. | Santa Cruz de la Sierra | | Nothing to declare |
| Brazil | Artur F. Schumacher-Schuh | | Universidade Federal do Rio Grande do Sul / Hospital de Clínicas de Porto Alegre | Porto Alegre | | Nothing to declare |
| Brazil | Carlos Rieder | | Federal University of Health Sciences of Porto Alegre | Porto Alegre | | Nothing to declare |
| Brazil | Paula Saffie Awad | | Universidade Federal do Rio Grande do Sul | Porto Alegre | | Global Parkinson's Genetics Program |
| Brazil | Vitor Tumas | | University of São Paulo | São Paulo | | Nothing to declare |
| Brazil | Sarah Camargos | | Universidade Federal de Minas Gerais | Belo Horizonte | | Nothing to declare |
| Brazil | Lucas Faria Costa | | Universidade Federal de Minas Gerais | Belo Horizonte | | Nothing to declare |
| Brazil | Pedro Braga Neto | | Federal University of Ceará | Fortaleza | | Nothing to declare |
| Brazil | Juan Sebastián Sánchez León | | Hospital de Clínicas de Porto Alegre | Porto Alegre | | Nothing to declare |
| Brazil | Maira Rozenfeld Olchik | | Hospital de Clínicas de Porto Alegre (HCPA) | Porto Alegre | | Nothing to declare |
| Brazil | Bruno Lopes Santos-Lobato | | Hospital Ophir Loyola | Belém | | Nothing to declare |
| Brazil | Grace Helena Letro | | Hospital da Pontificia Universiade Católica de Campinas ( Hospital PUC-Campinas | Campinas | | Nothing to declare |
| Brazil | Daniel Teixeira-dos-Santos | | Hospital de Clinicas de Porto Alegre | Porto Alegre | | Nothing to declare |
| Brazil | Henrique Ballalai Ferraz | | Universidade Federal de São Paulo (UNIFESP/EPM) | São Paulo | | Nothing to declare |
| Brazil | Carolina Candeias da Silva | | Universidade Federal de São Paulo UNIFESP/EPM | São Paulo | | Nothing to declare |
| Brazil | Vanderci Borges | | Universidade Federal de São Paulo UNIFESP/EPM | São Paulo | | Michael G. Fox Foundation and Fundacão de Apoio à Pesquisa -FAP UNIFESP |
| Brazil | Dayany Leonel Boone | | Universidade Federal de São Paulo - UNIFESP/EPM | São Paulo | | Nothing to declare |
| Brazil | Mariana Cavalcanti Costa | | Universidade Federal de São Paulo - UNIFESP/EPM | São Paulo | | Nothing to declare |
| Canada | Oury Monchi | | Institut universitaire de gériatrie de Montréal | Montreal | Quebec | CIHR, Brain Canada, Parkinson Canada |
| Canada | Edward Fon | | McGill University | Montreal | Quebec | Nothing to declare |
| Canada | Robert Thibault | | Aligning Science Across Parkinson's | Vancouver | British Columbia | Nothing to declare |
| Canada | Ziv Gan-Or | | McGill University | Montreal | Quebec | Nothing to declare |
| Canada | Meron Teferra | | McGill University | Montreal | Quebec | Nothing to declare |
| Canada | Anthony Lang | | University of Toronto | Toronto | Ontario | Nothing to declare |
| Canada | Konstantin Senkevich | | McGill University | Montreal | Quebec | Nothing to declare |
| Chile | Marcelo Miranda | | Departament of Neurology, Clinica MEDS, Santiago , Chile . Fundación Diagnosis , Santiago , Chile | Santiago | | Nothing to declare |
| Chile | María Leonor Bustamante | | Faculty of Medicine Universidad de Chile | Santiago | | Nothing to declare |
| Chile | Juan Cristobal Nuñez | | Universidad de Chile - Clínica Alemana Santiago | Santiago | | Nothing to declare |
| Chile | Boris Lucero | | Universidad Católica del Maule | Talca | | Nothing to declare |
| Chile | Alicia Colombo | | University of Chile | Santiago | | Nothing to declare |
| Chile | Maria Teresa Muñoz Personal | | Universidad de Chile | Santiago | | Internal Research Fund – Occupational pesticide exposure and Parkinson's disease risk in older Chilean adults from the LARGE-PD cohort, School of Public Health, Universidad de Chile.<br><br>FONDECYT Regular 1240899 – Environmental exposure to pesticides and neurobehavioral and neurophysiological effects in adults from rural communities in the Maule Region, Chile |
| Chile | Eduardo Pérez Palma | | Universidad del Desarrollo | Santiago | | Nothing to declare |
| Chile | Pedro Chana-Cuevas | | Universidad de Santiago de Chile | Santiago | | Nothing to declare |
| Chile | Ana Belen Miranda Cortes | | Fundación Diagnosis | Santiago | | Nothing to declare |
| Chile | María Eugenia Contreras Pinto | | Hospital San Juan de Dios | La serena | | Nothing to declare |
| Chile | Francisca Canals | | Inmov | Santiago | | Nothing to declare |
| Chile | Patricio Alejandro Olguín Aguilera | | Universidad de Chile. Facultad de Medicina | Santiago | | Nothing to declare |
| Chile | Elias Fernandez-Toledo | | University of Concepción | Concepcion | | Nothing to declare |
| Chile | Benjamin Pizarro Gallequillos | | Centro de Imagenología, Hospital Clínico Universidad de Chile | Santiago | | Nothing to declare |
| Chile | Carlos Hernandez | | Universidad del Desarrollo | Santiago | | Nothing to declare |
| Chile | Floria C. Pancetti | | Universidad Católica del Norte | Coquimbo | | Nothing to declare |
| Chile | Juan Cristobal Nuñez Fuster | | Hospital Clínico Unversidad de Chile - Clínica Alemana Santiago | Santiago | | Nothing to declare |
| Chile | José Manuel Fernández | | INMOV | Buenos Aires | | Nothing to declare |
| Chile | Lorena Hudson Aros | | INMOV (Instituto Nacional de Movimientos Anormales) | Buenos Aires | | Nothing to declare |
| China | Beisha Tang | | Central South University | Changsha | | Nothing to declare |
| China | Huifang Shang | | West China Hospital Sichuan University | Chengdu | | Nothing to declare |
| China | Jifeng Guo | | Xiangya Hospital | Changsha | | Nothing to declare |
| China | Piu Chan | | Capital Medical University | Beijing | | Nothing to declare |
| China | Wei Luo | | Zhejiang University | Hangzhou | | Nothing to declare |
| China | Zhenhua Liu | | Xiangya Hospital, Central South University | Changsha | | Nothing to declare |
| China | Germaine Hiu-Fai Chan | | Queen Elizabeth Hospital | Kowloon | | Nothing to declare |
| China | Nancy Ip | | The Hong Kong University of Science and Technology | Kowloon | | Nothing to declare |
| China | Nelson Yuk-Fai Cheung | | Queen Elizabeth Hospital | Kowloon | | Nothing to declare |
| China | Phillip Chan | | The Hong Kong University of Science and Technology | Kowloon | | Nothing to declare |
| China | Xiaopu Zhou | | The Hong Kong University of Science and Technology | Kowloon | | Nothing to declare |
| Colombia | Gonzalo Arboleda | | Universidad Nacional de Colombia | Bogotá | | Nothing to declare |
| Colombia | Jorge Orozco | | Fundación Valle del Lili | Santiago De Cali | | Nothing to declare |
| Colombia | David Antonio Pineda-Salazar | | GRUPO DE NEUROCIENCIAS DE ANTIOQUIA (GNA) | Medellín | | Nothing to declare |

| | | | | | | |
|---|---|---|---|---|---|---|
| **Colombia** | Beatriz Munoz Ospina | | Fundacion Valle del Lili | Santiago De Cali | | Nothing to declare |
| **Colombia** | Tatiana Lopez-Gonzalez | | Universidad Nacional de Colombia | Bogotá | | Nothing to declare |
| **Colombia** | Carlos Velez-Pardo | | Universidad de Antioquia | Medellín | | Nothing to declare |
| **Colombia** | Marlene Jimenez-Del-Rio | | Universidad de Antioquia | Medellín | | Nothing to declare |
| **Colombia** | Sonia Moreno Masmela | | Universidad de Antioquia | Medellín | | Nothing to declare |
| **Colombia** | Valentina Quintana Pena | | Icesi University | Cali | | Nothing to declare |
| **Colombia** | David Fernando Aguillon Niño | | University of Antioquia | Medellín | | Nothing to declare |
| **Colombia** | Eliana Pineda Mateus | | Universidad del Rosario | Bogotá | | Nothing to declare |
| **Colombia** | Karen Lizzette Velasquez Mendez | | Universidad Nacional de Colombia | Bogotá | | Nothing to declare |
| **Costa Rica** | Alvaro Hernandez | | University of Costa Rica | San Jose | | Nothing to declare |
| **Costa Rica** | Jaime Fornaguera Trías | | Universidad de Costa Rica | San José | | Nothing to declare |
| **Costa Rica** | Roger Rodriguez Monge | | Universidad de Costa Rica | San José | | Nothing to declare |
| **Denmark** | Per Borghammer | | Aarhus University | Aarhus | | Nothing to declare |
| **Dominican Republic** | Rossy Cruz Vicioso | | Unión Médica Del Norte, Clínica Universitaria | Santiago de los Caballeros | | Nothing to declare |
| **Dominican Republic** | Alpher Perez Arias | | Clinica Union Medica del Norte | Santiago de los Caballeros | | Nothing to declare |
| **Dominican Republic** | Ernestina Castro Salazar | | Unión Medica del Norte | Santiago de los Caballeros | | Nothing to declare |
| **Ecuador** | Carlos Rodriguez-Alarcon | | Interlab | Guayaquil | | Nothing to declare |
| **Egypt** | Mohamed Salama | | The American University in Cairo | Cairo | | The AUC/ ASRT/ DAAD |
| **Egypt** | Ali Shalash | | Ain Shams University Hospital | Cairo | | TRAPCAF |
| **El Salvador** | Tatiana Ascencio | | Dr. Andres Bello university | San Salvador | | Nothing to declare |
| **El Salvador** | Oscar Peña-Rodas | | Universidad Dr Andrés Bello | San Salvador | | Nothing to declare |
| **El Salvador** | Susana Lissette Peña Martínez | | UNAB | San Salvador | | Nothing to declare |
| **Ethiopia** | Yared Z. Zewde | | Addis Ababa University | Addis Ababa | | Nothing to declare |
| **France** | Alexis Brice | | Paris Brain Institute | Paris | | Nothing to declare |
| **France** | Jean-Christophe Corvol | | Sorbonne Université | Paris | | Nothing to declare |
| **France** | Mari Vidailhet | | Salpêtriere Hospital (AP-HP), Sorbonne Université | Paris | | Nothing to declare |
| **France** | Mathieu Anheim | | University Hospital of Strasbourg, Strasbourg, France | STRASBOURG | | Nothing to declare |
| **France** | Yves Agid | | Paris Brain Institute | Paris | | Nothing to declare |
| **France** | Louise-Laure Mariani | | Paris Brain Institute - Sorbonne University | Paris | | Nothing to declare |
| **France** | Alexandra Durr | | Paris Brain Institute | Paris | | Nothing to declare |
| **France** | Rascol | | université Toulouse | toulouse | | Nothing to declare |
| **France** | Ory Magne Fabienne | | chu toulouse | toulouse | | Nothing to declare |
| **France** | Suzanne Lesage | | Paris Brain Institute (ICM) | Paris | | Nothing to declare |
| **France** | Defebvre Luc | | CHU Lille | Lille | | Nothing to declare |
| **France** | Tesson Christelle | | Institut du Cerveau-Paris Brain Institute-ICM | Paris | | Nothing to declare |
| **France** | Philippe Damier | | Nantes Université | Nantes | | Nothing to declare |
| **France** | Francois Tison | | University of Bordeaux, France | Bordeaux | | Nothing to declare |
| **France** | Stéphane Thobois | | Hospices civils de Lyon, Hopital Neurologique Pierre Wertheimer | BRON | | Nothing to declare |
| **France** | Jean-Luc Houeto | | Limoges University Hospital | Limoges | | Nothing to declare |
| **France** | Brefel Courbon Christine | | CHU Toulouse | TOULOUSE | | Nothing to declare |
| **France** | Sara Sambin | | CIC Neurosciences ,Paris Brain institute | Paris | | Nothing to declare |
| **France** | Aymeric Lanore | | Paris Brain Institute | Paris | | Nothing to declare |
| **France** | NS-PARK Consortium | / | / | / | | / |
| **France** | Isabelle Arnulf | | Hopital Universitaire Pitie Salpetriere, AP-HP | Paris | | Nothing to declare |
| **France** | Stéphane Lehericy | | Hopital Universitaire Pitie Salpetriere, AP-HP | Paris | | Nothing to declare |
| **France** | Graziella Mangone | | Hopital Universitaire Pitie Salpetriere, AP-HP | Paris | | Nothing to declare |
| **France** | Poornima Menon | | Hopital Universitaire Pitie Salpetriere, AP-HP | Paris | | Nothing to declare |
| **France** | David Grabli | | Hopital Universitaire Pitie Salpetriere, AP-HP | Paris | | Nothing to declare |
| **France** | Florence Cormier-Dequaire | | Hopital Universitaire Pitie Salpetriere, AP-HP | Paris | | Nothing to declare |
| **France** | Bertrand Degos | | Avicenne Hospital | Bobigny | | Nothing to declare |
| **France** | Marie-Alexandrine | | Hopital Universitaire Pitie Salpetriere, AP-HP | Paris | | Nothing to declare |
| **France** | Eve Benchetrit | | La Timone Hospital | Marseille | | Nothing to declare |
| **France** | Anais Raud | | Centre de réadaptation de l'Estuaire & Résidence les jardines de la chenaie | Le Havre | | Nothing to declare |
| **France** | Virginie Czernecki | | Hopital Universitaire Pitie Salpetriere, AP-HP | Paris | | Nothing to declare |
| **France** | Pierre Pouget | | ICM | Paris | | Nothing to declare |
| **France** | Tymothée Poitou | | ICM | Paris | | Nothing to declare |
| **France** | Elodie Hainque | | Hopital Universitaire Pitie Salpetriere, AP-HP | Paris | | Nothing to declare |
| **France** | Smaranda Leu Semenescu | | Hopital Universitaire Pitie Salpetriere, AP-HP | Paris | | Nothing to declare |
| **France** | Pauline Dodet | | Hopital Universitaire Pitie Salpetriere, AP-HP | Paris | | Nothing to declare |
| **France** | Samir Bekadar | | Eli Lilly and Company | Paris | | Nothing to declare |
| **France** | Fanny Mochel | | Hopital Universitaire Pitie Salpetriere, AP-HP | Paris | | Nothing to declare |
| **France** | Farid Ichou | | Hopital Universitaire Pitie Salpetriere, AP-HP | Paris | | Nothing to declare |
| **France** | Rahul Gaurav | | Hopital Universitaire Pitie Salpetriere, AP-HP | Paris | | Nothing to declare |
| **France** | Nadya Pyatigorskaya | | Hopital Universitaire Pitie Salpetriere, AP-HP | Paris | | Nothing to declare |
| **France** | Romain Valabregue | | ICM | Paris | | Nothing to declare |
| **France** | Cécile Galléa | | ICM | Paris | | Nothing to declare |
| **France** | Marie-Odile Habert | | Hopital Universitaire Pitie Salpetriere, AP-HP | Paris | | Nothing to declare |
| **France** | Dijana Petrovska | | Telecom Sud Paris, Evry | Évry | | Nothing to declare |
| **France** | Laetitia Jeancolas | | Telecom Sud Paris, Evry / ICM | Évry | | Nothing to declare |
| **France** | Manon Gomes | | Hopital Universitaire Pitie Salpetriere, AP-HP | Paris | | Nothing to declare |
| **France** | Stephanie Carvalho | | Hopital Universitaire Pitie Salpetriere, AP-HP | Paris | | Nothing to declare |
| **France** | Alizé Chalançon | | Hopital Universitaire Pitie Salpetriere, AP-HP | Paris | | Nothing to declare |
| **France** | Carole Dongmo-Kenfack | | Hopital Universitaire Pitie Salpetriere, AP-HP | Paris | | Nothing to declare |
| **France** | Mickael Le | | Hopital Universitaire Pitie Salpetriere, AP-HP | Paris | | Nothing to declare |
| **France** | Amelie Bernardo | | Hopital Universitaire Pitie Salpetriere, AP-HP | Paris | | Nothing to declare |
| **France** | Avigaelle Abitbol | | Hopital Universitaire Pitie Salpetriere, AP-HP | Paris | | Nothing to declare |
| **France** | Marion Houot | | Hopital Universitaire Pitie Salpetriere, AP-HP | Paris | | Nothing to declare |
| **France** | Sylvie Forlani | | Hopital Universitaire Pitie Salpetriere, AP-HP | Paris | | Nothing to declare |
| **France** | Sophia Loiodice | | Hopital Universitaire Pitie Salpetriere, AP-HP | Paris | | Nothing to declare |
| **France** | Ludmila Jornea | | Hopital Universitaire Pitie Salpetriere, AP-HP | Paris | | Nothing to declare |
| **Georgia** | Mariam Kekenadze | | Tbilisi State Medical University | Tbilisi | | Nothing to declare |
| **Georgia** | Irine Khatiashvili | | S. Khechinashvili University Hospital | Tbilisi | | Nothing to declare |
| **Georgia** | Maia Beridze | | Tbilisi State Medical University | Tbilisi | | Nothing to declare |
| **Georgia** | Sophia Sopromadze | | Ivane Javakhishvili Tbilisi State University | Tbilisi | | Nothing to declare |
| **Georgia** | Irine Khatiashvilli | | Ivane Javakhishvili Tbilisi State University | Tbilisi | | Nothing to declare |
| **Georgia** | Mariam Mshvenieradze | | Ivane Javakhishvili Tbilisi State University | Tbilisi | | Nothing to declare |
| **Georgia** | Marika Megrelishvili | | Ilia State University | Tbilisi | | Nothing to declare |
| **Georgia** | Alexander Tsiskaridze | | Ivane Javakhishvili Tbilisi State University | Tbilisi | | Nothing to declare |
| **Germany** | Ana Westenberger | | University of Lübeck | Lübeck | | Nothing to declare |
| **Germany** | Anastasia Illarionova | | Deutsches Zentrum für Neurodegenerative Erkrankungen | Göttingen | | Nothing to declare |
| **Germany** | Brit Mollenhauer | | University Medical Center Göttingen | Göttingen | | Nothing to declare |
| **Germany** | Christine Klein | | University of Lübeck | Lübeck | | CK serves as a medical Advisor to Centogene on genetic testing reports in the field of movement disorders, except Parkinson's disease, and is a member of the Scientific Advisory Board of Retromer Therapeutics |
| **Germany** | Eva-Juliane Vollstedt | | University of Lübeck | Lübeck | | Nothing to declare |
| **Germany** | Franziska Hopfner | | Department of Neurology, University Hospital, LMU Munich | Munich | | Nothing to declare |
| **Germany** | Günter Höglinger | | Department of Neurology, University Hospital, LMU Munich | Munich | | Nothing to declare |
| **Germany** | Harutyun Madoev | | University of Lübeck | Lübeck | | Nothing to declare |
| **Germany** | Joanne Trinh | | University of Lübeck | Lübeck | | Nothing to declare |
| **Germany** | Katja Lohmann | | University of Lübeck | Lübeck | | Nothing to declare |
| **Germany** | Manu Sharma | | University of Tubingen | Tübingen | | Dr. Sharma is further funded by the Michael J Fox Foundation, USA Genetic Diversity in PD Program: GAP-India Grant ID: 009411. |
| **Germany** | Sergiu Groppa | | University of Mainz | Mainz | | Nothing to declare |
| **Germany** | Thomas Gasser | | University of Tübingen | Tübingen | | Nothing to declare |
| **Germany** | Zih-Hua Fang | | The German Center for Neurodegenerative Diseases | Göttingen | | Nothing to declare |
| **Germany** | Karl Heilbron | | Charité - Universitätsmedizin Berlin | Berlin | | Former employee of 23andMe, Inc. Owns stock and/or stock options in 23andMe, Inc. |
| **Germany** | Wenhua Sun | | University of Teubingen | Tübingen | | Nothing to declare |
| **Germany** | Inke König | | University of Lübeck | Lübeck | | Nothing to declare |
| **Germany** | Daniela Berg | | University Medical Center Schleswig-Holstein | Lübeck | | Nothing to declare |
| **Germany** | Bernhard Haslinger | | Technical University of Munich | Munich | | Nothing to declare |
| **Germany** | Teresa Kleinz | | University of Lübeck | Lübeck | | Nothing to declare |
| **Germany** | Norbert Brüggemann | | University of Lübeck | Lübeck | | Dr. Brüggemann received honaria from Abbvie, Esteve, Ipsen, Merz, Teva and Zambon |
| **Germany** | Konstantin Kufer | | German Centre for Neurodegenerative Diseases (DZNE) / University Hospital Bonn | Bonn | | Nothing to declare |
| **Germany** | Antonia Maria Buchal | | University Hospital Bonn | Bonn | | Nothing to declare |
| **Germany** | Matthias Höllerhage | | Hannover Medical School | Hannover | | Nothing to declare |
| **Germany** | Florian Wegner | | Hannover Medical School | Hannover | | Nothing to declare |
| **Germany** | Nils Schroeter | | UKS, University of Saarland | Homburg and Mainz | | Nothing to declare |
| **Germany** | Kathrin Brockmann | | University of Tübingen | Tübingen | | KB has received research funding from the Michael J. Fox Foundation for Parkinson's Research, the German Society for Parkinson DPG, the Health Forum Baden Wuerttemberg, the Else Kröner Fresenius Stiftung, the University of Tuebingen, and from the German Research Foundation DFG. KB is a consultant for F. Hoffmann-La Roche Ltd., Vanqua Bio, and the Michael J. Fox Foundation for Parkinson's Research and has received speaker honoraria from Abbvie, Lundbeck, UCB and Zambon. |
| **Germany** | Isabel Wurster | | University of Tübingen | Tübingen | | Nothing to declare |
| **Germany** | Theresa Lüth | | University of Lübeck | Lübeck | | Nothing to declare |
| **Germany** | Christian Beetz | | CENTOGENE | Rostock | | Nothing to declare |
| **Germany** | Krishnakumar Kandaswamy | | Centogene GmBH | Berlin | | Nothing to declare |
| **Germany** | Eva Schäffer | | Kiel University | Kiel | | Nothing to declare |
| **Germany** | Kirsten Zeuner | | Kiel University | Kiel | | Nothing to declare |
| **Germany** | Gregor Kuhlenbäumer | | Kiel University | Kiel | | Nothing to declare |
| **Germany** | Peter Bauer | | Centogene GmbH | Rostock | | Nothing to declare |
| **Germany** | Martin Klietz | | Hannover Medical School | Hannover | | Nothing to declare |
| **Germany** | Carolin Gabbert | | University of Lübeck | Lübeck | | Nothing to declare |
| **Germany** | Alexander Balck | | University of Lübeck | Lübeck | | Nothing to declare |
| **Germany** | Christoph Westenberger | | University of Lübeck | Lübeck | | Nothing to declare |
| **Germany** | Claudia Schulte | | University of Tübingen | Tübingen | | Nothing to declare |
| **Germany** | Sebastian Schade | | Paracelsus-Elena-Klinik Kassel | Kassel | | Nothing to declare |
| **Germany** | Michael Sommerauer | | University Hospital Bonn | Bonn | | Nothing to declare |
| **Germany** | Andre Fienemann | | University of Lübeck | Lübeck | | Nothing to declare |

| | | | | | | |
|---|---|---|---|---|---|---|
| **Germany** | Ann-Kathrin Hauser | | University of Tübingen | Tübingen | | Nothing to declare |
| **Germany** | Claudia Schulte | | University of Tübingen | Tübingen | | Nothing to declare |
| **Germany** | Sana Hrir | | University of Lübeck | Lübeck | | Nothing to declare |
| **Ghana** | Albert Akpalu | | University of Ghana Medical School | Accra | | Nothing to declare |
| **Ghana** | Momodou Cham | | Richard Novati Catholic Hospital, Catholic Health Service Trust | Accra | | Nothing to declare |
| **Ghana** | Vida Obese | | Kwame Nkrumah University of Science and Technology | Kumasi | | Nothing to declare |
| **Ghana** | Andrew Jacobs Bilson | | Kwame Nkrumah University of Science and Technology | Kumasi | | Nothing to declare |
| **Greece** | Georgia Xiromerisiou | | University of Thessaly | Volos | | Nothing to declare |
| **Greece** | Georgios Hadjigorgiou | | University of Thessaly | Volos | | Nothing to declare |
| **Greece** | Ioannis Dagklis | | Aristotle University of Thessaloniki | Thessaloniki | | Nothing to declare |
| **Greece** | Ioannis Tarnanas | | Ionian University | Corfu | | Nothing to declare |
| **Greece** | Leonidas Stefanis | | Biomedical research Foundation of the Academy of Athens | Athens | | PPMI2 (funded by MJFF), ALAMEDA (H2020 grant), funding by HFRI |
| **Greece** | Maria Stamelou | | Diagnostic and Therapeutic Centre HYGEIA Hospital | Marousi | | The non-profit organisation for scientific research in Parkinson's disease and related disorders |
| **Greece** | Efthymios Dadiotis | | University of Thessaly | Volos | | Nothing to declare |
| **Greece** | Tsamis Konstantinos | | University of Ioannina | Ioannina | | Honoraria and research funding: Sanofi, Genesis, Abbvie, Lavipharm, Medochemie, IPSEN, Bennett, PD Neurotechnology |
| **Greece** | Konitsiotis Spyridon | | University of Ioannina | Ioannina | | Honoraria and research funding: Novartis, Sanofi, BMS, TEVA, Merck, Genesis, Innovis, Viatris, Abbvie, Lundbeck, Angelini, UCB, Lavipharm, Φαρμασερβ, Elpen, Roche, Sandoz, Medtronic, Medison, AstraZeneca, ITF, IPSEN, PD Neurotechnology |
| **Greece** | Iro Boura | | University of Crete | Heraklion | | University of Crete, Changeing-Excellence Hub, AbbVie |
| **Greece** | Makrygianni Mariza | | HYGEIA Hospital | Athens | | Nothing to declare |
| **Greece** | Lina Florentin | | HYGEIA Hospital | Athens | | Nothing to declare |
| **Greece** | Maria Makrygianni | | HYGEIA Hospital | Athens | | Nothing to declare |
| **Greece** | Foivos S. Kanellos | | University of Ioannina | Ioannina | | Nothing to declare |
| **Greece** | Cleanthe Spanaki | | University of Crete | Heraklion | | C.S. has received honoraria for lecturing, advisory fees, educational grants, and travel grants from ITF Hellas, Innovis, Merck Serono, AbbVie, CSL Behring, Roche and Teva. She received research funding from the Greek National Precision Medicine Network, the European Union's Horizon 2020 research innovation program under the Marie Sklodowska-Curie grant agreement No 101007926 and from the European Union's program Excellence Hubs - HORIZON-WIDERA-2022-ACCESS-04-01 under grant agreement No. 101087071. |
| **Honduras** | Alex Medina | | Hospital San Felipe | Tegucigalpa | | Nothing to declare |
| **Honduras** | Evelin Álvarez Herrera | | Universidad Tecnológica Centroamericana (UNITEC) | Tegucigalpa | | Nothing to declare |
| **Honduras** | Heike Hesse Joya | | Universidad Tecnológica Centroamericana UNITEC | Tegucigalpa | | Nothing to declare |
| **Honduras** | Reyna M. Durón | | Universidad Tecnológica Centroamericana | Tegucigalpa | | Nothing to declare |
| **Honduras** | Glenda Oliva Fuentes | | Fundación Lucas para la Salud | Tegucigalpa | | Nothing to declare |
| **Honduras** | Eduardo Jose Ponce Murillo | | Universidad Tecnológica Centroamericana | Tegucigalpa | | Nothing to declare |
| **Iceland** | Kari Stefansson | | deCODE genetics/Amgen Inc., Reykjavik, Iceland Faculty of Medicine, University of Iceland, Reykjavik, Iceland | Reykjavik | | employee of deCODE genetics/Amgen inc |
| **Iceland** | Hreinn Stefansson | | deCODE genetics/Amgen Inc., Reykjavik, Iceland Faculty of Medicine, University of Iceland, Reykjavik, Iceland | Reykjavik | | employee of deCODE genetics/Amgen inc |
| **Iceland** | Vala Palmadottir | | deCODE genetics/Amgen Inc., Reykjavik, Iceland Faculty of Medicine, University of Iceland, Reykjavik, Iceland | Reykjavik | | employee of deCODE genetics/Amgen inc |
| **Iceland** | Astros Th. Skuladottir | | deCODE genetics/Amgen Inc., Reykjavik, Iceland Faculty of Medicine, University of Iceland, Reykjavik, Iceland | Reykjavik | | employee of deCODE genetics/Amgen inc |
| **India** | Asha Kishore | | Aster Medcity | Kochi | | Michael J. Fox Foundation |
| **India** | Divya Kp | | Sree Chitra Tirunal Institute for Medical Sciences and Technology | Thiruvananthapuram | | Nothing to declare |
| **India** | Pramod Pal | | National Institute of Mental Health & Neurosciences | Bengaluru | | Nothing to declare |
| **India** | Prashanth Lingappa Kukkle | | Manipal Hospital | Delhi | | Nothing to declare |
| **India** | Roopa Rajan | | All India Institute of Medical Sciences | Delhi | | Nothing to declare |
| **India** | Rupam Borgohain | | Nizam's Institute Of Medical Sciences | Hyderabad | | Nothing to declare |
| **Iran** | Mehri Salari | | Shahid Beheshti University of Medical Science | Tehran | | Nothing to declare |
| **Israel** | Tamara Shiner | | Tel Aviv Sourasky Medical Center | Tel Aviv-Yafo | | Nothing to declare |
| **Israel** | Avner Thaler | | Tel Aviv Sourasky Medical Center | Tel Aviv-Yafo | | Nothing to declare |
| **Israel** | Noa Bregman | | Tel Aviv Medical Center | Tel Aviv-Yafo | | Nothing to declare |
| **Italy** | Andrea Quattrone | | Magna Græcia University of Catanzaro | Catanzaro | | Nothing to declare |
| **Italy** | Enza Maria Valente | | University of Pavia | Pavia | | Nothing to declare |
| **Italy** | Grazia Annesi | | National Research Council | Cosenza | | Nothing to declare |
| **Italy** | Lucilla Parnetti | | University of Perugia | Perugia | | Nothing to declare |
| **Italy** | Micol Avenali | | University of Pavia | Pavia | | Nothing to declare |
| **Italy** | Monica Gagliardi | | Magna Graecia University | Catanzaro | | Nothing to declare |
| **Italy** | Tommaso Schirinzi | | University of Rome Tor Vergata | Rome | | Nothing to declare |
| **Italy** | Caterina Galandra | | IRCCS Mondino Foundation | Pavia | | Nothing to declare |
| **Italy** | Anna De Rosa | | University of Naples Federico II | Naples | | Nothing to declare |
| **Italy** | Rosangela Ferese | | IRCCS Neuromed | Pozzilli | | Nothing to declare |
| **Italy** | Jolanda Buonocore | | Magna Graecia University | Catanzaro | | Nothing to declare |
| **Italy** | Radha Procopio | | Magna Graecia University | Catanzaro | | Nothing to declare |
| **Italy** | Ilaria Palmieri | | IRCCS Mondino Foundation | Pavia | | Nothing to declare |
| **Italy** | Michele Terzaghi | | University of Pavia | Pavia | | Nothing to declare |
| **Italy** | Paola Dimartino | | University of Pavia | Pavia | | Nothing to declare |
| **Italy** | Roberta Bovenzi | | University of Tor Vergata | Rome | | Nothing to declare |
| **Italy** | Maxine Paige-Pritchett | | Università di Pavia | Pavia | | Nothing to declare |
| **Japan** | Manabu Funayama | | Juntendo University | Tokyo | | Nothing to declare |
| **Japan** | Nobutaka Hattori | | Juntendo University faculty of medicine | Tokyo | | Nothing to declare |
| **Japan** | Tomotaka Shiraishi | | Jikei University School of Medicine | Tokyo | | Nothing to declare |
| **Japan** | Kensuke Daida | | Juntendo University | Bunkyō | | Nothing to declare |
| **Kazakhstan** | Altynay Karimova | | Institute of Neurology and Neurorehabilitation | Almaty | | Nothing to declare |
| **Kazakhstan** | Gulnaz Kaishibayeva | | Institute of Neurology and Neurorehabilitation | Almaty | | Nothing to declare |
| **Kazakhstan** | Aigerim Utegenova | | West Kazakhstan Marat Ospanov State Medical University | Aktobe | | Nothing to declare |
| **Kazakhstan** | Vadim Akhmetzhanov | | Medline medical center | Astana | | Nothing to declare |
| **Kazakhstan** | Seitzhan Aidarov | | National Center for Neurosurgery | Astana | | Nothing to declare |
| **Kazakhstan** | Tautanova Raushan | | Astana Medical University | Astana | | Nothing to declare |
| **Kazakhstan** | Dinara Alzhanova | | Astana Medical University | Astana | | Nothing to declare |
| **Kazakhstan** | Zhanybek Myrzayev | | International University of Postgraduate Education | Almaty | | Nothing to declare |
| **Kazakhstan** | Saltanat Abdraimova | | South Kazakhstan Medical Academy | Shymkent | | Nothing to declare |
| **Kazakhstan** | Nazira Zharkinbekova | | South Kazakhstan Medical Academy | Shymkent | | South Kazakhstan Medical academy |
| **Kazakhstan** | Chingiz Shashkin | | International Research Institute of Postgraduate Education | Almaty | | Nothing to declare |
| **Kazakhstan** | Guzel Shiderova | | Institute of Neurology and Neurorehabilitation | Almaty | | Nothing to declare |
| **Kazakhstan** | Bagzhan Syzdykova | | Astana Medical University | Astana | | Nothing to declare |
| **Kazakhstan** | Aigul. P. Yermagambetova | | West Kazakhstan Marat Ospanov Medical University | Aktobe | | Nothing to declare |
| **Kazakhstan** | Alima A. Khamidulla | | West Kazakhstan Marat Ospanov Medical University | Aktobe | | Nothing to declare |
| **Kazakhstan** | Zhanylsyn U.Urasheva | | West Kazakhstan Marat Ospanov Medical University | Aktobe | | Nothing to declare |
| **Kazakhstan** | Gulnar B. Kabdrakhmanova | | West Kazakhstan Marat Ospanov Medical University | Aktobe | | Nothing to declare |
| **Kazakhstan** | Talgat Khaibullin | | Semey Medical University | Semey | | Nothing to declare |
| **Kazakhstan** | Altynay Talgatkyzy | | Semey Medical University | Semey | | Nothing to declare |
| **Kazakhstan** | Akper Sagynysh | | Multiple Sclerosis and Autoimmune Neurological Disorders Center at the Multidisciplinary City Hospital No. 1 | Moscow | | Nothing to declare |
| **Kazakhstan** | Sadenova Aigul | | Multidisciplinary City Hospital No. 1 | Moscow | | Nothing to declare |
| **Kenya** | Eunice Nyambane | | Aga Khan University Nairobi | Nairobi | | Nothing to declare |
| **Kyrgyzstan** | Cholpon Shambetova | | Kyrgyz State Medical Academy | Bishkek | | Nothing to declare |
| **Kyrgyzstan** | Bermet Nurbekova | | I.K. Akhunbaev Kyrgyz State Medical Academy (KSMA) | Bishkek | | Nothing to declare |
| **Kyrgyzstan** | Nurayim Beishembieva | | I.K. Akhunbaev Kyrgyz State Medical Academy (KSMA) | Bishkek | | Nothing to declare |
| **Kyrgyzstan** | Nargiza Atambekova | | I.K. Akhunbaev Kyrgyz State Medical Academy (KSMA), Clinical Hospital of Emergency Medical Care | Bishkek | | Nothing to declare |
| **Kyrgyzstan** | Zhumagul Osmonova | | Kyrgyz State Medical Institute of postgraduate training and continuous education named after S.B. Daniyarov | Bishkek | | Nothing to declare |
| **Kyrgyzstan** | Maatali Abdimanap uulu | | Osh State University, Osh City Clinical Hospital | Osh | | Nothing to declare |
| **Kyrgyzstan** | Aisuikum Abdumalikova | | I.K. Akhunbaev Kyrgyz State Medical Academy (KSMA), Osh City Clinical Hospital | Osh | | Nothing to declare |
| **Kyrgyzstan** | Adylbek Iusupov | | I.K. Akhunbaev Kyrgyz State Medical Academy (KSMA), Osh City Clinical Hospital | Osh | | Nothing to declare |
| **Kyrgyzstan** | Aliia Mukhanova | | International Higher School of Medicine | Bishkek | | Nothing to declare |
| **Kyrgyzstan** | Zhyldyz Imanalieva | | I.K. Akhunbaev Kyrgyz State Medical Academy (KSMA), "Aqua Lab" LLC | Bishkek | | Nothing to declare |
| **Kyrgyzstan** | Gulmira Zhamilova | | Clinical Hospital of the Presidential Administration of the Kyrgyz Republic | Bishkek | | Nothing to declare |
| **Kyrgyzstan** | Shokhista Artyshova | | Clinical Hospital of the Presidential Administration of the Kyrgyz Republic | Bishkek | | Nothing to declare |
| **Kyrgyzstan** | Sabina Baltabaeva | | I.K. Akhunbaev Kyrgyz State Medical Academy (KSMA), Medcenter.kg" clinic | Bishkek | | Nothing to declare |
| **Kyrgyzstan** | Mariia Rekaeva | | I.K. Akhunbaev Kyrgyz State Medical Academy (KSMA), "Unimed" Clinic | Bishkek | | Nothing to declare |
| **Kyrgyzstan** | Aziza Dalbaeva | | National Hospital | Bishkek | | Nothing to declare |
| **Kyrgyzstan** | Anna Khamzina | | I.K. Akhunbaev Kyrgyz State Medical Academy (KSMA), "Unimed" Clinic | Bishkek | | Nothing to declare |
| **Kyrgyzstan** | Saule Temirbaeva | | National Hospital | Bishkek | | Nothing to declare |
| **Kyrgyzstan** | Mukhammadiusuf Abdykadyrov | | Osh State University | Osh | | Nothing to declare |
| **Kyrgyzstan** | Zynan Ruslan uulu | | I.K. Akhunbaev Kyrgyz State Medical Academy (KSMA) | Bishkek | | Nothing to declare |
| **Kyrgyzstan** | Nurbakyt Kadyrov | | Osh City Clinical Hospital | Osh | | Nothing to declare |
| **Kyrgyzstan** | Batma Sattarova | | Batken Regional Hospital | Batken | | Nothing to declare |
| **Kyrgyzstan** | Begimai Sovetbekova | | I.K. Akhunbaev Kyrgyz State Medical Academy (KSMA) | Bishkek | | Nothing to declare |
| **Lebanon** | Fouad Khoury | | Université Saint George - Hôpital national de neurologie et de neurochirurgie | Beirut | | Nothing to declare |
| **Lithuania** | Gintare Baranauskiene | | Vilnius University | Vilnius | | Nothing to declare |
| **Luxembourg** | Rejko Krüger | | University of Luxembourg | Esch-sur-Alzette | | Nothing to declare |
| **Luxembourg** | Patrick May | | University of Luxembourg | Esch-sur-Alzette | | Nothing to declare |
| **Madagascar** | Nomena Rasaholiarison | | Faculty of Medicine, University of Fianarantsoa | Fianarantsoa | | Nothing to declare |
| **Malaysia** | Ai Huey Tan | | University of Malaya | Kuala Lumpur | | Nothing to declare |
| **Malaysia** | Azlina Ahmad-Annuar | | University of Malaya | Kuala Lumpur | | Nothing to declare |
| **Malaysia** | Mohamed Ibrahim Norlinah | | Universiti Kebangsaan Malaysia | Selangor | | Nothing to declare |
| **Malaysia** | Nor Azian Abdul Murad | | UKM Medical Molecular Biology Institute | Kuala Lumpur | | Nothing to declare |
| **Malaysia** | Shahrul Azmin | | Universiti Kebangsaan Malaysia Medical Centre | Kuala Lumpur | | Nothing to declare |
| **Malaysia** | Shen-Yang Lim | | University of Malaya | Kuala Lumpur | | Nothing to declare |
| **Malaysia** | Wael Mohamed | | International Islamic University | Kuala Lumpur | | Nothing to declare |
| **Malaysia** | Yi Wen Tay | | University of Malaya | Kuala Lumpur | | Nothing to declare |
| **Malaysia** | Lim Kai-Shi | | University of Malaya | Kuala Lumpur | | Nothing to declare |
| **Malaysia** | Azalea Tenerife Pajo | | University of Malaya | Kuala Lumpur | | Nothing to declare |
| **Malaysia** | Chia Yuen Kang | | Hospital Queen Elizabeth | Kota Kinabalu | | Nothing to declare |

| | | | | | | |
|---|---|---|---|---|---|---|
| **Malaysia** | Joshua Ooi Chin Ern | | Queen Elizabeth Hospital | Kota Kinabalu | | Nothing to declare |
| **Malaysia** | Khairul Azmi Ibrahim | | HOSPITAL SULTANAH NUR ZAHIRAH KUALA TERENGGANU | KUALA TERENGGANU | | Nothing to declare |
| **Malaysia** | Ahmad Shahir Bin Mawardi | | Hospita Kuala Lumpur | Kuala Lumpur | | Nothing to declare |
| **Malaysia** | Lim Thien Thien | | Island Hospital | Penang | | Nothing to declare |
| **Malaysia** | Tzi Shin Toh | | University of Malaya | Kuala Lumpur | | Nothing to declare |
| **Mexico** | Daniel Martinez-Ramirez | | Tecnologico de Monterrey | Monterrey | | Nothing to declare |
| **Mexico** | Paula Reyes-Pérez | | Universidad Nacional Autónoma de México | Santiago de Querétaro | | Global Parkinson's Genetics Program |
| **Mexico** | Alejandra Medina Rivera | | Universidad Nacional Autónoma de México | Santiago de Querétaro | | Nothing to declare |
| **Mexico** | Nancy Monroy Jaramillo | | Instituto Nacional de Neurología y Neurocirugía | Mexico City | | Nothing to declare |
| **Mexico** | Nadia Alejandra Gandarilla Martinez | | Centro Neurológico del Centro Médico ABC, Campus Santa Fe | Mexico City | | Nothing to declare |
| **Mexico** | Ingrid Estrada-Bellmann | | UNIVERSITY HOSPITAL "DR JOSE E GONZALEZ" | Monterrey | | Nothing to declare |
| **Mexico** | Araliz Puente | | Hospital Ángeles Puebla | Puebla | | Nothing to declare |
| **Mexico** | Ana Paula Angulo Arrieta | | Hospital Ángeles Puebla, Universidad Anáhuac Puebla | Puebla | | Nothing to declare |
| **Mexico** | Eugenia Morelos Figaredo | | ISSSTE Morelia | Morelia | | Nothing to declare |
| **Mexico** | Karla Salinas Barboza | | HOSPITAL GENERAL DE MEXICO | Mexico City | | Nothing to declare |
| **Mexico** | Dante Bernardo Oropeza Canto | | Hospital Angeles de Puebla | Puebla | | Nothing to declare |
| **Mexico** | Mavela Rodríguez-Violante | | Instituto Nacional de Neurología y Neurocirugía | Mexico City | | Nothing to declare |
| **Mexico** | Ana Jimena Hernández-Medrano | | Instituto Nacional de Neurología y Neurocirugía Manuel Velasco Suárez | Mexico City | | Nothing to declare |
| **Mexico** | Amin Cervantes-Arriaga | | Instituto Nacional de Neurología y Neurocirugía | Mexico City | | Nothing to declare |
| **Mexico** | Edith Janeth Gaspar Martínez | | Universidad Nacional Autónoma de México | Santiago de Querétaro | | Nothing to declare |
| **Mexico** | Alejandra E- Ruiz-Contreras | | Universidad Nacional Autonoma de Mexico | Mexico City | | Nothing to declare |
| **Mexico** | Alejandra Lázaro-Figueroa | | National Autonomous University of Mexico | Mexico City | | Nothing to declare |
| **Mexico** | Sarael Alcauter | Blvd. Juriquilla 3001, 76230 | Universidad Nacional Autónoma de México | Mexico City | | Nothing to declare |
| **Mexico** | Edwin Roberto Ramírez-Benítez | | Instituto Nacional de Neurología y Neurocirugía | Mexico City | | Nothing to declare |
| **Mexico** | Karina Cruz-Santillán | | Instituto Nacional de Neurología y Neurocirugía | Mexico City | | Nothing to declare |
| **Mexico** | Rodolfo Solís-Vivanco | | Instituto Nacional de Neurología y Neurocirugía | Mexico City | | Nothing to declare |
| **Mongolia** | Bayasgalan Tserensodnom | | Mongolian National University of Medical Sciences | Ulaanbaatar | | Nothing to declare |
| **Mongolia** | Khosbayar Tulgaa | | Mongolian National University of Medical Sciences | Ulaanbaatar | | Nothing to declare |
| **Mongolia** | Oyujin Ulziibaatar | | Mongolian National University of Medical Sciences | Ulaanbaatar | | Nothing to declare |
| **Morocco** | Ahmed Bouhouche | | Specialties Hospital, CHU Ibn Sina | Rabat | | Nothing to declare |
| **Morocco** | Mossafa Hossain | | Clinique OCEANIC | CASABLANCA | | Nothing to declare |
| **Nepal** | Rajeev Ojha | | Tribhuvan University | Kirtipur | | Nothing to declare |
| **Netherlands** | Wilma Van De Berg | | Vanderbilt University Medical Center | Amsterdam | | Nothing to declare |
| **Netherlands** | Bas Bleom | | Radboud University | Nijmegen | | Nothing to declare |
| **Netherlands** | Bart Van De Warrenburg | | Radboud University Medical Center | Nijmegen | | Nothing to declare |
| **Netherlands** | Lisette Charbonnier | | Brain Research and Innovation Center | Amsterdam | | Nothing to declare |
| **New Zealand** | Tim J. Anderson | | University of Otago | Dunedin | | Heath Research Council of New Zealand; Ministry of Business Innovation and Employment (New Zealand), Neurological Foundation of New Zealand |
| **New Zealand** | Toni L. Pitcher | | University of Otago | Dunedin | | Funding - Health Research Council of New Zealand |
| **New Zealand** | Daniel Jeremy Myall | | New Zealand Brain Research Institution | Christchurch | | Nothing to declare |
| **New Zealand** | John C. Dalrymple-Alford | | University of Canterbury | Christchurch | | Nothing to declare |
| **New Zealand** | Joseph Donnelly | | Te Whatu Ora - Health New Zealand Auckland, New Zealand | Auckland | | Nothing to declare |
| **Nigeria** | Arinola Sanyaolu | | University of Lagos | Lagos | | Nothing to declare |
| **Nigeria** | Njideka Okubadejo | | University of Lagos | Lagos | | Michael J Fox Foundation; Tertiary Education Trust Fund (TETFUND) National Research Fund |
| **Nigeria** | Oluwadamilola Ojo | | College of Medicine of the University of Lagos | Lagos | | Nothing to declare |
| **Nigeria** | Simon Izuchukwu Ozomma | | University of Calabar Teaching Hospital | Calabar | | Nothing to declare |
| **Nigeria** | Kolawole Wahab | | University of Ilorin | Ilorin | | Nothing to declare |
| **Nigeria** | Oladunni Abiodun | | General Hospital | Lagos | | Nothing to declare |
| **Nigeria** | Sani Abubakar | | Ahmadu Bello University | Kaduna State | | Nothing to declare |
| **Nigeria** | Fatimah Abdulai | | University of Abuja Teaching Hospital | Gwagwalada | | Nothing to declare |
| **Nigeria** | Charles Achoru | | Jos University Teaching Hospital | Jos | | Nothing to declare |
| **Nigeria** | Osigwe Agabi | | College of Medicine, University of Lagos | Lagos | | Nothing to declare |
| **Nigeria** | Uchechi Agulanna | | Lagos University Teaching Hospital | Lagos | | Nothing to declare |
| **Nigeria** | Rufus Akinyemi | | Neuroscience and Ageing Research Unit, Institute for Advanced Medical Research and Training, College of | Ibadan | | Nothing to declare |
| **Nigeria** | Wemimo Alaofin | | University of Ilorin | Ilorin | | Nothing to declare |
| **Nigeria** | Ifeyinwa Ani-Osheku | | Asokoro District Hospital | Abuja | | Nothing to declare |
| **Nigeria** | Roosevelt Anyanwu | | College of Medicine, University of Lagos | Lagos | | Nothing to declare |
| **Nigeria** | Cyril Erameh | | Irrua Specialist Teaching Hospital | Ilorin | | Nothing to declare |
| **Nigeria** | Daniel Ezuduemoih | | Lagos University Teaching Hospital | Lagos | | Nothing to declare |
| **Nigeria** | Abdullahi Ibrahim | | Federal University of Health Sciences Teaching Hospital | Azare | | Nothing to declare |
| **Nigeria** | Erica Ikwenu | | Lagos University Teaching Hospital | Lagos | | Nothing to declare |
| **Nigeria** | Frank Imarhiagbe | | University of Benin | Benin City | | Nothing to declare |
| **Nigeria** | Ismaila Ishola | | College of Medicine, University of Lagos | Lagos | | Nothing to declare |
| **Nigeria** | Emmanuel Iwuozo | | Benue State University | Makurdi | | Nothing to declare |
| **Nigeria** | Morenikeji Komolafe | | Obafemi Awolowo University | Ile-Ife | | Nothing to declare |
| **Nigeria** | Alero Nnama | | University of Port Harcourt Teaching Hospital | Port Harcourt | | Nothing to declare |
| **Nigeria** | Paul Nwani | | Nnamdi Azikiwe University Teaching Hospital | Nnewi | | Nothing to declare |
| **Nigeria** | Franscisca Nwaokorie | | College of Medicine, University of Lagos | Lagos | | Nothing to declare |
| **Nigeria** | Ernest Nwazor | | Rivers State University Teaching Hospital | Port Harcourt | | Nothing to declare |
| **Nigeria** | Yahaya Obiabo | | Federal University of Health Sciences | Otukpo | | Nothing to declare |
| **Nigeria** | Nkechi Obianozie | | University of Abuja Teaching Hospital | Gwagwalada | | Nothing to declare |
| **Nigeria** | Olanike Odeniyi | | General Hospital | Lagos | | Nothing to declare |
| **Nigeria** | Francis Odiase | | University of Benin | Benin City | | Nothing to declare |
| **Nigeria** | Ewere Marie Ogbimi | | Delta State University | Abraka | | Nothing to declare |
| **Nigeria** | Adebimpe Ogunmodede | | Federal Medical Center | Owo | | Nothing to declare |
| **Nigeria** | Francis Ojini | | University of Lagos | Lagos | | Nothing to declare |
| **Nigeria** | Rashidat Olanigan | | Lagos State University Teaching Hospital | Ikeja | | Nothing to declare |
| **Nigeria** | Adedunni Olusanya | | College of Medicine, University of Lagos & R-Jolad Hospital | Lagos | | Nothing to declare |
| **Nigeria** | Chiamaka Okereke | | University of Nigeria Teaching Hospital | Ituku-Ozalla | | Nothing to declare |
| **Nigeria** | Gerald Onwuegbuzie | | University of Abuja | Abuja | | Nothing to declare |
| **Nigeria** | Godwin Osaigbovo | | Jos University Teaching Hospital | Jos | | Nothing to declare |
| **Nigeria** | Nosakhare Osemwegie | | University of Port Harcourt | Port Harcourt | | Nothing to declare |
| **Nigeria** | Olajumoke Oshinaike | | Lagos State University College of Medicine | Ikeja | | Nothing to declare |
| **Nigeria** | Lukman Owolabi | | Bayero University Kano | Kano | | Nothing to declare |
| **Nigeria** | Raymond Owolabi | | Federal Medical Center | Owo | | Nothing to declare |
| **Nigeria** | Shyngle Oyakhire | | National Hospital | Abuja | | Nothing to declare |
| **Nigeria** | Fadimatu Sa'Ad | | Federal Teaching Hospital | Gombe | | Nothing to declare |
| **Nigeria** | Funmilola Taiwo | | University College Hospital | Ibadan | | Nothing to declare |
| **Nigeria** | Francisca Nwaokorie | | University of Lagos | Lagos | Lagos | Nothing to declare |
| **Norway** | Lasse Pihlstrøm | | Oslo University Hospital | Oslo | | Southeastern Regional Health Authority, Norway and Michael J. Fox Foundation |
| **Norway** | Manuela Tan | | Oslo University Hospital | Oslo | | Southeastern Norway Regional Health Authority Norway and Michael J. Fox Foundation |
| **Norway** | Ingeborg Haugesag Lie | | Oslo University Hospital | Oslo | | Nothing to declare |
| **Norway** | Jodi Maple-Grødem | | Stavanger University Hospital | Stavanger | | Nothing to declare |
| **Norway** | Solveig E J Dalbro | | Oslo University Hospital | Oslo | | Nothing to declare |
| **Norway** | Ellen Hoven Maurtveten | | Oslo University Hospital | Oslo | | Nothing to declare |
| **Pakistan** | Shoaib Ur-Rehman | | University of Science and Technology Bannu | Bannu | | Nothing to declare |
| **Pakistan** | Mohamed Nour | | Razi Hospital | Rawalpindi | | Nothing to declare |
| **Peru** | Mario Cornejo-Olivas | | Universidad Cientifica del Sur | Lima | | Michael J. Fox Foundation for Parkinson's Research and Aligning Science Across Parkinson's Initiative |
| **Peru** | Nicanor Mori Quispe | | Hospital Nacional Daniel A. Carrión | Lima | | Nothing to declare |
| **Peru** | JULIA ESTHER RIOS PINTO | | UNIVERSIDAD PERUANA LOS ANDES | Huancayo | | Nothing to declare |
| **Peru** | Maryenela Illanes-Manrique | | Instituto Nacional de Ciencias Neurologicas | Lima | | Nothing to declare |
| **Peru** | Cintia Margoth Armas Puente | | Instituto Nacional de Ciencias Neurológicas | Lima | | Nothing to declare |
| **Peru** | Torres Luis | | Instituto Nacional de Ciencias Neurologicas | Lima | | Nothing to declare |
| **Philippines** | Maria Leila Doquenia | | Metropolitan Medical Center | Manila | | Nothing to declare |
| **Philippines** | Raymond Rosales | | Metropolitan Medical Center | Manila | | Nothing to declare |
| **Philippines** | Gerard Saranza | | Chong Hua Hospital | Cebu | | Nothing to declare |
| **Poland** | Agata Gajos | | Medical University of Lodz | Lodz | | Nothing to declare |
| **Russia** | Elena Iakovenko | | Research Center of Neurology | Moscow | | Nothing to declare |
| **Russia** | Anna Gareeva | | Ufa Federal Research Center | Ufa | | Nothing to declare |
| **Russia** | Gulnara Akhmadeeva | | Ufa Scientific Center | Ufa | | Nothing to declare |
| **Russia** | Irina Gilyazova | | Russian Academy of Sciences / Bashkir State Medical University | Ufa | | Nothing to declare |
| **Saudi Arabia** | Bashayer Al Mubarak | | King Faisal Specialist Hospital and Research Center | Riyadh | | Nothing to declare |
| **Saudi Arabia** | Muhammad Umair | | King Abdullah International Medical Research Center | Jeddah | | Nothing to declare |
| **Saudi Arabia** | Nada Abdullah Altassan | | King Faisal Specialist Hospital and Research Center | Riyadh | | Nothing to declare |
| **Serbia** | Nataša Dragašević Mišković | | Neurology Clinic, University Clinical Center of Serbia | Belgrade | | Nothing to declare |
| **Serbia** | Andona Milovanović | | Neurology Clinic, University Clinical Center of Serbia | Belgrade | | Nothing to declare |
| **Singapore** | Eng-King Tan | | National Neuroscience Institute | Singapore | | Singapore National Medical Research Council (MOH-OFLCG-000207) |
| **Singapore** | Jia Nee Foo | | Nanyang Technological University | Singapore | | Singapore National Medical Research Council (MOH-000559) |
| **Singapore** | Elaine Chew | | Nanyang Technological University | Singapore | | Nothing to declare |
| **Slovenia** | Vesna Van Midden | | Ljubljana University Medical Centre | Ljubljana | | Nothing to declare |
| **South Africa** | Ferzana Amod | | University of KwaZulu-Natal | Durban | | Nothing to declare |
| **South Africa** | Jonathan Carr | | University of Stellenbosch | Stellenbosch | | Nothing to declare |
| **South Africa** | Soraya Bardien | | Stellenbosch University | Stellenbosch | | National Research Foundation of South Africa[Grant Number 129249] |
| **South Africa** | Nikita Pillay | | University of the Western Cape | Bellville | | Nothing to declare |
| **South Africa** | Kathryn Step | | Stellenbosch University | Cape Town | | Nothing to declare |
| **South Africa** | Riaan Van Coller | | University of Pretoria | Pretoria | | Nothing to declare |
| **South Korea** | Beomseok Jeon | | Seoul National University Hospital | Seoul | | Nothing to declare |
| **South Korea** | Yun Joong Kim | | Yongin Severance Hospital | Seoul | | Nothing to declare |
| **South Korea** | Jung Hwan Shin | | Seoul National University | Seoul | | Nothing to declare |
| **South Korea** | Joowon Jang | | Seoul National University | Seoul | | Nothing to declare |
| **South Korea** | Jee-Young Lee | | SMG-SNU Boramae Medical Center, College of Medicine Seoul National University | Seoul | | Dr. Lee has received a research grant of National Research Foundation (NRF) funded by the Korean Government (MSIT), a multidisciplinary research grant-in-aid and a focused research grant-in-aid from the Seoul Metropolitan Government Seoul National University (SMG-SNU) Boramae Medical Center, and a research grant from Gemvax, a speaker honorarium from SK chemicals and Bial, a travel support from Ono, and a scientific advisory board of Regeners, Inc. |
| **South Korea** | Ryul Kim | | SMG-SNU Boramae Meidcal Center | Seoul |  | Nothing to declare |
| **South Korea** | HAN-Joon Kim | | Seoul National University Hospital | Seoul | | Nothing to declare |
| **Spain** | Esther Cubo | | Hospital Universitario Burgos | Burgos | | Nothing to declare |
| **Spain** | Ignacio Alvarez | | University Hospital Mutua Terrassa | Barcelona | | Nothing to declare |
| **Spain** | Janet Hoenicka | | Institut de Recerca Sant Joan de Deu | Barcelona | | Fondo de Investigación Sanitaria, Instituto Salud Carlos III, Grant PI019/00126 |

| | | | | | | |
|---|---|---|---|---|---|---|
| **Spain** | Katrin Beyer | | Research Institute Germans Trias i Pujol | Barcelona | | Nothing to declare |
| **Spain** | Maria Teresa Periñan | | Instituto de Biomedicina de Sevilla | Seville | | Nothing to declare |
| **Spain** | Pau Pastor | | University Hospital Germans Trias i Pujol | Barcelona | | Nothing to declare |
| **Spain** | Ruben Fernandez-Santiago | | Hospital Clínic de Barcelona | Barcelona | | Nothing to declare |
| **Spain** | Pilar Gómez Garre | | Instituto de Biomedicina de Sevilla | Seville | | Nothing to declare |
| **Spain** | Pablo Mir | | Instituto de Biomedicina de Sevilla | Seville | | Nothing to declare |
| **Spain** | Mario Ezquerra | | FCRB-IDIBAPS | Barcelona | | Nothing to declare |
| **Spain** | Celia Painous Marti | | Hospital Clinic Barcelona | Barcelona | | Nothing to declare |
| **Spain** | Lola J. Díaz-Feliz | | Fernando Pessoa University, San Roque Hospital | Las Palmas de Gran Canaria | | Nothing to declare |
| **Spain** | José Matías Arbelo González | | Hospital Universitario San Roque Las Palmas/ Universidad Fernando Pessoa Canarias (UFPC) | Las Palmas de Gran Canaria | | Nothing to declare |
| **Spain** | Juan Carlos Martínez Castrillo | | Hospital Ramón y Cajal | Madrid | | Nothing to declare |
| **Spain** | Marina Mata | | Hospital Universitario Infanta Sofia | Madrid | | Nothing to declare |
| **Spain** | Oriol De Fabregues | | Hospital Universitari Vall d'Hebron | Barcelona | | Nothing to declare |
| **Spain** | Lydia Vela-Desojo | | Hospital Universitario Fundación Alcorcón | Madrid | | Nothing to declare |
| **Spain** | Manuel Menendez Gonzalez | | Hospital Universitario Central de Asturias | Oviedo | | Nothing to declare |
| **Spain** | Yaroslau Compta | | IDIBAPS / Hospital Clinic | Barcelona | | Nothing to declare |
| **Spain** | Alicia Garrido | | IDIBAPS-FCRB, Hospital Clinic Barcelona | Barcelona | | Nothing to declare |
| **Spain** | Maria J Marti | | Hospital Clinic de Barcelona, Institut d'Investigacio Biomedica August Pi i Sunyer (IDIBAPS) | Barcelona | | Nothing to declare |
| **Spain** | Almudena Sánchez-Gómez | | Hospital Clinic of Barcelona | Barcelona | | Nothing to declare |
| **Spain** | Alexia T. Sánchez Reyes | | Universidad Fernando Pessoa Canarias | Las Palmas de Gran Canaria | | Nothing to declare |
| **Spain** | Laia Muñoz Llahuna | | IR Sant Pau | Barcelona | | Nothing to declare |
| **Spain** | Joaquim Aumatell Escabies | | IR SANT PAU | Barcelona | | Nothing to declare |
| **Spain** | Javier Pagonabarraga Mora | | IR SANT PAU | Barcelona | | Nothing to declare |
| **Spain** | Ignacio Illán Gala | | IR SANT PAU | Barcelona | | Nothing to declare |
| **Spain** | Esteban Muñoz | | Hospital Clínic de Barcelona | Barcelona | | Nothing to declare |
| **Spain** | Manuela San Eufrasio Martínez | | Instituto de Biomedicina de Sevilla | Sevilla | | Nothing to declare |
| **Spain** | Laura Muñoz Delgado | | Instituto de Biomedicina de Sevilla | Sevilla | | Nothing to declare |
| **Spain** | Rafael Díaz Belloso | | Instituto de Biomedicina de Sevilla | Sevilla | | Nothing to declare |
| **Spain** | Sergio García Díaz | | Instituto de Biomedicina de Sevilla | Sevilla | | Nothing to declare |
| **Spain** | Marta Bonilla Toribio | | Instituto de Biomedicina de Sevilla | Sevilla | | Nothing to declare |
| **Spain** | Dolores Buiza Rueda | | Instituto de Biomedicina de Sevilla | Sevilla | | Nothing to declare |
| **Spain** | Antonio Cristobal Luque Ambrosiani | | Instituto de Biomedicina de Sevilla | Sevilla | | Nothing to declare |
| **Spain** | Silvia Jesus Maestre | | Instituto de Biomedicina de Sevilla | Sevilla | | Nothing to declare |
| **Spain** | Daniel Macías García | | Instituto de Biomedicina de Sevilla | Sevilla | | Nothing to declare |
| **Spain** | Elena Ojeda Lepe | | Instituto de Biomedicina de Sevilla | Sevilla | | Nothing to declare |
| **Spain** | Rocío Pineda Sánchez | | Instituto de Biomedicina de Sevilla | Sevilla | | Nothing to declare |
| **Spain** | Ana Castellano Guerrero | | Instituto de Biomedicina de Sevilla | Sevilla | | Nothing to declare |
| **Spain** | Astrid Daniela Adarmes Gómez | | Instituto de Biomedicina de Sevilla | Sevilla | | Nothing to declare |
| **Spain** | Cristina Pérez Calvo | | Instituto de Biomedicina de Sevilla | Seville | | Nothing to declare |
| **Spain** | Alejandro Salguero Oviedo | | Instituto de Biomedicina de Sevilla | Sevilla | | Nothing to declare |
| **Spain** | Lorena Clavijo Jiménez | | Instituto de Biomedicina de Sevilla | Sevilla | | Nothing to declare |
| **Spain** | Natalia Lopez | | Global Parkinson's Genetics Program (GP2) | Madrid | | Nothing to declare |
| **Sri Lanka** | Gamini Karapitiya Pathirana | | National Hospital of Sri Lanka | Colombo | | Nothing to declare |
| **Sri Lanka** | Ruwani Wijeyekoon | | Association of Sri Lankan Neurologists | Colombo | | Nothing to declare |
| **Sri Lanka** | Muhammed Saamir Mohideen | | National Hospital Galle | Galle | | Nothing to declare |
| **Sri Lanka** | Manjula Caldera | | Teaching Hospital Anuradhapura | Anuradhapura |  | Nothing to declare |
| **Sri Lanka** | Arjuna Fernando | | National Hospital of Sri Lanka | Colombo | | Nothing to declare |
| **Sri Lanka** | Darshana Sirisena | | CNTH Ragama | Ragama | | Nothing to declare |
| **Sri Lanka** | Senaka Bandusena | | National Hospital of Sri Lanka | Colombo | | Nothing to declare |
| **Sri Lanka** | Thashi Chang | | University of Colombo, Sri Lanka | Colombo | | Nothing to declare |
| **Sri Lanka** | Darshana Dias Wijegunasinghe | | Colombo North Teaching hospital | Colombo | | Nothing to declare |
| **Sri Lanka** | Gamini Pathirana | | NHSL | Colombo | | Nothing to declare |
| **Sri Lanka** | Bimsara Sajotha Nalin Senanayake | | Institution of Neurology, National Hospital of Sri Lanka | Colombo |  | Nothing to declare |
| **Sri Lanka** | Ajantha Keshavaraj | | Teaching Hospital Jaffna | Jaffna | | Nothing to declare |
| **Sudan** | Sarah El-Sadig | | Faculty of medicine university of Khartoum | Khartoum | | Nothing to declare |
| **Sudan** | Inas Elsayed | | Faculty of pharmacy University of Gezira | Wad Madani | | Nothing to declare |
| **Sweden** | Kajsa Brolin | | Lund University | Lund | | Nothing to declare |
| **Sweden** | Per Svenningsson | | Karolinska Institute | Stockholm | | Nothing to declare |
| **Sweden** | Maria Swanberg | | Lund University | Lund | | Nothing to declare |
| **Switzerland** | Christiane Zweier | | Inselspital Bern, University of Bern | Bern | | Nothing to declare |
| **Switzerland** | Gerd Tinkhauser | | University Hospital Bern | Bern | | Nothing to declare |
| **Switzerland** | Paul Krack | | Inselspital Bern, University of Bern | Bern | | Nothing to declare |
| **Switzerland** | Deborah Bartholdi | | University Hospital Bern | Bern | | Nothing to declare |
| **Switzerland** | Jonas Ihle | | Hopital Triemli Zürich | Zürich | | Nothing to declare |
| **Taiwan** | Chin-Hsien Lin | | National Taiwan University Hospital | Taipei City | | Nothing to declare |
| **Taiwan** | Hsiu-Chuan Wu | | Chang Gung Memorial Hospital | Taoyuan City | | Nothing to declare |
| **Taiwan** | Pin-Jui Kung | | National Taiwan University | Taipei City | | Global Parkinson's Genetics Program |
| **Taiwan** | Ruey-Meei Wu | | National Taiwan University Hospital | Taipei City | | Minister of Science and Technology, Taiwan Government; National Taiwan University; Parkinson foundation (USA); Michael J. Fox Foundation |
| **Taiwan** | Yihru Wu | | Chang Gung Memorial Hospital | Taoyuan City | | Nothing to declare |
| **Taiwan** | Pin-Shiuan, Chen | | National Taiwan University Hospital | Taipei | | Nothing to declare |
| **Tajikistan** | Ganieva Manizha | | Avicenna Tajik State Medical University | Dushanbe | | Nothing to declare |
| **Tajikistan** | Maksudjon Isrofilov | | Avicenna Tajik State Medical University | Dushanbe | | Nothing to declare |
| **Tanzania** | Kigocha Okeng'o | | Muhimbili National Hospital-Mloganzila | Dar es Salaam | | Nothing to declare |
| **Tanzania** | Philip Adebayo | | The Aga Khan University | Dar es Salaam | | Nothing to declare |
| **Tunisia** | Rim Amouri | | Mongi Ben Hmida National Institute of Neurolog | Tunis | | Nothing to declare |
| **Tunisia** | Samia Ben Sassi | | Mongi Ben Hmida National Institute of Neurology | Tunis | | Nothing to declare |
| **Tunisia** | Chokri Mhiri | | Habib Bourguiba University Hospital | Sfax | | Nothing to declare |
| **Tunisia** | Nabli Fatnassi Fatma | | National institute Mongi Ben Hmida of Neurology | Tunis | | Nothing to declare |
| **Tunisia** | Amine Rachdi | | Mongi Ben Hamida institute of Neurology | | | Nothing to declare |
| **Tunisia** | Zakaria Saied | | National Institute Mongi Ben Hamida of Neurology | Tunis | | Nothing to declare |
| **Tunisia** | Mouna Ben Djebara | | Razi Hospital | Tunis | | Nothing to declare |
| **Tunisia** | Rania Zouari | | National institute of neurology mongi ben hmida | Tunis | | Nothing to declare |
| **Turkey** | A. Nazlı Başak | | Koç University | Istanbul | | Kirac Foundation and Koc Univ. |
| **Turkey** | Özgür Öztop Çakmak | | Koç University | Istanbul | | Nothing to declare |
| **Turkey** | Sibel Ertan | | Koç University | Istanbul | | Nothing to declare |
| **Turkey** | Rezzak Yilmaz | | University of Ankara | Ankara | | RY received grants from Ankara University, and honoraria/consultancy fees from Abbvie, Gen, Ali Raif, Abdi Ibrahim, and advisory board contribution from Gen, and Abbvie. |
| **Turkey** | Binnur Celik | | University of Ankara | Ankara | | Nothing to declare |
| **Turkey** | Gencer Genc | | Şişli Etfal Training and Research Hospital, University of Health Sciences, İstanbul, TR | Istanbul | | Nothing to declare |
| **Turkey** | Muhittin Cenk Akbostancı | | Private Practice | Ankara | | Nothing to declare |
| **Turkey** | Basar Bilgic | | Istanbul University, Faculty of Medicine | Istanbul | | Nothing to declare |
| **Turkey** | Bedia Samanci | | Istanbul University, Faculty of Medicine | Istanbul | | Nothing to declare |
| **Turkey** | Murat Emre | | Istanbul university | Istanbul | | Nothing to declare |
| **Turkey** | Hasmet Hanağasi | | ISTANBUL FACULTY OF MEDICINE | Istanbul | | Nothing to declare |
| **Turkey** | Aysegul Gunduz | | Istanbul University-Cerrahpasa, Cerrahpasa Faculty of Medicine | Istanbul | | Nothing to declare |
| **Turkey** | Gulcin Benbir Senel | | Istanbul University-Cerrahpasa, Cerrahpasa Faculty of Medicine | Istanbul | | Istanbul University-Cerrahpasa, Cerrahpasa Faculty of Medicine, Department of Neurology, Sleep and Disorders Unit, Istanbul, Turkiye |
| **United Kingdom** | Alastair Noyce | | Queen Mary University of London | London | | Prof. Noyce reports grants from Parkinson's UK, Barts Charity, Cure Parkinson's, NIHR, Innovate UK, Virginia Keiley benefaction, Alchemab, Aligning Science Across Parkinson's and Michael J Fox Foundation. Consultancy and personal fees from Astra Zeneca, AbbVie, Profile, Roche, Biogen, UCB, Bial, Charco Neurotech, uMedeor and Britannia. |
| **United Kingdom** | Anette Schrag | | University College London | London | | NIHR UCL/H Biomedical Research Centre |
| **United Kingdom** | Anthony Schapira | | University College London | London | | ASAP |
| **United Kingdom** | Camille Carroll | | University of Plymouth | Plymouth | | C Carroll receives salary from University of Plymouth, University Hospitals Plymouth NHS Trust and National Institute of Health Research; she has received advisory, consulting, and/or lecture fees from AbbVie, Bial, Lundbeck, Global Kinetics, Britannia and Medscape, and research funding from Parkinson's UK, Edmond J Safra Foundation, National Institute of Health Research and Cure Parkinson's |
| **United Kingdom** | Donald Grosset | | University of Glasgow | Glasgow | | Tracking Parkinson's (J-1101) is funded by Parkinson's UK |
| **United Kingdom** | Eleanor J. Stafford | | University College London | London | | Nothing to declare |
| **United Kingdom** | Henry Houlden | | University College London | London | | Nothing to declare |
| **United Kingdom** | Huw R Morris | | University College London | London | | Dr Morris is employed by UCL. In the last 12 months he reports paid consultancy from Roche and Amylyx ; lecture fees/honoraria - BMJ, Kyowa Kirin, Movement Disorders Society. Research Grants from Parkinson's UK, Cure Parkinson's Trust, PSP Association, CBD Solutions, Drake Foundation, Medical Research Council, Michael J Fox Foundation. Dr Morris is a co-applicant on a patent application related to C9ORF72 - Method for diagnosing a neurodegenerative disease (PCT/GB2012/052140) |
| **United Kingdom** | John Hardy | | University College London | London | | Nothing to declare |
| **United Kingdom** | Kin Ying Mok | | Univeristy College London | London | | Nothing to declare |
| **United Kingdom** | Mie Rizig | | University College London | London | | Nothing to declare |
| **United Kingdom** | Nicholas Wood | | University College London | London | | ASAP- CRN |
| **United Kingdom** | Nigel Williams | | Cardiff University | Cardiff | | Parkinson's UK |
| **United Kingdom** | Olaitan Okunoye | | University College London | London | | Nothing to declare |
| **United Kingdom** | Rauan Kaiyrzhanov | | University College London | London | | Nothing to declare |
| **United Kingdom** | Rimona Weil | | University College London | London | | Nothing to declare |
| **United Kingdom** | Seth Love | | University of Bristol | Bristol | | Medical Research Council (UK) MR/T018569/1 |

| | | | | | | |
|---|---|---|---|---|---|---|
| United Kingdom | Simona Jasaityte | | University College London | London | | Nothing to declare |
| United Kingdom | Sumit Dey | | Queen Mary University of London | London | | The Michael J. Fox Foundation for Parkinson's Research |
| United Kingdom | Spencer Finch | | Queen Mary University of London | London | | Nothing to declare |
| United Kingdom | Valentina Escott-Price | | Cardiff University | Cardiff | | Nothing to declare |
| United Kingdom | Hamin Lee | | St George's, University of London | London | | Nothing to declare |
| United Kingdom | Roger Barker | | University of Cambridge | Cambridge | | Nothing to declare |
| United Kingdom | Mina Ryten | | University College London | London | | Nothing to declare |
| United Kingdom | Michele Hu | | University of Oxford | Oxford | | Nothing to declare |
| United Kingdom | Laura Parkkinen | | University of Oxford | Oxford | | Nothing to declare |
| United Kingdom | Kailash Bhatia | | University College London | London | | Nothing to declare |
| United Kingdom | Richard Walker | | Northumbria Healthcare at NHS Foundation Trust | Newcastle upon Tyne | | Nothing to declare |
| United Kingdom | Steve Gentleman | | Imperial College London | London | | Nothing to declare |
| United Kingdom | Thomas Warner | | University College London | London | | Nothing to declare |
| United Kingdom | David Burn | | Newcastle University | Newcastle upon Tyne | | Nothing to declare |
| United Kingdom | Christian Lambert | | Imperial College London | London | | Nothing to declare |
| United Kingdom | Caroline Williams-Gray | | University of Cambridge | Cambridge | | Nothing to declare |
| United Kingdom | Deborah Attuah | | YLD | London | | Nothing to declare |
| United Kingdom | Raquel Real | | University College London | London | | Nothing to declare |
| United Kingdom | Yen Tai | | Imperial College London | London | | Nothing to declare |
| United Kingdom | Alexandra Zirra | | Queen Mary University of London | London | | Nothing to declare |
| United Kingdom | Christopher M Morris | | Newcastle University | Newcastle upon Tyne | | Nothing to declare |
| United Kingdom | Matilda Lily Fenn | | University College London | London | | Nothing to declare |
| United Kingdom | Andrew C Robinson | | The University of Manchester | Manchester | | Nothing to declare |
| United Kingdom | Lesley Yue Wu | | University College London | London | | Nothing to declare |
| United Kingdom | Tessa Du Toit | | UCL | Londo | | Nothing to declare |
| United Kingdom | Joshua Luc Isherwood Frost | | UCL Queen Square Institute of Neurology | London | | Nothing to declare |
| United Kingdom | Federico Roncaroli | | University of Manchester | Manchester | | Nothing to declare |
| United Kingdom | Ashvin Kuri | | Queen Mary University of London | London | | Nothing to declare |
| United Kingdom | Sheena Waters | | Queen Mary University of London | London | | Nothing to declare |
| United Kingdom | Laura Smith | | Queen Mary University of London | London | | Nothing to declare |
| United Kingdom | Eduardo De Pablo-Fernández | | Queen Mary University of London | London | | Nothing to declare |
| United Kingdom | Anisa Shahid | | Queen Mary University of London | London | | Nothing to declare |
| United Kingdom | Cristina Simonet | | Queen Mary University of London | London | | Nothing to declare |
| United Kingdom | Charlotte Dore | | University College London | London | | Nothing to declare |
| United Kingdom | Oiher Serrano-Asensio | | University College London | London | | Nothing to declare |
| United Kingdom | Marco Toffoli | | University College London | London | | Nothing to declare |
| United Kingdom | Riona Fumi | | University College London, Institute of Neurology | London | | Nothing to declare |
| United Kingdom | Brook Huxford | | Queen Mary University of London | London | | Nothing to declare |
| United Kingdom | Harneek Chohan | | Queen Mary University of London | London | | Nothing to declare |
| United Kingdom | Sophie Meyer | | Queen Mary University of London | London | | Nothing to declare |
| United Kingdom | Laura Pérez-Carbonell | | Queen Mary University London / Guy's and St Thomas' NHS Foundation Trust | London | | Nothing to declare |
| United Kingdom | Solomiia Bandrivska | | University College London | London | | Nothing to declare |
| United Kingdom | Saiesha Dindayal | | University College London | London | | Nothing to declare |
| United Kingdom | Charlotte Andrews | | Queen Mary University of London | London | | Nothing to declare |
| United Kingdom | Emily Navarro Jones | | University College London | London | | Nothing to declare |
| United Kingdom | Kamalesh Dey | | Queen Mary University of London | London | | Nothing to declare |
| United Kingdom | Faiza Durrani | | Centre for Preventive Neurology | London | | Nothing to declare |
| United Kingdom | Marte Jensen | | University College London | London | | Nothing to declare |
| United Kingdom | Yoana Kordovska | | University College London | London | | Nothing to declare |
| United Kingdom | Jennifer Brady | | University College London | London | | Nothing to declare |
| United Kingdom | Nolia Lumley | | University College London | London | | Nothing to declare |
| United Kingdom | Erika Lam | | University College London | London | | Nothing to declare |
| United Kingdom | Tatiana Georgiades | | University College London | London | | Nothing to declare |
| United Kingdom | Kawmadi Wasundera Gunawardena | | Oxford University Hospitals NHS Foundation Trust | Oxford | | Nothing to declare |
| Uruguay | Víctor E. Raggio Risso | | Facultad de Medciina, Universidad de la República | Montevideo | | Nothing to declare |
| Uruguay | Elena M.Dieguez | | Facultad de Medicina universidad de de la tepublica montevideo uruguay | Montevideo | | Nothing to declare |
| USA | Lara M. Lange | | Laboratory of Neurogenetics, National Institute on Aging/Institute of Neurogenetics, University of Luebeck | Bethesda | Maryland | Nothing to declare |
| USA | Angel Vinuela | | University of Puerto Rico | San Juan | | Nothing to declare |
| USA | Alyssa O'Grady | | The Michael J. Fox Foundation for Parkinson's Research | New York | New York | Nothing to declare |
| USA | Andrew B Singleton | | Global Parkinson's Genetics Program (GP2) | Bethesda | Maryland | Michael J Fox Foundation for Parkinson's disease Research and Aligning Science Across Parkinson's Initiative |
| USA | Andrew K. Sobering | | Augusta University / University of Georgia Medical Partnership | Augusta | Georgia | Nothing to declare |
| USA | Bernadette Siddiqi | | The Michael J. Fox Foundation for Parkinson's Research | New York | New York | Nothing to declare |
| USA | Bradford Casey | | The Michael J. Fox Foundation for Parkinson's Research | New York | New York | Nothing to declare |
| USA | Brian Fiske | | The Michael J. Fox Foundation for Parkinson's Research | New York | New York | Nothing to declare |
| USA | Cabell Jonas | | Mid-Atlantic Permanente Medical Group | Bethesda | Maryland | Nothing to declare |
| USA | Carlos Cruchaga | | Washington University | St. Louis | Missouri | National Institutes of Health (R01AG044546 (CC), P01AG003991(CC, JCM), RF1AG053303 (CC), RF1AG058501 (CC), U01AG058922 (CC), RF1AG074007 (YJS)), the Chuck Zuckerberg Initiative (CZI), the Michael J. Fox Foundation (LI, CC), and the Department of Defense (LI- W81XWH2010849). The recruitment and clinical characterization of research participants at Washington University were supported by NIH P30AG066444 (JCM), P01AG03991(JCM), and P01AG026276(JCM). |
| USA | Caroline B. Pantazis | | Coalition for Aligning Science | Bethesda | Maryland | Nothing to declare |
| USA | Claire Wegel | | Indiana University | Bloomington | Indiana | Nothing to declare |
| USA | Cornelis Blauwendraat | | Aligning Science Across Parkinson's (ASAP) | Bethesda | Maryland | Nothing to declare |
| USA | Dan Vitale | | Data Tecnica | Bethesda | Maryland | D.V.'s participation in this project was part of a competitive contract awarded to Data Tecnica International LLC by the National Institutes of Health to support open science research. |
| USA | Deborah Hall | | Rush University | Chicago | Illinois | Nothing to declare |
| USA | Dena Hernandez | | National Institutes of Health | Bethesda | Maryland | Nothing to declare |
| USA | Ekemini Riley | | Coalition for Aligning Science | Washington | Washington | Nothing to declare |
| USA | Faraz Faghri | | Data Tecnica | Bethesda | Maryland | F.F.'s participation in this research was supported in part by the Intramural Research Program of the NIH, National Institute on Aging (NIA), National Institutes of Health, Department of Health and Human Services; project number ZO1 AG000535, as well as the National Institute of Neurological Disorders and Stroke. F.F.'s participation in this project was part of a competitive contract awarded to Data Tecnica International LLC by the National Institutes of Health to support open science research. |
| USA | Geidy E. Serrano | | Banner Sun Health Research Institute | Sun City | Arizona | Banner Sun Health Research Institute Brain and Body Donation Program of Sun City, Arizona for the provision of human biological materials. The Brain and Body Donation Program has been supported by the National Institute of Neurological Disorders and Stroke (U24 NS072026 National Brain and Tissue Resource for Parkinson's Disease and Related Disorders), the National Institute on Aging (P30 AG19610 and P30AG072980, Arizona Alzheimer's Disease Center), the Arizona Department of Health Services (contract 211002, Arizona Alzheimer's Research Center), the Arizona Biomedical Research Commission (contracts 4001, 0011, 05-901 and 1001 to the Arizona Parkinson's Disease Consortium) and the Michael J. Fox Foundation for Parkinson's Research |
| USA | Hampton Leonard | | Data Tecnica | Bethesda | Maryland | H.L.L.'s participation in this project was part of a competitive contract awarded to Data Tecnica International LLC by the National Institutes of Health to support open science research. |
| USA | Hirotaka Iwaki | | Data Tecnica | Washington | Washington | H.I.'s participation in this project was part of a competitive contract awarded to Data Tecnica International LLC by the National Institutes of Health to support open science research. |
| USA | Honglei Chen | | Michigan State University | East Lansing | Michigan | NIH/DoD/Parkinson Foundation/MSU Foundation/Gibby vs. Parky Foundation - No COI to disclose |
| USA | Ignacio F. Mata | | Cleveland Clinic | Cleveland | Ohio | Funding from MJFF and NIH |
| USA | Ignacio Juan Keller Sarmiento | | Northwestern University | Evanston | Illinois | Nothing to declare |
| USA | Jared Williamson | | Kaiser Permanente | Oakland | California | Nothing to declare |
| USA | Jonggeol Jeff Kim | | Baylor College of Medicine | Bethesda | Maryland | Nothing to declare |
| USA | Joseph Jankovic | | Baylor College of Medicine | Houston | Texas | Nothing to declare |
| USA | Joshua Shulman | | Baylor College of Medicine / Texas Children's Hospital | Houston | Texas | Collection of samples and data at Baylor College of Medicine was supported by Huffington Foundation. |

| | | | | | | |
|---|---|---|---|---|---|---|
| USA | J Solle | | The Michael J. Fox Foundation for Parkinson's Research | New York | New York | Nothing to declare |
| USA | Kaileigh Murphy | | The Michael J. Fox Foundation for Parkinson's Research | New York | New York | Nothing to declare |
| USA | Kamalini Ghosh Galvelis | | Parkinson's Foundation | Princeton | New Jersey | Nothing to declare |
| USA | Karen Nuytemans | | University of Miami Miller School of Medicine | Miami | Florida | This work has been supported by the American Parkinson Disease Association and the Margaret Q. Landenberger Research Foundation. |
| USA | Karl Kieburtz | | Beth Israel Deaconess Medical Center | Boston | Massachusetts | Nothing to declare |
| USA | Katerina Markopoulou | | North Shore University Health System | Chicago | Illinois | Nothing to declare |
| USA | Kenneth Marek | | Institute for Neurodegenerative Disorders | New Haven | Connecticut | Consultant for Michael J Fox Foundation, GE Healthcare, Roche, UCB, BIAL, Denali, Takeda, , Cerapsir, UCB, Biohaven, Neuron23, Aprinoia, Astellas, Calico, Genentech, Invicro |
| USA | Kristin S. Levine | | Data Tecnica | Washington | Washington | K.S.L.'s participation in this project was part of a competitive contract awarded to Data Tecnica International LLC by the National Institutes of Health to support open science research. |
| USA | Lana M. Chahine | | University of Pittsburgh | Pittsburgh | Pennsylvania | Nothing to declare |
| USA | Laura Ibanez | | Washington University | St. Louis | Missouri | Nothing to declare |
| USA | Laurel Screven | | Global Parkinson's Genetics Program (GP2) | Bethesda | Maryland | Nothing to declare |
| USA | Lauren Ruffrage | | University of Alabama at Birmingham | Birmingham | Alabama | Nothing to declare |
| USA | Lisa Shulman | | University of Maryland | Baltimore | Maryland | Nothing to declare |
| USA | Luca Marsili | | University of Cincinnati | Cincinnati | Ohio | Nothing to declare |
| USA | Maggie Kuhl | | The Michael J. Fox Foundation for Parkinson's Research | New York | New York | Nothing to declare |
| USA | Marissa Dean | | University of Alabama at Birmingham | Birmingham | Alabama | Dr. Dean is an investigator in studies funded by Abbvie, Inc., Hoffmann-La Roche, CHDI Foundation, Inc., Annexon, Inc., Retrophin, Inc, Neurocrine Biosciences, UniQure Biopharma B.V., Praxis Precision Medicines, Neuraly, Inc., Michael J. Fox Foundation for Parkinson's Research, and US Army Medical Research and Material Command (grant#W81XWH-18-1-0508). In addition, Dr. Dean receives support through the Huntington's Disease Society of American Centers of Excellence program. |
| USA | Mary B Makarious | | Data Tecnica | Bethesda | Maryland | M.B.M.'s participation in this project was part of a competitive contract awarded to Data Tecnica International LLC by the National Institutes of Health to support open science research. |
| USA | Mathew Koretsky | | Data Tecnica | Bethesda | Maryland | M.K.'s participation in this project was part of a competitive contract awarded to Data Tecnica International LLC by the National Institutes of Health to support open science research. |
| USA | Megan J. Puckelwartz | | Northwestern University | Chicago | Illinois | Nothing to declare |
| USA | Mike A. Nalls | | Data Tecnica | Bethesda | Maryland | M.A.N.'s participation in this project was part of a competitive contract awarded to Data Tecnica International LLC by the National Institutes of Health to support open science research. M.A.N. also currently serves as an advisor for Clover Therapeutics and Neuron23 Inc. |
| USA | Naomi Louie | | The Michael J. Fox Foundation for Parkinson's Research | New York | New York | Nothing to declare |
| USA | Niccolò Emanuele Mencacci | | Northwestern University | Evanston | Illinois | Nothing to declare |
| USA | Roger Albin | | Universit of Michigan | Ann Arbor | Michigan | P50NS123067; Parkinson's Foundation |
| USA | Roy Alcalay | | Columbia University | New York | New York | Dr. Alcalay is funded by the Micharl J. Fox Foundation and the Parkinson's Foundation. He received consultation fees from Avrobio, Caraway, GSK, Merck, Sanofi, Ono Therapeutics and Takeda |
| USA | Ruth Walker | | James J. Peters Veterans Affairs Medical Center | New York | New York | Research funding from the Department of Veterans Affairs (CSRD Merit Award CX002342), consulted for Teladoc, Inc. and received an honorarium from New York University (NYU) Medical Center. |
| USA | Sohini Chowdhury | | The Michael J. Fox Foundation for Parkinson's Research | New York | New York | Nothing to declare |
| USA | Sonya Dumanis | | Aligning Science Across Parkinson's | Washington | Washington | Nothing to declare |
| USA | Steven Lubbe | | Northwestern University | Chicago | Illinois | Nothing to declare |
| USA | Tao Xie | | University of Chicago | Chicago | Illinois | Nothing to declare |
| USA | Tatiana Foroud | | Indiana University School of Medicine | Indianapolis | Indiana | The Michael J. Fox Foundation for Parkinson's Research |
| USA | Thomas Beach | | Sun Health Research Institution | Sun City | Arizona | Banner Sun Health Research Institute Brain and Body Donation Program of Sun City, Arizona for the provision of human biological materials. The Brain and Body Donation Program has been supported by the National Institute of Neurological Disorders and Stroke (U24NS072026 National Brain and Tissue Resource for Parkinson's Disease and Related Disorders), the National Institute on Aging (P30 AG19610 and P30AG072980, Arizona Alzheimer's Disease Center), the Arizona Department of Health Services (contract 211002, Arizona Alzheimer's Research Center), the Arizona Biomedical Research Commission (contracts 4001, 0011, 05-901 and 1001 to the Arizona Parkinson's Disease Consortium) and the Michael J. Fox Foundation for Parkinson's Research |
| USA | Todd Sherer | | The Michael J Fox Foundation for Parkinson's Research | New York | New York | Nothing to declare |
| USA | Dana Lewis | | Aligning Science Across Parkinson's | Baltimore | Maryland | Nothing to declare |
| USA | Shreya Menon | | Gladstone Institutes | San Francisco | California | Nothing to declare |
| USA | Melissa Nirenberg | | Icahn School of Medicine at Mount Sinai | New York | New York | Nothing to declare |
| USA | Spencer Grant | | National Institutes of Health | Bethesda | Maryland | Nothing to declare |
| USA | Shannon Ballard | | Data Tecnica | Bethesda | Maryland | S.B.'s participation in this project was part of a competitive contract awarded to Data Tecnica International LLC by the National Institutes of Health to support open science research. |
| USA | Chad Shaw | | Baylor College of Medicine | Houston | Texas | Nothing to declare |
| USA | Sidra Aslam | | Banner Health | Phoenix | Arizona | Nothing to declare |
| USA | Devin Sharp | | Aligning Science Across Parkinson's | Vancouver | British Columbia | Nothing to declare |
| USA | Rachel Saunders-Pullman | | Icahn School of Medicine at Mount Sinai | New York | New York | Nothing to declare |
| USA | Michiko Kimura Bruno | | The Queen's Medical Center | Honolulu | Hawaii | Nothing to declare |
| USA | Matt Farrer | | University of Florida College of Medicine | Gainesville | Florida | Nothing to declare |
| USA | Haydeh Payami | | The University of Alabama at Birmingham Heersink School of Medicine | Birmingham | Alabama | Nothing to declare |
| USA | Ryan Pfingst | | The Michael J Fox Foundation | New York | New York | Nothing to declare |
| USA | James B Leverenz | | Cleveland Clinic | Cleveland | Ohio | Nothing to declare |
| USA | Elizabeth Disbrow | | LSU Health Shreveport | Shreveport | Louisiana | Nothing to declare |
| USA | Debi Brooks | | The Michael J Fox Foundation | New York | New York | Nothing to declare |
| USA | Randy Schekman | | University of California, Berkeley | Berkeley | California | Nothing to declare |
| USA | Un Kang | | NYU Grossman School of Medicine | New York | New York | Nothing to declare |
| USA | Zbigniew K. Wszolek | | Mayo Clinic College of Medicine | Rochester | Minnesota | Nothing to declare |
| USA | Cyrus Zabetian | | VA Puget Sound Health Care System | Seattle | Washington | Nothing to declare |
| USA | Zach Chaney | | The Michael J Fox Foundation | New York | New York | Nothing to declare |
| USA | Christine Swanson-Fischer | | National Institutes of Health | Rockville | Maryland | Nothing to declare |
| USA | Conor Hennessey | | The Michael J Fox Foundation | New York | New York | Nothing to declare |
| USA | Cassandra Barrett | | The Michael J Fox Foundation | New York | New York | Nothing to declare |
| USA | Beate Ritz | | University of California, Los Angeles | Los Angeles | California | Nothing to declare |
| USA | Bradley Boeve | | Mayo Clinic | Rochester | Minnesota | Nothing to declare |
| USA | Ashley Rawls | | University of Florida College of Medicine | Gainesville | Florida | Nothing to declare |
| USA | Holly A. Shill | | Barrow Neurological Institute | Phoenix | Arizona | Nothing to declare |
| USA | Erika Driver-Dunckley | | Mayo Clinic AZ | Scottsdale | Arizona | Nothing to declare |
| USA | Bruce A. Chase | | Endeavor Health (formerly NorthShore University Health System) | Skokie | Illinois | Nothing to declare |
| USA | Mahesh Padmanaban | | University of Chicago | Chicago | Illinois | Nothing to declare |
| USA | Thiago Peixoto Leal | | Cleveland Clinic | Cleveland | Ohio | Thiago Peixoto Leal is supported by the National Institutes of Health (NIH) grant R01 1R01NS112499-01A1 and Veterans Affair Puget Sound Healthcare System grant 5I01ABX005978-2 |
| USA | Owen A. Ross | | Mayo Clinic | Jacksonville | Florida | Nothing to declare |
| USA | Michael Rose | | The Ohio State University Medical Center | Columbus | Ohio | Nothing to declare |
| USA | Ariane Park | | The Ohio State University Medical Center | Columbus | Ohio | I have received grants or contracts from the Parkinson's Foundation, Michael J. Fox Foundation, Parkinson Study Group and honoraria for consultancy from Cerevel. |
| USA | Victoria Klee | | The Ohio State University | Columbus | Ohio | Nothing to declare |
| USA | James C. Beck | | Parkinson's Foundation | New York | New York | Nothing to declare |
| USA | Suzanne Judd | | UAB | Birmingham | Alabama | Nothing to declare |
| USA | Daniel Weintraub | | U. Pennsylvania | Philadelphia | Pennsylvania | Nothing to declare |
| USA | Vikas Kotagal | | University of Michigan | Ann Arbor | Michigan | Nothing to declare |
| USA | Nicolaas I. Bohnen | | University of Michigan | Ann Arbor | Michigan | Nothing to declare |
| USA | Prabesh Kanel | | University of Michigan | Ann Arbor | Michigan | Nothing to declare |
| USA | Chatkaew Pongmala | | University of Michigan | Ann Arbor | Michigan | Nothing to declare |
| USA | Erin Williams | | Van Andel Institute | Grand Rapids | Michigan | Nothing to declare |
| USA | Audrey Strongosky | | Mayo Clinic Florida | Jacksonville | Florida | Nothing to declare |
| USA | Michael Henderson | | Van Andel Institute | Grand Rapids | Michigan | Nothing to declare |
| USA | Daniel C. Rohrer | | Van Andel Institute | Grand Rapids | Michigan | Nothing to declare |
| USA | Alexander Blanski | | Van Andel Research Institute | Grand Rapids | Michigan | Nothing to declare |
| USA | Christina Missler | | Van Andel Institute | Grand Rapids | Michigan | Nothing to declare |
| USA | Alyssa Johansson | | Van Andel Institute | Grand Rapids | Michigan | Nothing to declare |
| USA | Felipe Duarte-Zambrano | | Cleveland Clinic | Cleveland | Ohio | Nothing to declare |
| USA | Gist Croft | | The New York Stem Cell Foundation | New York | New York | Nothing to declare |
| USA | Lisa Voltolina | | New York Stem Cell Foundation Reseach Institute | New York | New York | Nothing to declare |
| USA | Whitley Aamodt | | University of Pennsylvania | Philadelphia | Pennsylvania | Nothing to declare |
| USA | Stewart A Factor | | Emory University | Atlanta | Georgia | Nothing to declare |
| USA | Alberto J. Espay | | University of Cincinnati | Cincinnati | Ohio | Nothing to declare |
| USA | Nabila Dahodwala | | University of Pennsyvlania | Philadelphia | Pennsylvania | Nothing to declare |
| USA | Chantale Branson | | Morehouse School of Medicine | Atlanta | Georgia | Nothing to declare |
| USA | Emily Hill | | University of Cincinnati | Cincinnati | Ohio | Nothing to declare |
| USA | Krutika Patel | | New York Stem Cell Foundation | Denver | Colorado | Nothing to declare |
| USA | Shyamal Mehta | | Mayo Clinic, Arizona | Scottsdale | Arizona | Nothing to declare |
| USA | Emily Waldo | | Cleveland Clinic | Cleveland | Ohio | Nothing to declare |
| USA | Miguel Inca Martinez | | Cleveland Clinic Foundation | Cleveland | Ohio | Nothing to declare |
| USA | Anne-Marie Wills | | Massachusetts General Hospital | Boston | Massachusetts | Nothing to declare |
| USA | Ejaz A. Shamim | | Kaiser Permanente, MidAtlantic Permanente Research Institute | Washington | Washington | Nothing to declare |
| USA | Charles H. Adler | | Mayo Clinic College of Medicine, Mayo Clinic Arizona | Scottsdale | Arizona | Nothing to declare |
| USA | Ileana Lorenzini | | Banner Sun Health Research Institute | Sun City | Arizona | Nothing to declare |
| USA | Peter Heutink | | Global Parkinson's Genetics Program (GP2) | Pacifica | California | Nothing to declare |
| USA | Bernabé Bustos | | Northwestern University Feinberg School of Medicine | Chicago | Illinois | Nothing to declare |
| USA | Shalini Padmanabhan | | Michael J. Fox Foundation | New York City | New York | Nothing to declare |
| USA | Serena Fong | | Michael J. Fox Foundation | New York City | New York | Nothing to declare |
| USA | Vidhu Agarwal | | National Institute of Health | Bethesda | Maryland | Nothing to declare |
| USA | Nicole Kuznetsov | | National Institute of Health | Bethesda | Maryland | Nothing to declare |
| USA | Lietsel Jones | | Data Tecnica International | Bethesda | Maryland | Nothing to declare |
| USA | Julia Staisch | | Ochsner Clinic Foundation | New Orleans | Louisiana | Nothing to declare |
| USA | Camilla Kilbane | | University Hospitals Cleveland Medical Center | Cleveland | Ohio | Nothing to declare |

| | | | | | | |
|---|---|---|---|---|---|---|
| **USA** | Erin Furr Stimming | | The University of Texas Health Science Center at Houston | Houston | Texas | Nothing to declare |
| **USA** | Scott Norris | | Washington University in St. Louis | St. Louis | Missouri | Nothing to declare |
| **USA** | Erica Bell | | The Ohio State University Medical Center | Columbus | Ohio | Nothing to declare |
| **USA** | Natalia Chunga | | Louisiana State University Health Sciences Center at Shreveport | Shreveport | Louisiana | Nothing to declare |
| **USA** | Mohamed Elkasaby | | University Hospitals Cleveland Medical Center/Case Western Reserve University | Cleveland | Ohio | Nothing to declare |
| **USA** | Shivika Chandra | | UTHealth-Houston | Houston | Texas | Nothing to declare |
| **USA** | Erin Foster | | Washington University School of Medicine | St. Louis | Missouri | Nothing to declare |
| **USA** | Vanessa Hinson | | MUSC | Charleston | South Carolina | Nothing to declare |
| **USA** | Scott A. Norris | | Washington University | St. Louis | Missouri | Nothing to declare |
| **USA** | Andrew Nader AMeri | | MUSC | Charleston | South Carolina | Nothing to declare |
| **Vietnam** | Duan Nguyen | | Hue University | Huế | | Nothing to declare |
| **Vietnam** | Toan Nguyen | | Hue University | Huế | | Nothing to declare |
| **Vietnam** | NguyễN Thái Thuỳ Ngân | | University Medical Center, Ho Chi Minh City | Ho Chi Minh | | Nothing to declare |
| **Vietnam** | Ha Ngoc Le Uyen | | University Medical Center Ho Chi Minh city | Ho Chi Minh | | Nothing to declare |
| **Vietnam** | Tai Ngoc Tran | | University Medical Center HCMC | Ho Chi Minh | | Nothing to declare |
| **Vietnam** | Khang Vo | | University Medical Center | Ho Chi Minh | | Nothing to declare |
| **Vietnam** | Vinh Thanh Nguyen | | University Medical Center Ho Chi Minh City | Ho Chi Minh | | Nothing to declare |
| **Vietnam** | Lam Hoang Son | | University Medical Center Ho Chi Minh City | Ho Chi Minh City | | Nothing to declare |
| **Zambia** | Masharip Atadzhanov | | University of Zambia | Lusaka | | Nothing to declare |
